# Deep immune cell phenotyping predicts influenza vaccine immunogenicity in HCT recipients

**DOI:** 10.64898/2026.11.01.26367040

**Authors:** Amir Asiaee, Samhita Pal, Joshua D. Simmons, Andrew J. Spieker, James D. Chappell, Natasha B. Halasa, Spyros A. Kalams, Influenza Vaccine in HCT Study Group

**Author notes:** Corresponding author: Spyros A. Kalams, MD, Division of Infectious Diseases, Department of Medicine, Vanderbilt University Medical Center, Nashville, TN, USA.

## Abstract

Annual influenza vaccination is recommended after allogeneic hematopoietic cell transplantation (HCT), but antibody responses vary widely. We asked whether pre-vaccination immune-cell composition predicts the antibody response beyond baseline titers and clinical factors. We studied the first vaccination season of 260 participants in 2 multicenter phase 2 trials of high- versus standard-dose influenza vaccine, 1 pediatric and 1 adult, and held out for validation 74 second-season episodes from participants who re-enrolled the following year. Thirty-marker mass cytometry measured blood cell composition, and models predicted the mean antibody log fold-rise across 3 antigens in recipients left out of nested cross-validation. Adding immune-cell composition to baseline antibody titers reduced mean squared prediction error by 15.3% (95% CI, 7.0–23.2%; *P* < .001), and adding it to titers plus clinical factors by 12.5% (4.6–20.3%; *P* = .001). A cellular composition score identified recipients unlikely to respond: in its lowest fifth, 15% had a fourfold antibody rise against 51% of the others. At a given baseline titer, each standard deviation of the score about doubled the odds of a fourfold rise (odds ratio, 2.2; *P* = .007), in children and adults alike; low scores reflected fewer B cells and more monocytes. In the held-out second season, composition reduced prediction error by 39%, and prediction was similar with 15 markers. Pre-vaccination immune-cell composition therefore predicts influenza vaccine immunogenicity after HCT, and simpler assays merit prospective validation. The trials are registered at www.clinicaltrials.gov as #NCT02860039 (pediatric) and #NCT03179761 (adult).

**Visual Abstract:** 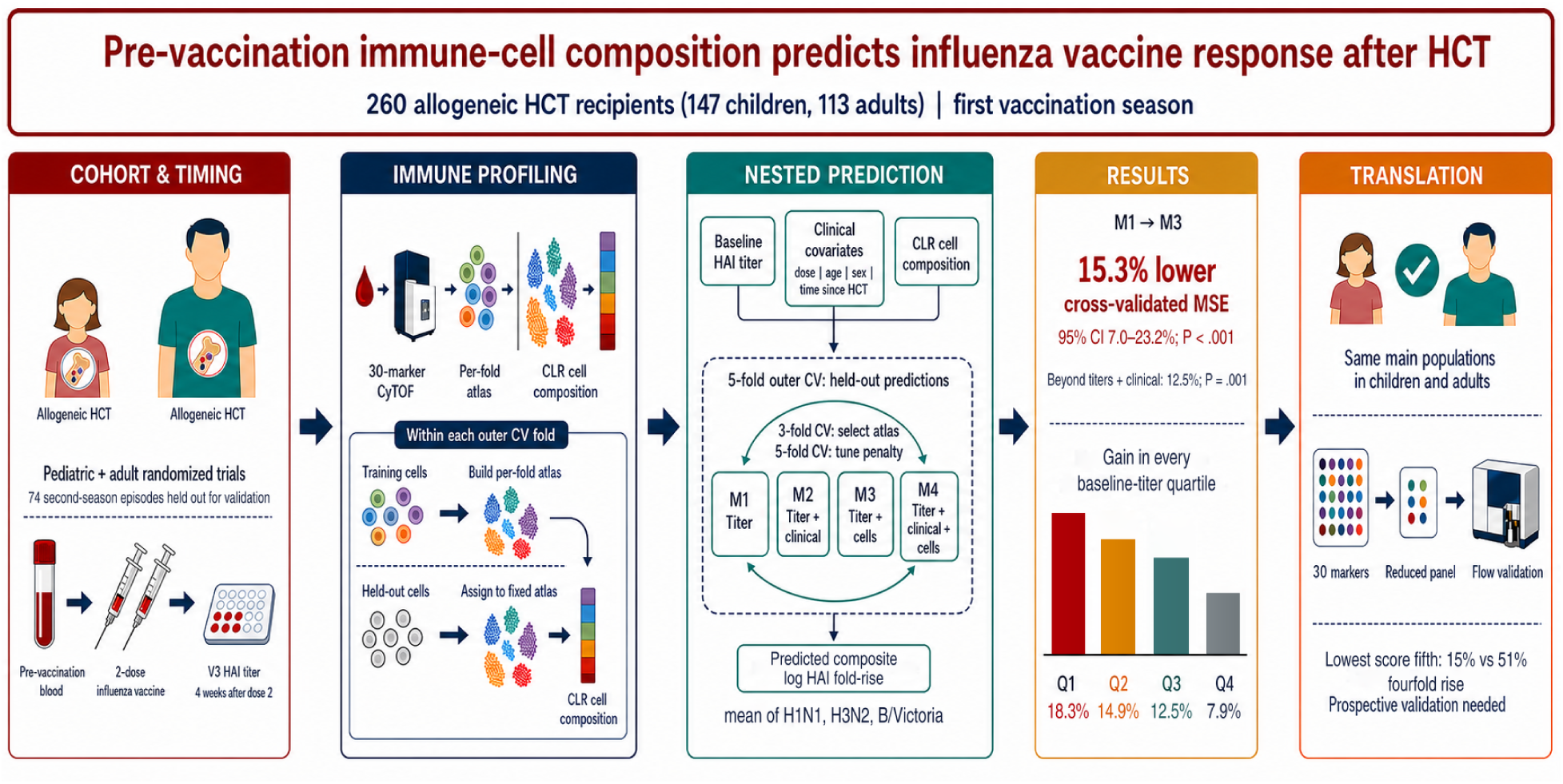

**Key Points:**

- Pre-vaccination immune-cell composition improves prediction of the influenza vaccine response after HCT beyond baseline titers.
- A low cellular composition score identified recipients unlikely to respond to vaccination, in children and adults alike.

## 1 Introduction

Allogeneic hematopoietic cell transplantation (HCT) is a curative therapy for hematologic malignancies and bone marrow failure syndromes, but recipients remain at substantially elevated risk of severe infection for months to years after transplantation because of the slow, incomplete, and variable recovery of adaptive immunity.^1–5^ Annual influenza vaccination is recommended for all HCT recipients, yet serologic responses are highly variable and a meaningful fraction of recipients fail to mount a fourfold rise in hemagglutination inhibition (HAI) titer to any included antigen.^6–12^ The transplant physician facing a recipient at a routine clinical encounter has no pre-vaccination tool to identify recipients at greatest risk of a weak antibody response, and therefore no rational basis on which to time revaccination, prioritize booster doses, or recommend high-dose formulations in those most likely to benefit.^13,14^

Immune reconstitution after transplantation shapes both susceptibility to infection and the strength of the vaccine response.^15^ The clinical correlates of a poor response are known in aggregate (short time since transplantation, active graft-versus-host disease, ongoing immunosuppression^16–20^), but they explain only part of the between-recipient variability and do not yield a usable pre-vaccination tool. We hypothesized that the capacity to mount a serologic vaccine response is reflected in part in the composition of the peripheral lymphoid and myeloid compartments at the time of vaccination, and that this composition carries information that clinical factors alone do not.

Direct measurement of that composition has become feasible with single-cell techniques. In the general population, systems-vaccinology studies have shown that the pre-vaccination immune state predicts the antibody response to influenza and other vaccines,^21–26^ but this baseline-state framework has not been established after allogeneic transplantation. In pediatric HCT recipients enrolled in a multi-center vaccine trial, Amarin and colleagues used mass cytometry to show that the pre-vaccination frequencies of selected B and CD4^+^ T cell subpopulations are associated with the subsequent influenza HAI response.^27^ That analysis was a single-cohort study of univariate associations, and an association is not yet a prediction: a population that differs between responders and non-responders may still add nothing once the pre-vaccination titer is known.

We therefore asked whether the whole pre-vaccination cellular composition predicts the antibody response in recipients the model has never seen, and whether it does so beyond baseline titers and the clinical factors already available at the visit. We studied the first vaccination season of 260 pediatric and adult recipients from 2 multicenter phase 2 trials, profiled by 30-marker mass cytometry before vaccination, and summarized the models as one cellular composition score per recipient, so that the gain could be read as a difference in antibody response. We then asked whether that score identifies recipients unlikely to respond, whether children and adults share the same cellular profile, whether the score predicts response a season later in the same recipients, and whether fewer populations and markers carry the same information.

## 2 Methods

### 2.1 Study population

We analyzed the first vaccination season of 260 allogeneic HCT recipients in 2 parallel randomized trials, 1 pediatric (*n* = 147) and 1 adult (*n* = 113). Both trials compared high-dose with standard-dose inactivated influenza vaccine in a 2-dose regimen,^28–31^ and their primary outcomes are published.^9–11^ A vaccination episode is one recipient’s course in one study year: a pre-vaccination sample at visit V1, the day of vaccination, paired with the titer at V3, 4 weeks after the second dose. The 74 episodes contributed by recipients who re-enrolled the following year were held out and used only for validation (Section 2.7). Episodes without a recorded V1 or V3 titer were excluded (2 first-season and 7 second-season episodes). Pre-vaccination blood was profiled by 30-marker mass cytometry^32,33^ with the panel of Amarin et al.,^27^ yielding about 9.9 million first-season cells, a median of about 40,000 per recipient. Marker intensities were arcsinh-transformed (cofactor 5) and corrected for acquisition batch (58 run dates, both cohorts together) with Harmony^34^ before analysis. Both trials were approved by the institutional review boards of the participating centers, and all participants or their guardians gave written informed consent. A.A. and S.P. analyzed the data, and all authors had access to the primary trial data.

### 2.2 Outcome

The response to each antigen was the log fold-rise in hemagglutination inhibition (HAI) titer from V1 to V3, for H1N1, H3N2 and B/Victoria. Each titer was the geometric mean of the duplicate measurements recorded in the trial databases. Samples below the assay detection limit (<1:10) were assigned a titer of 1:5 and samples above the ceiling the ceiling value. B/Yamagata was excluded because it is not in the high-dose formulation for adults.

The primary outcome was the composite response, the mean of the 3 antigen log fold-rises, whose exponential is the geometric mean fold-rise across antigens. One outcome per recipient answers the clinical question, whether this recipient will respond to this vaccination, with the information in all 3 antigens and without testing 3 correlated endpoints separately. A fourfold composite rise is a geometric mean fold-rise of at least 4, which a recipient can meet with a large rise to 1 or 2 antigens and not the third. A fourfold rise is a serologic response criterion and does not directly measure protection against clinical influenza.^35,36^ Models for the 3 antigens separately are in supplemental Section S2.5.

### 2.3 Prediction models

Four models differ only in their predictor blocks, so their differences isolate what each block adds: M1, the log pre-vaccination titer of each of the 3 antigens; M2, M1 plus vaccine dose, age at consent, sex and months since transplantation; M3, M1 plus the cellular-composition features; and M4, all of them. Each model was an elastic-net penalized linear regression (*α* = 0.9) of the composite response, with the penalty chosen by cross-validation inside each training fold. Using one penalized model family for all 4 models keeps the penalty from being confounded with the added predictors; fitting M1 and M2 by ordinary least squares instead changed their error by less than 0.2%. Supplemental Methods explains why the pre-vaccination titer can be a predictor of a fold-rise that contains it.

### 2.4 Cellular composition features

Thirty markers allow cells to be grouped at many levels of detail, and the level that best predicts the antibody response is not known in advance. We therefore treated the clustering resolution as a tuning parameter and let predictive performance choose it.

Within each outer training fold, *k*-means grouped one million cells into 5,000 micro-clusters and Leiden community detection grouped the micro-cluster centers into the cell populations used as features. Training and test cells were assigned to the nearest micro-cluster and inherited its population label, so a held-out recipient’s cells influenced neither the populations nor the model. Each recipient’s population fractions entered the models as centered log-ratios, which express each fraction relative to the geometric mean of all fractions in that sample. The resolution, and with it the number of populations, was chosen by 3-fold recipient-grouped cross-validation inside the training fold, by the error of M4 for the composite response relative to an unpenalized titers-plus-clinical model. The selected numbers of populations were 42, 35, 42, 37 and 25 in the 5 folds, and prediction changed modestly across the grid (supplemental Methods; supplemental Figure S1).

For interpretation we also built one shared atlas of 35 populations from the cells of both cohorts and annotated it from marker profiles. The shared atlas is used only to name and compare populations, never to fit the primary models.

### 2.5 Cellular composition score

The cellular composition score is one number per recipient: the part of the predicted composite log fold-rise that comes from immune-cell composition. It is the sum of the recipient’s population features, each multiplied by its coefficient in the M3 model fitted without that recipient, excluding the intercept and the baseline-titer terms. The score is high when a recipient’s blood holds relatively more of the populations that accompanied strong responses in other recipients and relatively less of those that accompanied weak ones. Because each fold has its own atlas, scores were standardized within fold (mean 0, standard deviation [SD] 1) before pooling; 1 SD corresponded to a 1.9-fold difference in predicted fold-rise. Standardized scores ranged from −2.8 to 2.1 (5th to 95th percentile, −1.7 to 1.5).

### 2.6 Statistical analysis

Performance was evaluated by 5-fold outer cross-validation with folds grouped by recipient, and the mean squared error (MSE) averaged across folds, weighted by the number of test recipients. Model comparisons used a recipient-level bootstrap of the out-of-fold prediction error (*B* = 2,000, fixed seed), with the 95% CI the 2.5th to 97.5th percentile and *P* two-sided. We summarized the score in fifths, estimated its association with a fourfold composite rise by logistic regression adjusted for baseline titer (odds ratio per SD) and with the observed composite fold-rise by Spearman correlation, and compared the area under the receiver operating characteristic curve (AUC) of the titers-only and titers-plus-composition models for a fourfold composite rise. The *P* values for these score statistics come from a null bootstrap that repeats resolution selection, penalty tuning and coefficient fitting in each of 999 replicates generated under no cellular effect beyond baseline titers, so they account for the whole fitting pipeline; conventional Wald and DeLong tests are reported beside them (supplemental Table S1). All *P* values are 2-sided. Analyses used R 4.6.1 with `glmnet`^37^ and `igraph`;^38^ the analysis scripts accompany the study. Mechanics of the atlas, the bootstrap and the null bootstrap are in supplemental Methods.

### 2.7 Second-season validation

Each second-season episode was assigned to the cross-validation fold of its recipient, mapped to that fold’s atlas and scored with that fold’s models, which were fitted without the recipient. Neither season of a recipient contributed to the model that scored it. In these episodes we compared the prediction error and AUC of the titers-only and titers-plus-composition models and, among recipients with both seasons, the change in score and in baseline titer by response in each season. We also compared the response 4 weeks after the first dose between seasons. The same pipeline run on all 334 episodes with complete titers, with both seasons of a returning recipient in the same fold, is a sensitivity analysis (supplemental Section S3.7).

## 3 Results

### 3.1 Recipient characteristics

The 260 recipients had a median age of 16.6 years (range, 3.1–72.8), 147 in the pediatric trial (median, 11.8 years) and 113 in the adult trial (median, 57.9 years), and were vaccinated a median of 6.3 months after transplantation (range, 3.0–35.9). Half received the high-dose formulation (130 of 260). Before vaccination the geometric mean HAI titer across the 3 antigens was 46, and 114 recipients (44%) achieved a fourfold composite rise. Pre-vaccination blood yielded a median of 40,000 profiled cells per recipient. Cohort characteristics are in Table 1.

**Table 1:**
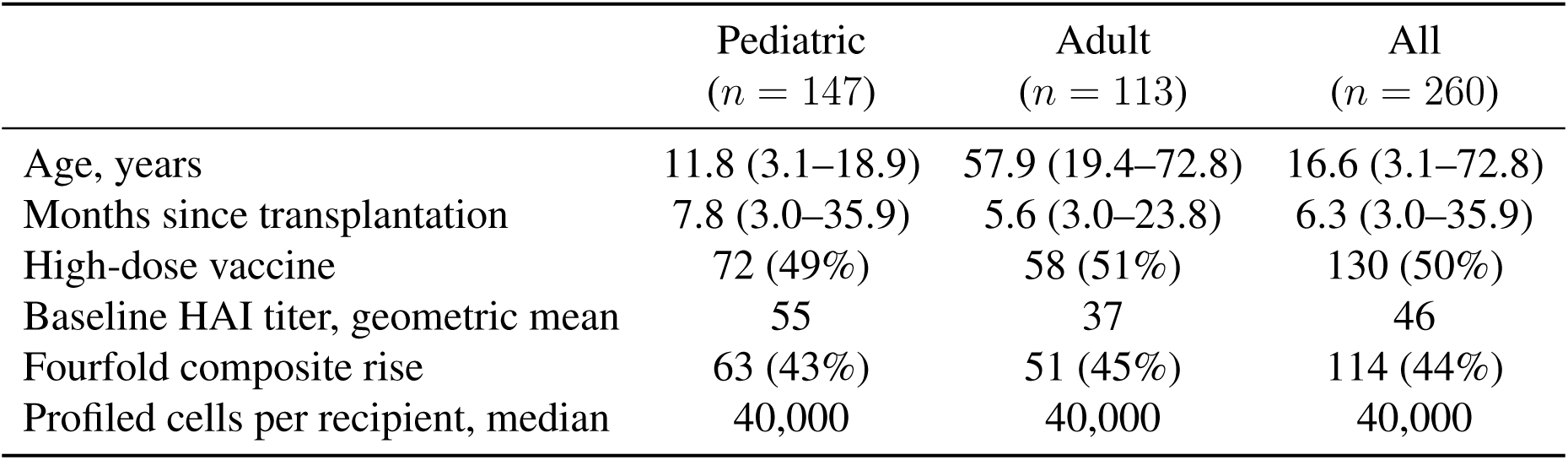
Characteristics of the 260 recipients. Values are median (range), *n* (%), or as labeled. Months since transplantation and the baseline titer are measured at the vaccination visit; the baseline titer is the geometric mean HAI titer across H1N1, H3N2 and B/Victoria. A fourfold composite rise is a geometric mean fold-rise of at least 4 from the day of vaccination to 4 weeks after the second dose.

|  | Pediatric<br>(*n* = 147) | Adult<br>(*n* = 113) | All<br>(*n* = 260) |
|---|---|---|---|
| Age, years | 11.8 (3.1–18.9) | 57.9 (19.4–72.8) | 16.6 (3.1–72.8) |
| Months since transplantation | 7.8 (3.0–35.9) | 5.6 (3.0–23.8) | 6.3 (3.0–35.9) |
| High-dose vaccine | 72 (49%) | 58 (51%) | 130 (50%) |
| Baseline HAI titer, geometric mean | 55 | 37 | 46 |
| Fourfold composite rise | 63 (43%) | 51 (45%) | 114 (44%) |
| Profiled cells per recipient, median | 40,000 | 40,000 | 40,000 |

### 3.2 The shared atlas resolved 35 immune-cell populations

To give the cellular predictors a biological interpretation, we constructed a shared 35-population atlas using cells from both cohorts and annotated its populations from marker profiles (supplemental Table S2). This provides common cell labels; prediction in the primary analysis used atlases constructed separately within each outer training fold. Because the panel lacked dedicated myeloid and natural killer (NK)-cell markers (CD14, CD16, CD11c, CD123, CD56), antigen-presenting cell (APC) and monocyte labels were inferred by exclusion (CD3^−^CD19^−^HLA-DR^+^), and so was the NK-like label. The 35 populations comprised 9 B-cell populations (including naive, transitional, cycling, CD21-low and CCR7^−^ B cells and plasmablasts), 7 CD4^+^ and 6 CD8^+^ T-cell populations, 2 double-negative (CD4^−^CD8^−^) T-cell populations, 6 monocyte and APC populations, 4 lineage-negative populations and 1 NK-like population (Figure 1B; supplemental Figures S2 and S3). Memory T-cell subsets were named from CD45RO and CCR7; because the panel has no CD45RA, effector-memory cells re-expressing CD45RA (EMRA) were identified as CD45RO^−^CCR7^−^. No B-cell population had the CD27^+^ phenotype of memory B cells. We labeled populations activated when they expressed activation markers (CD69, CD38, OX40 or CD137) above other populations of the same lineage, and cycling when they expressed Ki-67. For example, the double-negative T cells of cluster 32 had the highest CD38 and TIGIT expression of any T-cell population (supplemental Figure S3).

**Figure 1:**
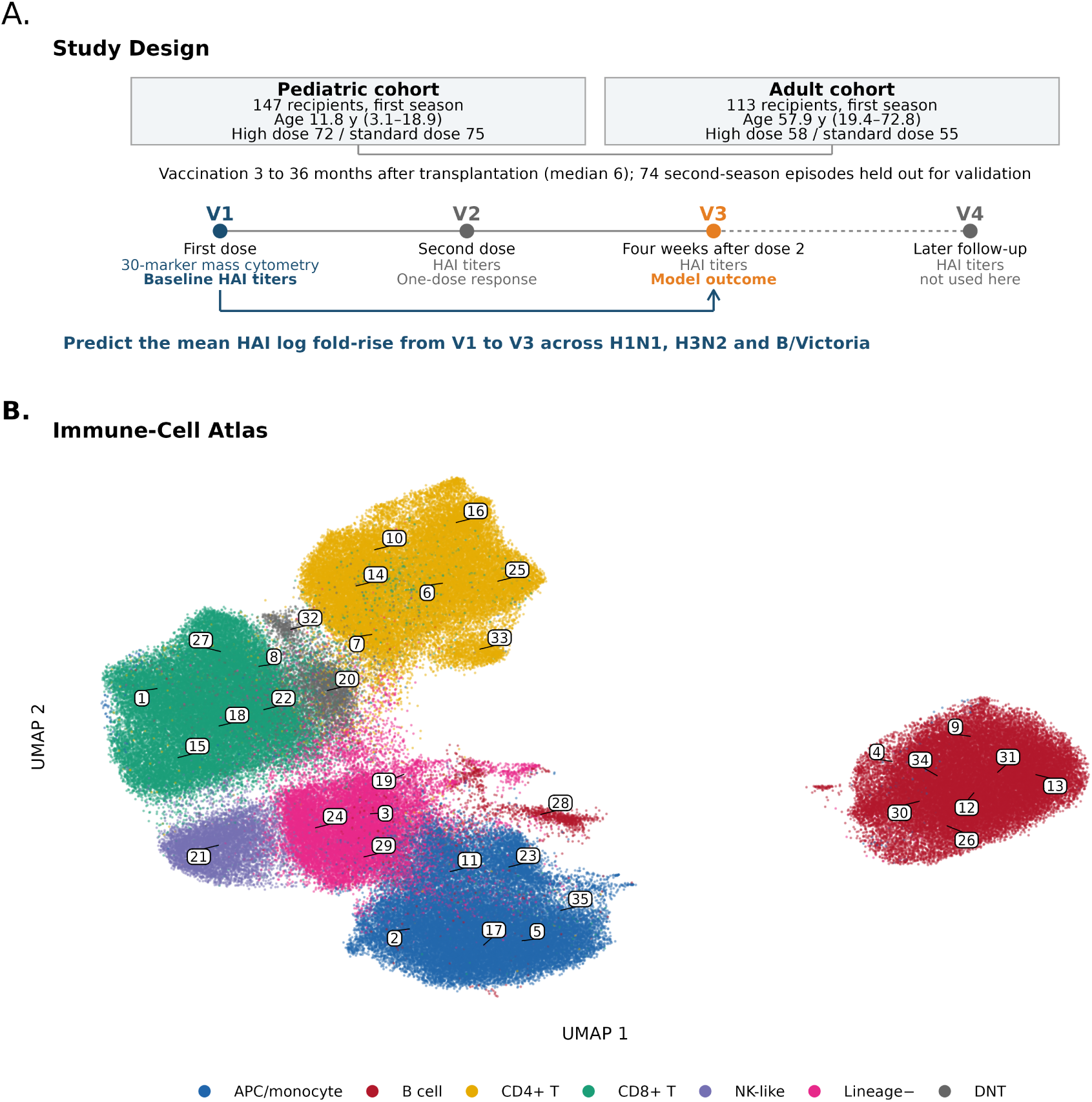
Study design and pre-vaccination immune-cell populations. **A.** The 260 first-season recipients came from two parallel randomized trials of two doses of high-dose versus standard-dose inactivated influenza vaccine, one pediatric and one adult. Ages are medians with ranges, and vaccination occurred 3 to 36 months after transplantation (median 6). Seventy-four second-season episodes of recipients who re-enrolled the following season were held out for validation. At V1 the first dose was given and pre-vaccination blood was profiled by 30-marker mass cytometry; that profile and the baseline HAI titers are the model inputs. The outcome is the mean log fold-rise in HAI titer, log(V3/V1), across H1N1, H3N2 and B/Victoria, where V3 is 4 weeks after the second dose. Titers at the second-dose visit (V2) are used only in the second-season comparison, those at a later follow-up (V4) not at all, and B/Yamagata was excluded because it is not in the high-dose formulation for adults. Visit spacing is schematic. **B.** Each point is a cell, placed by UMAP according to its 30-marker cytometry profile. Colors indicate the annotated lineage and numbers identify the 35 populations of the shared atlas built from both cohorts. The models use each recipient’s share of cells in each population. APC, monocyte and NK-like labels are provisional because the panel lacks dedicated myeloid and NK-cell markers. DNT, double-negative T cells.

### 3.3 Adding immune-cell composition improved prediction beyond baseline titers

The four-model comparison isolates the added predictive value of immune-cell composition. Model accuracy is reported as mean squared error (MSE), the average squared difference between the predicted and observed composite log fold-rise; a percentage improvement is the relative reduction in that error, not a proportion of recipients.

Adding immune-cell composition to baseline titers (M1→M3) reduced MSE from 2.277 to 1.929, an improvement of 15.3% (95% recipient-bootstrap CI, 7.0–23.2%; *P* < .001; Table 2, Figure 2). Clinical factors alone reduced MSE by 6.0% (M1→M2; *P* = .072), and adding immune-cell composition on top of them still improved prediction by 12.5% (M2→M4; 4.6–20.3%; *P* = .001). Adding clinical factors to a model that already had cellular composition gave 3.0% (*P* = .20), and the two information sources together improved prediction by 17.8% over titers alone (M1→M4; 8.7–26.5%; *P* = .001). Cellular composition therefore carried information that the clinical factors did not, while the reverse was less clear.

**Figure 2:**
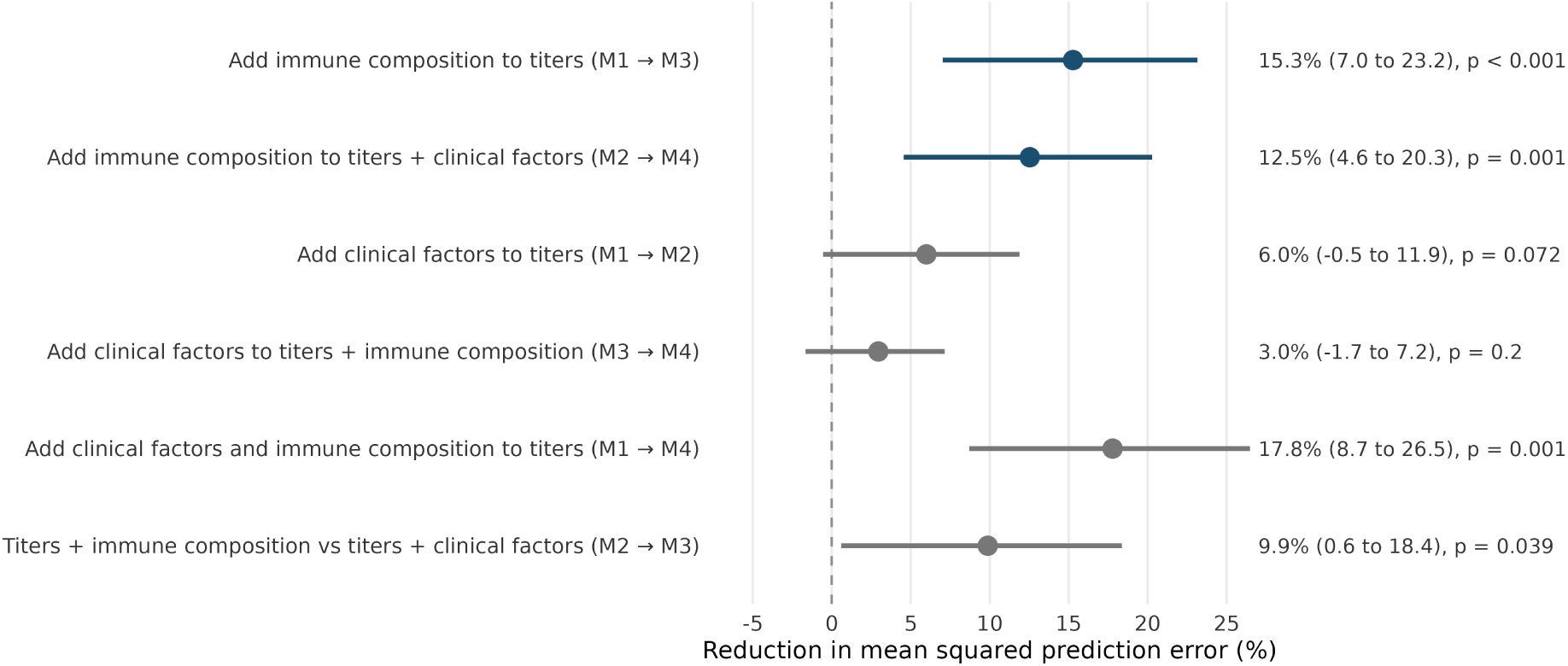
Added predictive value of immune-cell composition. Points show the percentage reduction in mean squared error of the predicted composite antibody response between models; positive values favor the second model. Navy marks the two comparisons that isolate immune-cell composition. Models contain baseline titers (M1), titers and clinical factors (M2), titers and immune-cell composition (M3), or all three (M4). Predictions were evaluated in recipients excluded from model fitting, with atlas construction and tuning performed inside training folds. Whiskers are 95% intervals from 2,000 paired bootstrap samples that resample whole recipients. All comparisons use the same 260 recipients.

**Table 2:** Four-model ladder: prediction of the composite antibody response. A. Mean squared error (MSE) of the out-of-fold predicted composite log fold-rise, from 5-fold nested cross-validation; lower is better. All models are elastic nets (*α* = 0.9). **B.** Pairwise comparisons. ΔMSE% is the relative reduction in MSE, so positive values mean the richer model predicts better. Intervals are paired bootstrap (*B* = 2,000) resampling whole recipients; *P* is two-sided.

| A. Prediction error by model |  |  |  |
|---|---|---|---|
| Model | Predictors | MSE |  |
| M1 | Baseline titers | 2.277 |  |
| M2 | Titers + clinical factors | 2.140 |  |
| M3 | Titers + immune-cell composition | 1.929 |  |
| M4 | Titers + clinical factors + composition | **1.872** |  |
| B. Pairwise model comparisons |  |  |  |
| Comparison | ΔMSE (%) | 95% CI | *P* |
| M1→M3 (composition over titers) | 15.3 | 7.0 to 23.2 | <.001 |
| M1→M4 (full over titers) | 17.8 | 8.7 to 26.5 | .001 |
| M2→M4 (composition over clinical) | 12.5 | 4.6 to 20.3 | .001 |
| M2→M3 (composition vs clinical) | 9.9 | 0.6 to 18.4 | .039 |
| M1→M2 (clinical over titers) | 6.0 | −0.5 to 11.9 | .072 |
| M3→M4 (clinical on top of composition) | 3.0 | −1.7 to 7.2 | .196 |

Models fitted to the 3 antigens separately gave the same picture with smaller intervals: composition improved prediction by 11.2% beyond titers (4.6–17.6%; *P* = .003) and by 8.6% beyond titers plus clinical factors (*P* = .006), with per-antigen gains of 8.7% for H1N1, 12.4% for H3N2 and 12.2% for B/Victoria (supplemental Section S2.5, Table S3).

### 3.4 B-cell, T-cell and monocyte populations drove the prediction

To see which populations the model used, we refitted it on the shared atlas in 200 bootstrap resamples of recipients. Ten of the 35 populations were selected in at least half of the refits (Figure 3; supplemental Figures S4, S5 and S6), including all 4 of the rarest populations (supplemental Figure S7).

**Figure 3:**
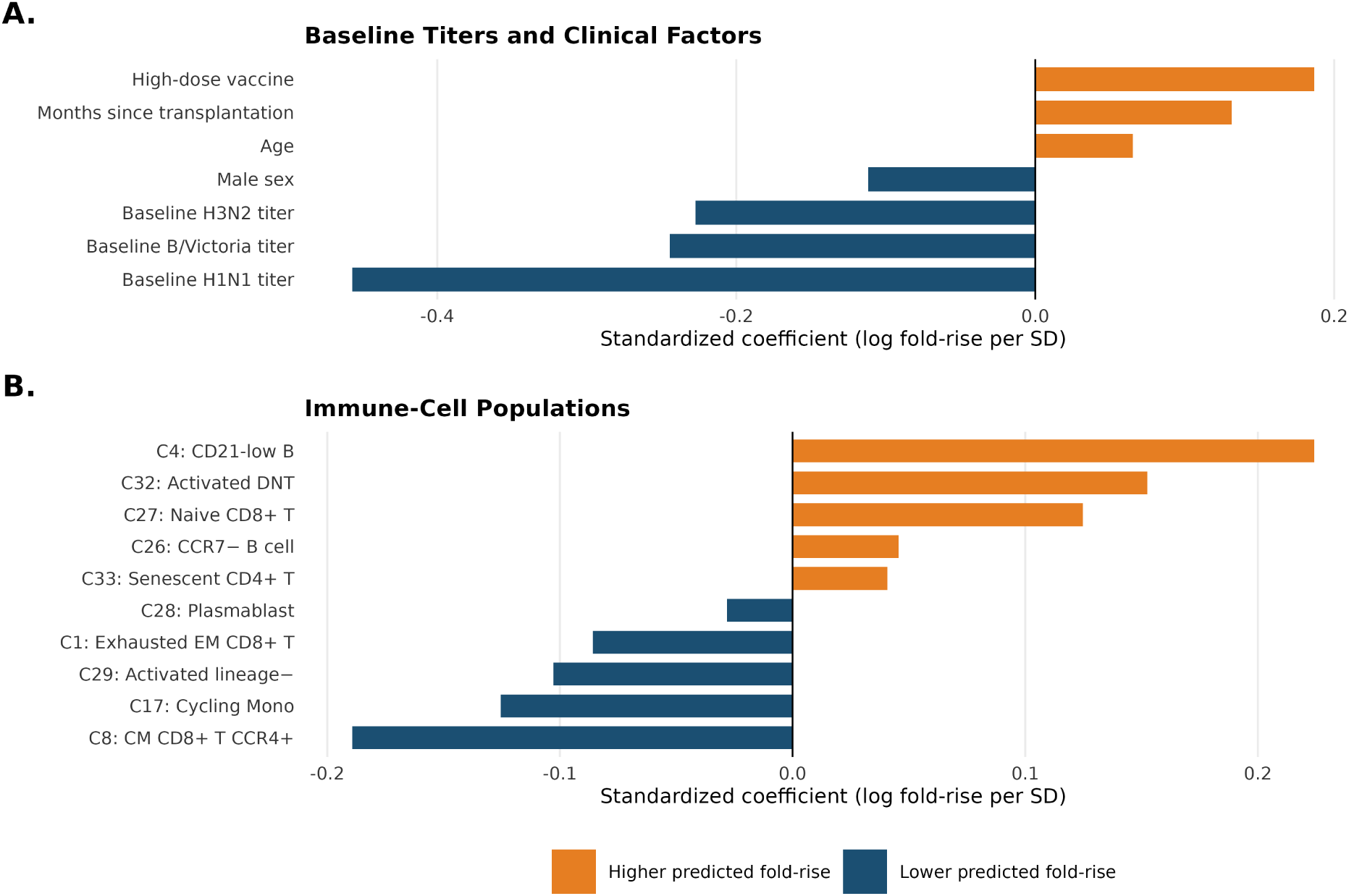
Predictors of the composite antibody response. Bars show standardized coefficients of the model containing baseline titers, clinical factors and immune-cell composition (M4), fitted on the shared 35-population atlas in 200 bootstrap resamples of recipients: the coefficient averaged across refits, multiplied by the predictor’s standard deviation. **A.** Baseline titers and clinical factors. **B.** The ten immune-cell populations selected in at least half of the refits (CM, central memory; DNT, double-negative T cells; EM, effector memory; Mono, monocytes). Positive values indicate a higher predicted composite log fold-rise and negative values a lower one, accounting for the other predictors. A negative baseline-titer coefficient means that a higher starting titer predicts a smaller proportional rise, not a lower post-vaccination titer. Horizontal scales differ between A and B.

Five were associated with a larger antibody response: B cells with low CD21 expression (CD21-low; cluster 4), activated double-negative T cells with high CD38 and TIGIT expression (cluster 32), naive CD8^+^ T cells (cluster 27), CCR7^−^ B cells (cluster 26) and senescent CD4^+^ T cells (CD57^+^; cluster 33). Five were associated with a smaller response: central-memory CD8^+^ T cells expressing CCR4 (cluster 8), cycling monocytes (Ki-67^+^; cluster 17), activated lineage-negative cells expressing CD38, CD138 and CXCR3 (cluster 29), exhausted effector-memory CD8^+^ T cells (PD-1^+^TIGIT^+^; cluster 1) and plasmablasts (cluster 28). Population 8 is heterogeneous: about half of its cells are CD8^+^CD4^−^, a fifth CD4^+^CD8^−^and a quarter CD4^−^CD8^−^, with dim CD8 and high CD69, HLA-DR, CXCR3 and CCR4, so it is best read as an activated T-cell population rather than a single CD8^+^ subset. Baseline-titer terms remain among the largest coefficients; the cellular coefficients describe associations conditional on those titers and the clinical factors.

Cluster 29 carries CD38, CD138 and CXCR3 without clear B- or T-cell lineage markers, so we could not assign it a cell type. On the shared atlas, removing the four lineage-negative populations that lack lineage markers (clusters 3, 19, 24 and 29) and recomputing composition from the remaining 31 did not reduce the gain beyond baseline titers (18.6% versus 18.1% with all 35 shared-atlas populations; supplemental Section S2.6), so the prediction does not depend on these unassigned cells.

### 3.5 Prediction gains were present in every baseline-titer quartile

A high pre-vaccination titer has been associated with a smaller fold-rise after vaccination in several studies,^22,39^ and recipients with low baseline titers are the ones at most risk, so we asked where the gain came from across the baseline range of HAI titers.

Immune-cell composition improved prediction in all four baseline-titer quartiles, by 18.3% in the lowest, 14.9% and 12.5% in the middle two and 7.9% in the highest (Table 3). The gain was largest where it matters most, among recipients whose pre-vaccination titers leave the most room to respond, and it fell steadily as baseline titers rose.

**Table 3:** Prediction by baseline-titer quartile. Mean squared error of the out-of-fold predicted composite log fold-rise for the model with baseline titers (M1) and the model that adds immune-cell composition (M3), within quartiles of the geometric mean baseline titer across the three antigens. The interval is a recipient-level bootstrap (*B* = 2,000).

| Quartile | Titer range | *n* | M1 MSE | M3 MSE | Improvement, % (95% CI) |
|---|---|---|---|---|---|
| Q1 (low baseline) | 5–14 | 67 | 4.049 | 3.306 | 18.3 (5.4 to 30.7) |
| Q2 | 16–45 | 64 | 2.160 | 1.838 | 14.9 (−0.6 to 30.8) |
| Q3 | 50–127 | 66 | 1.872 | 1.638 | 12.5 (−5.2 to 27.9) |
| Q4 (high baseline) | 143–6451 | 63 | 0.935 | 0.861 | 7.9 (−27.8 to 35.9) |

### 3.6 A low cellular composition score identified recipients unlikely to respond

A reduction in squared error is hard to act on at the bedside, so we summarized the same models as one number per recipient: the cellular composition score, the part of the predicted response that comes from immune-cell composition (Section 2.5). Standardized scores ranged from −2.8 to 2.1 SD.

A low score corresponds to a recognizable blood composition (Figure 4A). Compared with recipients in the highest fifth of the score, those in the lowest fifth had a median B-cell share about about one-eighth as large (3.8% versus 28.9% of PBMCs, with plasmablasts counted as B cells) and about half the share of CD4^+^ T cells, but about 3.8 times the share of monocytes and other antigen-presenting cells and 2.4 times the share of lineage-negative cells (supplemental Figure S8). Fourfold composite rises became more frequent from the lowest to the highest fifth of the score, from 15% (8 of 52 recipients) to 67% (35 of 52; Figure 4B). Outside the lowest fifth, 51% of recipients had a fourfold rise (difference, 36 percentage points; 95% CI, 23–47; *P* = .001). At a given baseline titer, each SD of the score was associated with 2.2-fold higher odds of a fourfold rise (95% CI, 1.6–3.0; *P* = .007) and a 1.9-fold larger geometric mean fold-rise (1.6–2.2). Figure 4C shows this across the range of baseline titers: at any baseline titer, recipients with lower scores were less likely to respond. Among recipients with a pre-vaccination HAI titer below 1:40, 24% to 50% of those in the lowest fifth reached 1:40, depending on antigen, against 71% to 87% of the others (supplemental Figure S9B).

**Figure 4:**
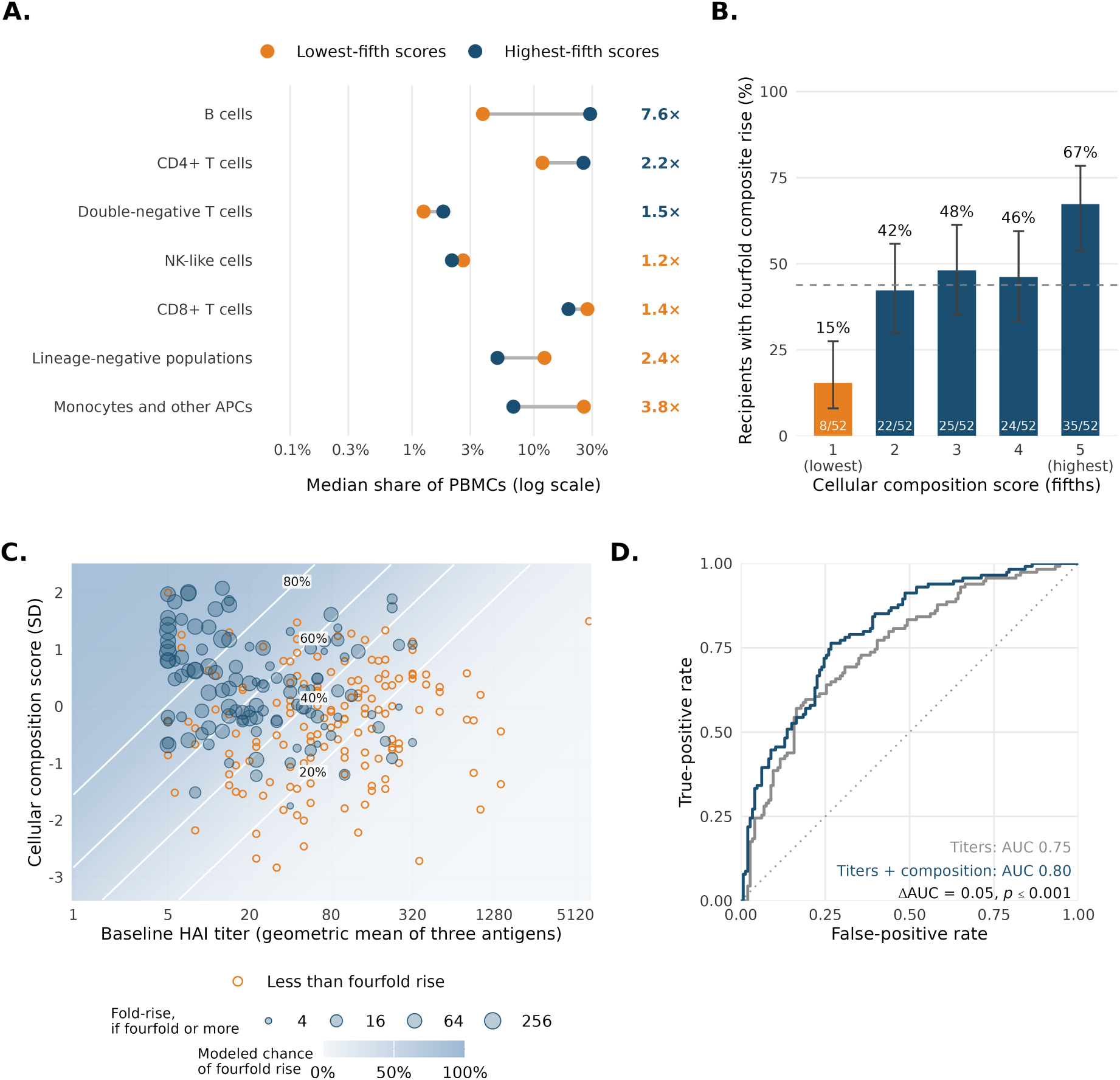
The cellular composition score in clinical terms. Scores come from models fitted without the recipient and were standardized within cross-validation fold. **A.** Median share of PBMCs in each cell lineage among recipients in the lowest (orange) and highest (navy) fifth of the score; numbers give the fold difference, colored by the group with the larger share. **B.** Percentage of recipients with a fourfold composite rise (geometric mean fold-rise across the 3 antigens of at least four) in each fifth of the score, with 95% Wilson intervals; the dashed line marks the overall rate. **C.** Baseline titer and score of each recipient. Open circles mark recipients below a fourfold rise; translucent blue circles mark a fourfold rise or more, with area increasing with the composite fold-rise, so overlapping points remain visible. Shading and contours show the modeled probability of a fourfold rise from a logistic regression on baseline titer and score. **D.** Receiver operating characteristic curves for a fourfold composite rise from out-of-fold predictions of the model with baseline titers alone (M1) and the model that adds immune-cell composition (M3); the *P* value comes from the null bootstrap that repeats the whole model fitting.

Adding immune-cell composition to baseline titers raised the AUC for a fourfold composite rise from 0.75 to 0.80 (difference, 0.050; 95% CI, 0.013–0.089; training-aware *P* ≤ .001, DeLong *P* = .009; Figure 4D), and adding it to titers and clinical factors raised the AUC from 0.78 to 0.81 (*P* = .034). Immune-cell composition was associated with time since transplantation (supplemental Section S3.4), but the score added information beyond it: adjusted for months since transplantation, vaccine dose, age and cohort, the odds ratio per SD was unchanged (2.1; 95% CI, 1.5–3.0; supplemental Figure S9A). The Spearman rank correlation between the score and the observed composite fold-rise was 0.39 (*P* ≤ .001), and the same gradient appeared for each antigen separately (supplemental Figures S10, S11 and S12; supplemental Table S4). When recipients in each of the lower three baseline-titer quartiles, where a fourfold rise is attainable, were split at that quartile’s median score and pooled, those above the median had a 2.7-fold larger geometric mean composite fold-rise than those at or below it (9.9 versus 3.7; supplemental Section S2.9, Figure S13 and Table S5). At the same score, responders differed from non-responders mainly in a lower baseline titer and a longer interval since transplantation (supplemental Figure S14). Responses overlapped across score levels, so the score separates groups more clearly than it separates individual recipients.

### 3.7 Children and adults shared the cellular profile

Pediatric and adult recipients differ in age, conditioning and immune recovery, so pooling them has to be justified. We evaluated the pooled model within each cohort and refitted the models separately in each cohort (Figure 5A). The pooled model reduced prediction error beyond baseline titers by 9.4% in children (*P* = .15) and 22.9% in adults (*P* < .001), so the pooled model improved prediction in both, more in adults. Refitting within each cohort on the shared 35-population atlas reduced prediction error by 6.2% in children (*n* = 147; *P* = .23) and 25.9% in adults (*n* = 113; *P* < .001), with wide intervals in each cohort; gains by baseline-titer quartile within each cohort are in supplemental Figure S15.

**Figure 5:**
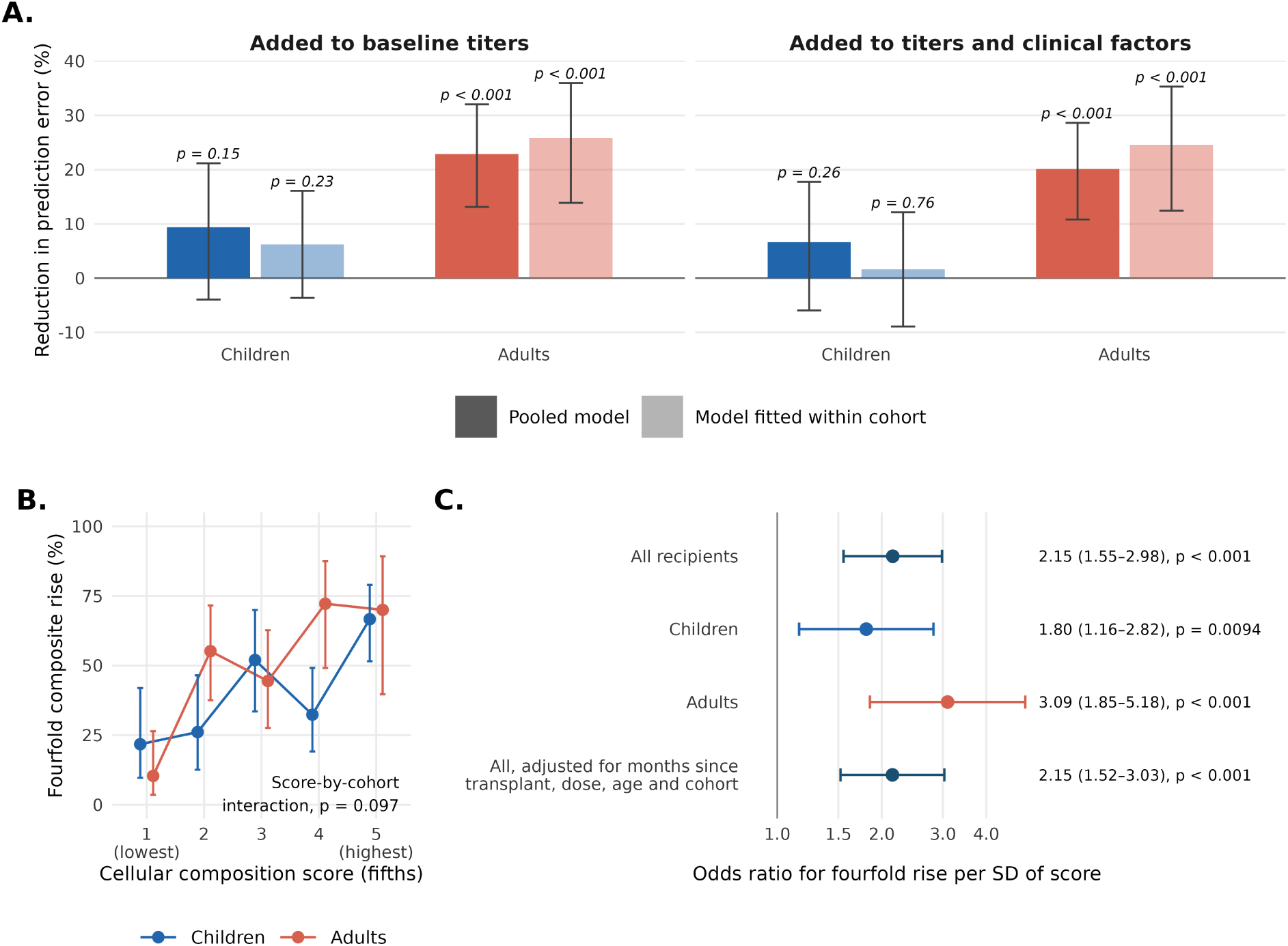
Children and adults share the cellular profile. **A.** Reduction in mean squared error of the predicted composite response when immune-cell composition is added to baseline titers (M1→M3) or to titers and clinical factors (M2→M4), for the pooled model evaluated within each cohort (dark bars) and for models refitted within each cohort (light bars), in 147 pediatric and 113 adult recipients. Whiskers are 95% recipient-bootstrap intervals. The cohort-specific models used the shared atlas built from both cohorts and are exploratory. **B.** Percentage of recipients with a fourfold composite rise by fifth of the score (fifths defined on all recipients), with 95% Wilson intervals; the interaction *P* value tests whether the association of the score with the response differs between cohorts. **C.** Odds ratio of a fourfold composite rise per SD of the score, adjusted for baseline titer, with 95% intervals and Wald *P* values.

The score pointed the same way in both cohorts, more strongly in adults. The odds ratio per SD was 1.8 in children and 3.1 in adults (interaction *P* = .10), the rank correlation with the observed fold-rise was 0.37 (*P* = .002) and 0.50 (*P* ≤ .001), and fourfold rises were least frequent in the lowest fifth in both (22% and 10%; Figure 5B and C).

The predictive populations were also shared. Of the 10 populations that the pooled model selected in at least half of its bootstrap refits, 8 had the same direction of association in children and in adults, and the three with the largest pooled coefficients, CD21-low B cells (cluster 4), CCR4^+^ central-memory CD8^+^ T cells (cluster 8) and activated double-negative T cells (cluster 32), kept their direction in both (supplemental Table S6, Figure S16). How often a population was selected differed more than its direction: the adult refits leaned on CD21-low B cells, exhausted CD8^+^ T cells and CCR4^+^ central-memory CD8^+^ T cells, the pediatric refits on activated double-negative T cells. A model trained in children predicted adult responses well, while the reverse did not hold (supplemental Section S2.10, Figure S17).

### 3.8 The score predicted response a season later in the same recipients

The 74 second-season episodes of recipients vaccinated in consecutive years (35 adults and 39 children) were never used to build an atlas or fit a model, so they test the first-season models in the same recipients one season later (Figure 6). Adding immune-cell composition to baseline titers reduced the prediction error of the composite fold-rise in these episodes by 39.2% (95% recipient-bootstrap CI, 28.3–48.9%), and the AUC for a fourfold composite rise rose from 0.65 with titers alone to 0.71 with composition (difference, 0.06; 95% CI, −0.03 to 0.14; DeLong *P* = .20; Figure 6A). Each SD of the first-season-standardized score was associated with 1.8-fold higher odds of a fourfold rise (95% CI, 0.9–3.5; *P* = .086).

**Figure 6:**
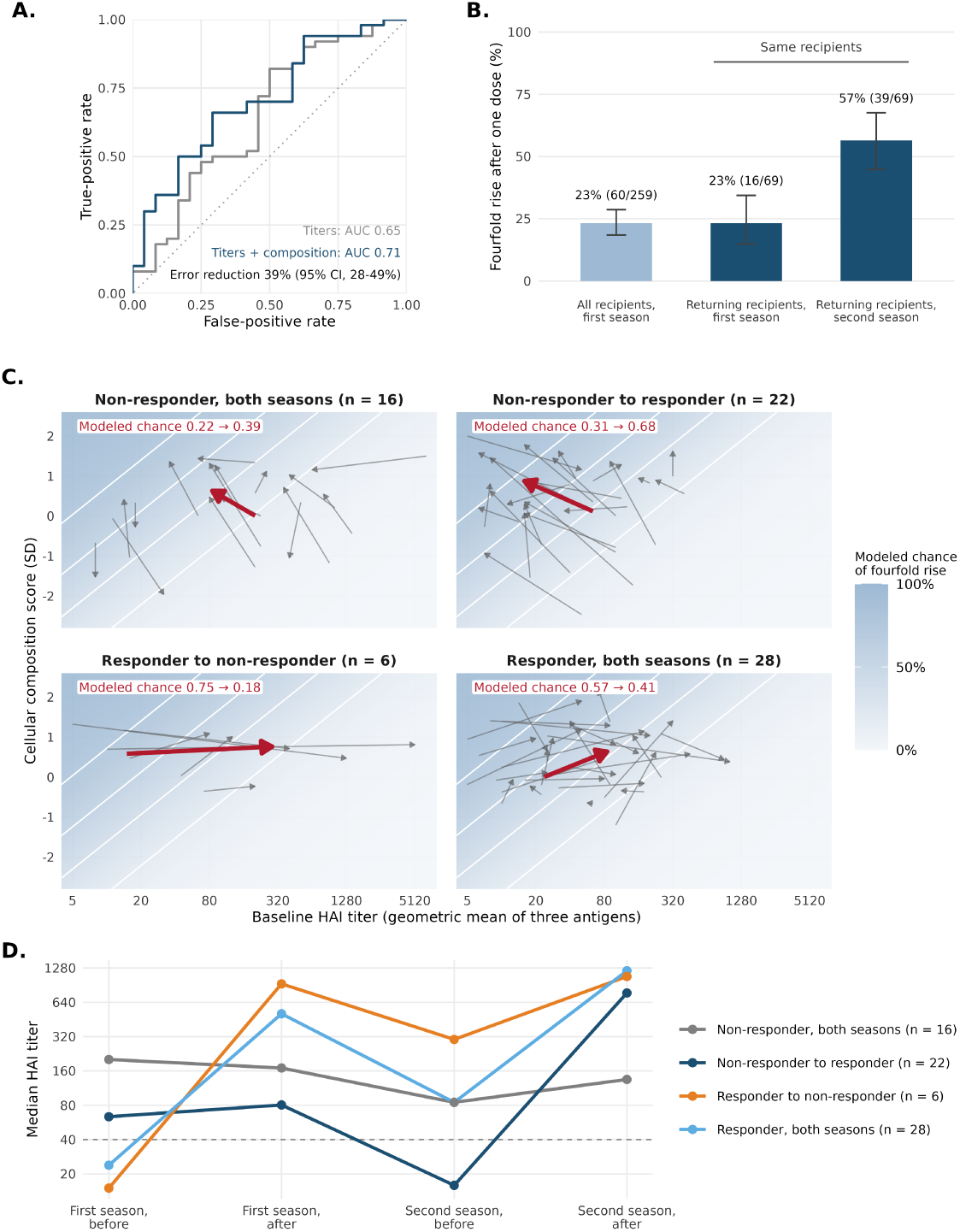
Validation in the second season of the same recipients. First-season models scored the 74 second-season episodes with recorded titers. These episodes were used neither to build an atlas nor to fit a model, and each was scored by the model that held out its recipient. A responder had at least a fourfold composite rise. **A.** Receiver operating characteristic curves for a fourfold composite rise in second-season episodes, from models with baseline titers alone or with immune-cell composition added. The error reduction is the bootstrap reduction in mean squared prediction error for the composite fold-rise. **B.** Percentage with a fourfold composite rise 4 weeks after the first dose, with 95% Wilson intervals, among all first-season recipients and among the 69 returning recipients with a one-dose titer in both seasons; the two dark bars are the same recipients. **C.** Each returning recipient’s move from the first to the second season on the response map of Figure 4C, by response transition. Shading and white contours give the modeled probability of a fourfold rise from the first-season logistic regression on baseline titer and score. Gray arrows are individual recipients, the red arrow joins the group’s median baseline titer and score from the first to the second season, and the red label gives the group’s median modeled probability of a fourfold rise in the two seasons (not a change in score or in the share of responders). Second-season scores are standardized with the first season’s fold mean and SD. **D.** Median HAI titer (geometric mean across antigens) before vaccination and 4 weeks after the second dose in each season, by response transition; the dashed line marks 1:40. Titers fell between seasons in every group. Because response is defined by the rise within a season, each group’s status follows the slope of its segments: recipients who did not respond in either season stayed nearly flat, those who became responders started the second season low and rose, and those who stopped responding started the second season high after a strong first-season response.

Responses were stronger in the second season. After the first dose alone, 57% of returning recipients (39 of 69) had a fourfold composite rise in the second season, against 23% (16 of 69) of the same recipients and 23% (60 of 259) of all recipients in the first season (Figure 6B), and 50 of 74 second-season episodes met the fourfold criterion after 2 doses. Of the 72 recipients with both seasons, 22 changed from non-responder to responder, 6 from responder to non-responder, 16 responded in neither season and 28 in both, where a responder had at least a fourfold composite rise. Cellular scores rose from the first to the second season in every transition group (median increase, 0.1 to 0.9 SD), consistent with continuing immune reconstitution. On the response map of Figure 4C, recipients who became responders moved toward lower baseline titers and higher scores, and their median modeled probability of a fourfold rise rose from 0.31 to 0.68, whereas recipients who stopped responding moved toward higher titers, and their median modeled probability fell from 0.75 to 0.18 (Figure 6C). The titer trajectories explain the second movement (Figure 6D). A strong first-season response left high antibody at the start of the second season, which leaves little room for a further fourfold rise, while recipients who had not responded started the second season with low titers. Changes in response status between seasons therefore reflect both the baseline titer and the cellular score, as the first-season model predicts.

### 3.9 Fewer populations and markers carried the predictive information

A simpler assay would be more practical clinically, prompting us to test whether fewer populations and markers preserved prediction accuracy. On the shared atlas, we ranked populations inside each training fold and refitted the model with only the top-ranked ones. Five populations recovered most of the gain of all 35 (17.3% versus 18.1% beyond titers; supplemental Figure S18); CD21-low B cells and CCR4^+^ central-memory CD8^+^ T cells ranked in the top 5 in every fold, and activated double-negative T cells and cycling monocytes in 4 of 5. An unweighted signature of the 3 most positive and 3 most negative populations, chosen inside each training fold, gave a gain of the same size as the full composition (18.6%; 95% CI, 9.1–27.0%; supplemental Section S2.6, Table S7), so a handful of populations, counted and combined without weights, carries most of the signal. Starting from the populations that drive the score, greedy forward selection,^40^ which adds at each step the marker that most improves identification of these populations in training recipients, found that 8 markers (CD25, CD45RO, CD8, CD27, CD38, CCR4, CD19 and CXCR3) reproduced the 30-marker cellular score computed from the same sampled cells of held-out recipients (*r* = 0.87; 95% CI, 0.80–0.93), although they assigned only 44% of cells to the correct population (supplemental Figure S19).

To reduce markers, for each training fold, we fixed the full-panel micro-centroids and their mapping to meta-clusters, including the number of clusters. Markers were prioritized by how strongly their expression differed across these meta-clusters. Using only the retained markers, we reassigned cells to the nearest micro-centroid and its meta-cluster. We then refitted M4 coefficients and penalty strength for each reduced panel; the prediction model itself was not held fixed.

Prediction error changed little across the tested panels. Test root mean squared error (RMSE) of the composite response was 1.395 with 30 markers, 1.403 with 15, 1.410 with eight and 1.380 with six, against 1.509 for baseline titers alone; fewer than six were not tested. Agreement with full-panel cell labels, by contrast, fell steadily, to 52.4% at 15 markers and 28.0% at six (Figure 7; supplemental Figure S20). Fewer markers therefore preserved the predictive information more faithfully than they preserved individual cell labels. Scores computed from 15 markers tracked 30-marker scores computed from the same cells closely (*r* = 0.91 for the composite score, after removing differences between cross-validation folds), and the score-group differences in observed responses were somewhat smaller but in the same direction (supplemental Section S2.6, Figure S21). The consensus 15-marker set is shown in Figure 7C and listed in supplemental Section S4.

**Figure 7:**
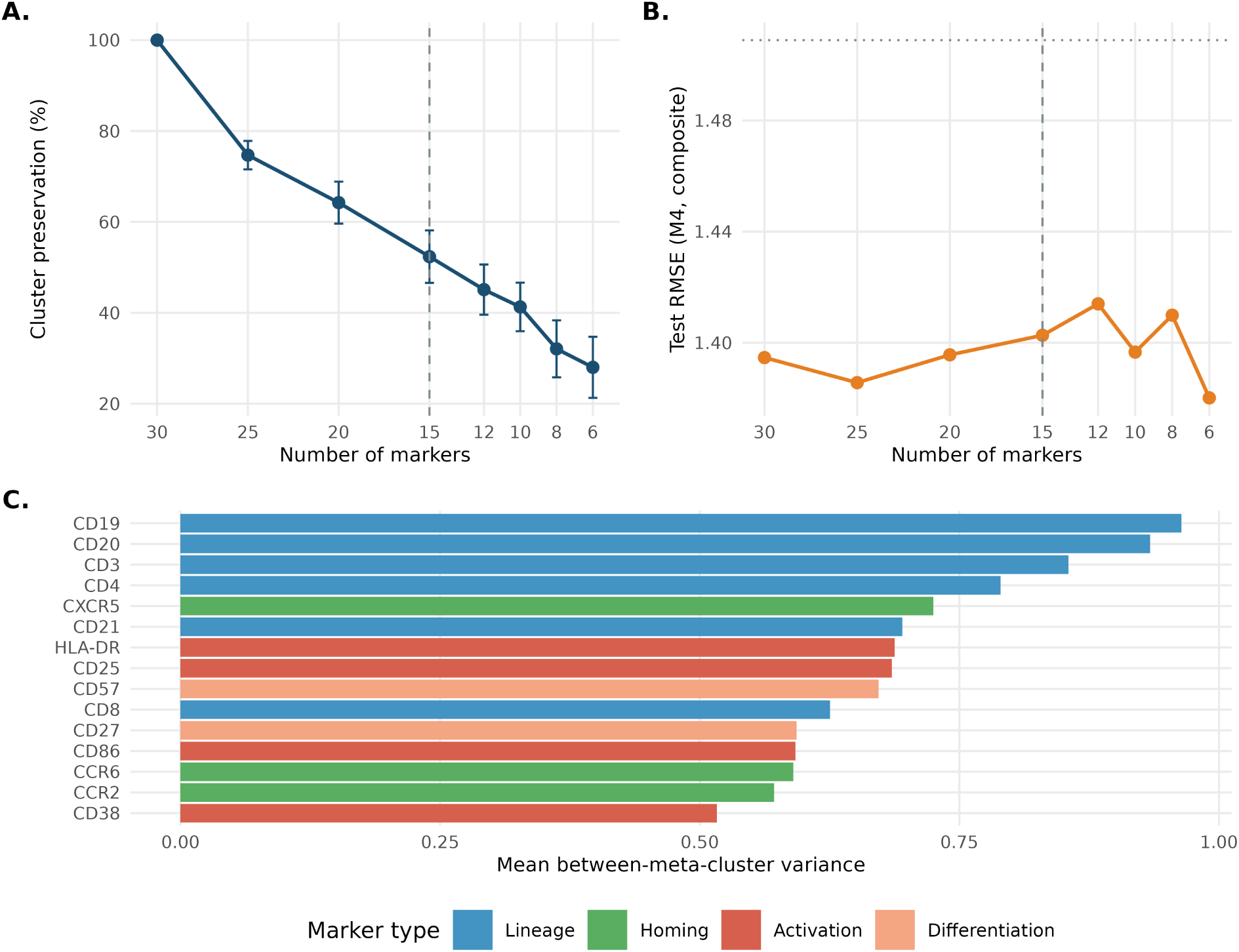
Prediction with fewer cytometry markers. The full-panel atlas was held fixed while cells were reassigned using smaller marker sets; prediction models were refitted for each set. **A.** Percentage of cells retaining their full-panel meta-cluster label, shown as the mean and standard deviation across 5 folds. **B.** Held-out root mean squared prediction error of the composite response for the model containing titers, clinical factors, and immune-cell composition (M4); lower values indicate better prediction. The dotted horizontal line shows the baseline-titer-only model. **C.** The 15 markers with the largest average between-meta-cluster variance across folds; individual fold-specific panels could differ. Prediction used up to 2,000 cells per episode at every marker budget, so the full-panel reference differs from the full-depth primary analysis. The vertical dashed line marks 15 markers.

### 3.10 The gain persisted in sensitivity analyses

The gain did not depend on the choices we made in building the models. Modeling the 3 antigens separately instead of their composite gave 11.2% beyond titers (supplemental Section S2.5). Running the whole pipeline on all 334 episodes with complete titers, from 262 recipients, with both seasons of a returning recipient kept in the same fold, gave 13.9% for the 3 antigens modeled jointly (*P* < .001; supplemental Section S3.7, Figures S22 and S23). Fixing the number of populations at any value between 5 and 42 instead of selecting it inside each fold gave between 10.4% and 16.4% (supplemental Section S1.2, Figure S24), so the result does not rest on how finely the cells were grouped. Allowing all pairwise interactions among titers and clinical factors did not change the picture: that model improved on titers alone by 3.1% without cellular composition and 15.7% with it (supplemental Table S8).

## 4 Discussion

In 260 allogeneic HCT recipients from 2 multicenter phase 2 randomized trials, the composition of peripheral blood before vaccination predicted the influenza antibody response beyond the pre-vaccination titer and the clinical factors a transplant physician already has. The practical outcome is the ability to identify the recipients least likely to respond to vaccination: in the lowest fifth of the cellular composition score, 15% reached a fourfold rise, compared with 51% of everyone else. The same score separated responders from non-responders in children and in adults, with the main populations pointing in the same direction, and it held when the first-season models were applied to the held-out second-season vaccination episodes of the same recipients.

Read as a biomarker, a low score marks a recipient whose B-cell compartment has not returned. A blood composition compatible with a response appears to be necessary but not sufficient: recipients in the highest fifth of the score still failed to reach a fourfold rise 33% of the time, while in the lowest fifth only 15% responded at all. That asymmetry is what makes the score useful for identifying likely non-responders, and it is also why the score should not be read as a prediction that an individual recipient will respond.

These results extend the pediatric association evidence of Amarin and colleagues^27^ by quantifying prediction beyond baseline titers and clinical factors in both cohorts, by turning the associations into one number per recipient, and by testing that number a season later in the same people.

Immune-cell composition reduced prediction error by 15.3% beyond titers alone and by 12.5% beyond titers plus clinical factors. A reduction in squared error is not a quantity clinicians work with, so we also put the signal on the antibody scale. At the same baseline titer, recipients at the 75th percentile of the score had post-vaccination titers about 2.2 times those at the 25th percentile (supplemental Table S9). For scale, 2 high-dose vaccinations produced adjusted geometric mean HAI titers about 1.2 to 2.1 times those after 2 standard doses in the adult trial,^10^ and about 1.9 to 3.2 times in children vaccinated six or more months after transplantation.^11^ The comparison is descriptive: vaccine dose was randomized, whereas the score summarizes observed differences between recipients. Against an assumed level of HAI assay noise, the gain from composition closes about 19% of the gap between the titers-only model and the smallest error the assay allows (supplemental Section S5, Table S10).

That difference matters most where recipients start low. A rise from 1:5 to 1:20 and a rise from 1:20 to 1:80 are both fourfold, but only the second leaves a titer in the range associated with protection in healthy adults,^35,36^ and the recipients who start at the detection limit are the ones in whom an extra doubling changes the end point. At high baseline titers the fold-rise is compressed by the ceiling of the assay and by the antibody already present, which is why we report the gain within baseline-titer quartiles rather than pooling across them.

What a transplant physician lacks before vaccinating is any way to tell, at the visit, which recipients are unlikely to seroconvert. Time since transplantation is the usual guide, and it is coarse: the score remained associated with the response after adjusting for it. A measurement taken from the same blood draw that identifies a fifth of recipients among whom fewer than one in five responds would, if it holds prospectively, be a reason to prioritize them for a second dose or a high-dose formulation, to revaccinate later, to give pre-exposure oseltamivir prophylaxis during the season,^41^ or to protect them by vaccinating household contacts.^42^ Each of those uses needs a prospective study with a defined threshold; what this study supports is that the measurement carries the information, not that a particular action follows from it. Models fitted separately in the high-dose and standard-dose arms ranked recipients similarly (Spearman *ρ* = 0.74; supplemental Figure S25), so these data do not identify who would gain from the high-dose formulation. The reduced panels point the same way: a flow-cytometry assay built on 15 or fewer markers would be the practical form of such a test.^43^

The populations the model used read as a picture of where a recipient sits in immune reconstitution. B cells and activated T cells carry the positive side: the largest coefficients belong to CD21-low B cells (cluster 4) and activated double-negative T cells (cluster 32), and across the score range the B-cell share rises nearly eightfold from the lowest to the highest fifth, in line with reports linking B-cell recovery to vaccine responses after allogeneic transplantation.^20^ CD21-low B cells are described as hyporesponsive in other settings,^44,45^ so their positive association here needs biological characterization; one possibility is that in this setting they mark an expanding B-cell compartment rather than a functional subset.

The negative side is carried by cycling monocytes, activated lineage-negative cells, memory CD8^+^ T cells and plasmablasts (clusters 17, 29, 8, 1 and 28). Because composition is relative, a high share of one population implies a lower share of others, and the lowest-score recipients had more monocytes and lineage-negative cells and few B cells. One reading, which these data support but do not establish, is a staged recovery: myeloid and lineage-negative cells predominate early after transplantation, and the lymphoid compartment, especially B cells, fills in later (supplemental Figure S26).^3,15^ Time since transplantation is consistent with that reading without accounting for it: the composition features explained half the variance in that interval, and the score remained associated with response after adjustment for it.

Our populations differ from those Amarin and colleagues reported,^27^ who found naive B cells, naive CD4^+^ T cells and circulating T follicular helper cells as univariate predictors in the pediatric cohort. Two differences are expected. Their analysis tested populations one at a time, which favors the populations positively associated with response, whereas a penalized model fitted to all populations at once keeps whichever combination predicts best and can drop a strong univariate predictor whose information another population already supplies. And a compositional model gives weight to populations that are high when the response is poor, which is how the monocyte and lineage-negative populations enter here. Circulating T follicular helper cells, which have forecast influenza responses in other studies,^46,47^ were not resolved as a separate population at the resolution of our atlas, and the CD4^+^ population richest in PD-1, ICOS and CXCR5 (cluster 25) contributed little to this model.

Prediction did not need the full panel. An unweighted signature of six populations predicted as well as all 35, and both the 15-marker and the smaller top-down panels reproduced most of the score. That a simpler count carries the same information argues that most of the signal lies in the broad composition, and it is what makes a flow-cytometry version of this measurement plausible. Reproducing a score is not the same as reproducing a cell-type assignment: at 15 markers only about half of cells kept their full-panel label.

The score was derived and tested in the same two trials. Nested cross-validation and the held-out second season guard against overfitting within this population, but there is no external cohort, and the transfer analysis shows that a model trained in one cohort does not automatically carry to the other. Although the trials were multi-center and randomized,^10,11^ the recipients are those two protocols enrolled, and results may not generalize to other transplant populations or vaccination schedules. With 260 recipients, within-cohort and quartile comparisons are imprecise.

The models used baseline titers, age, sex, vaccine dose and time since transplantation. They did not use transplant type, conditioning intensity, donor source, graft-versus-host disease or ongoing immunosuppression, each of which shapes immune recovery and may itself be reflected in the cellular composition the model reads. How well the score holds across that heterogeneity is an open question that a larger or prospective cohort would answer.

Mass cytometry is not widely available clinically, so translation would require adaptation to spectral flow cytometry.^48^ The panel lacks dedicated myeloid and NK-cell markers (CD14, CD16, CD11c, CD56) and CD45RA, so APC, monocyte and NK-like annotations were inferred by exclusion and EMRA T cells were defined without CD45RA; these labels should be validated with lineage-specific panels. We report prediction error and group contrasts, not performance against a decision threshold; what accuracy would be needed to guide the care of an individual recipient remains to be established prospectively. Finally, the outcome is the circulating antibody titer, so the analysis speaks only to humoral immunity; influenza vaccination also elicits cellular responses that may matter as much in this population^49^ and are not captured by the HAI assay.

The next step is to validate the score in an independent HCT cohort, ideally one with different conditioning and graft-versus-host disease prophylaxis. A reduced panel on spectral flow cytometry would need its own atlas, clustered and calibrated on those markers, and combining the cellular measurement with other baseline markers, such as gene expression or serum cytokines, may add what composition alone does not. Only a prospective test of whether the recipients the score flags benefit from a different vaccination strategy can turn a predictive measurement into a clinical decision.

## Data Availability

The de-identified CyTOF data and analysis code supporting the findings of this study are available from the corresponding author upon reasonable request. The analysis code is available at https://github.com/AsiaeeLab/hct-flu-cytof-prediction.

## Acknowledgments

This work was supported by the National Institute of Allergy and Infectious Diseases (U01 AI125135 and U01 AI132004), the National Center for Advancing Translational Sciences (UL1 TR002243) and the Tennessee Center for AIDS Research (NIH P30 AI110527). We thank the recipients and families who took part in the two trials, and the study teams at the participating transplant centers.

## Authorship Contributions

A.A. conceived the computational framework, performed the statistical and machine learning analyses, generated the figures, and drafted the manuscript. S.A.K. conceived the study, provided sample access and clinical context, and contributed to the interpretation of the results. J.D.S. performed the initial data preparation and exploratory bioinformatics and contributed to cell population annotation and biological interpretation. S.P. contributed to the predictive modeling and statistical analyses. A.J.S. advised on the statistical methodology and inference and critically reviewed the manuscript. J.D.C. contributed to the clinical interpretation and critically reviewed the manuscript. N.B.H. designed and led the two vaccine trials, provided the trial data and clinical context, and critically reviewed the manuscript. All authors reviewed and edited the manuscript and approved the final version.

## References

1. M Tomblyn, T Chiller, H Einsele, R Gress, K Sepkowitz, J Storek, J R Wingard, J-A H Young, and M J Boeckh. Guidelines for preventing infectious complications among hematopoietic cell transplantation recipients: a global perspective. Biol Blood Marrow Transplant, 15(10):1143–1238, 2009.

2. P Ljungman, C Cordonnier, H Einsele, J Englund, C M Machado, J Storek, and T Small. Vaccination of hematopoietic cell transplant recipients. Bone Marrow Transplant, 44(8):521–526, 2009.

3. J Storek, M A Dawson, B Storer, T Stevens-Ayers, D G Maloney, K A Marr, R P Witherspoon, W Bensinger, M E D Flowers, P Martin, R Storb, F R Appelbaum, and M Boeckh. Immune reconstitution after allogeneic marrow transplantation compared with blood stem cell transplantation. Blood, 97(11):3380–3389, 2001.

4. M J Memoli, R Athota, S Reed, L Czajkowski, T Bristol, K Proudfoot, R Hagey, J Voell, C Fiorentino, A Ademposi, S Shoham, and J K Taubenberger. The natural history of influenza infection in the severely immunocompromised vs nonimmunocompromised hosts. Clin Infect Dis, 58(2):214–224, 2014.

5. J Kmeid, J Vanichanan, D P Shah, F El Chaer, J Azzi, E J Ariza-Heredia, C Hosing, V Mulanovich, and R F Chemaly. Outcomes of influenza infections in hematopoietic cell transplant recipients: application of an immunodeficiency scoring index. Biol Blood Marrow Transplant, 22(3):542–548, 2016.

6. D Engelhard, B Mohty, R de la Camara, C Cordonnier, and P Ljungman. European guidelines for prevention and management of influenza in hematopoietic stem cell transplantation and leukemia patients: summary of ECIL-4 (2011). Transpl Infect Dis, 15(3):219–232, 2013.

7. L G Rubin, M J Levin, P Ljungman, E G Davies, R Avery, M Tomblyn, A Bousvaros, S Dhanireddy, L Sung, H Keyserling, and I Kang. Executive summary: 2013 IDSA clinical practice guideline for vaccination of the immunocompromised host. Clin Infect Dis, 58(3):309–318, 2014.

8. C Cordonnier, S Einarsdottir, S Cesaro, R Di Blasi, M Mikulska, C Rieger, H de Lavallade, G Gallo, T Lehrnbecher, D Engelhard, and P Ljungman. Vaccination of haemopoietic stem cell transplant recipients: guidelines of the 2017 European Conference on Infections in Leukaemia (ECIL 7). Lancet Infect Dis, 19(6):e200–e212, 2019.

9. N A Karras, M Weeres, W Sessions, X Xu, T Defor, J-A H Young, H Stefanski, C Brunstein, S Cooley, J S Miller, B R Blazar, J E Wagner, and M R Verneris. A randomized trial of one versus two doses of influenza vaccine after allogeneic transplantation. Biol Blood Marrow Transplant, 19(1):109–116, 2013.

10. Lora D Thomas, Einas Batarseh, Lubna Hamdan, Zaid Haddadin, Daniel Dulek, Spyros Kalams, Laura S Stewart, Anna L Stahl, Herdi Rahman, Justin Z Amarin, Haya Hayek, Michael Ison, Edgar T Overton, Steven A Pergam, Andrew J Spieker, Natasha B Halasa, and the Adult HCT Flu Study. Comparison of two high-dose versus two standard-dose influenza vaccines in adult allogeneic hematopoietic cell transplant recipients. Clin Infect Dis, 77(12):1723–1732, 2023.

11. Jennifer E Schuster, Lubna Hamdan, Daniel E Dulek, Carrie L Kitko, Einas Batarseh, Zaid Haddadin, Laura S Stewart, Anna Stahl, Molly Potter, Herdi Rahman, Spyros A Kalams, Claire E Bocchini, Elizabeth A Moulton, Susan E Coffin, Monica I Ardura, Rachel L Wattier, Gabriela Maron, Michael Grimley, Grant Paulsen, Christopher J Harrison, Jason L Freedman, Paul A Carpenter, Janet A Englund, Flor M Munoz, Lara Danziger-Isakov, Andrew J Spieker, Natasha B Halasa, and for the Pediatric HCT Flu Study. The durability of antibody responses of two doses of high-dose relative to two doses of standard-dose inactivated influenza vaccine in pediatric hematopoietic cell transplant recipients: A multi-center randomized controlled trial. Clin Infect Dis, 78(1):217–226, 2024.

12. M Janssen, A Bruns, J Kuball, R Raijmakers, and D van Baarle. Vaccine responses in adult hematopoietic stem cell transplant recipients: a comprehensive review. Cancers (Basel), 13(23):6140, 2021.

13. P A Carpenter and J A Englund. Commentary: is immune recovery-based post-transplantation vaccination in children better than time-based revaccination? Transplant Cell Ther, 27(4):281–283, 2021.

14. A S Haynes, D J Curtis, K Campbell, R H Giller, R R Quinones, M R Verneris, and M J Abzug. An immune recovery-based revaccination protocol for pediatric hematopoietic stem cell transplant recipients: revaccination outcomes following pediatric HSCT. Transplant Cell Ther, 27(4):317–326, 2021.

15. J Ogonek, M Kralj Juric, S Ghimire, P R Varanasi, E Holler, H Greinix, and E Weissinger. Immune reconstitution after allogeneic hematopoietic stem cell transplantation. Front Immunol, 7, 2016.

16. G Avetisyan, J Aschan, M Hassan, and P Ljungman. Evaluation of immune responses to seasonal influenza vaccination in healthy volunteers and in patients after stem cell transplantation. Transplantation, 86(2):257–263, 2008.

17. C M Machado, M R A Cardoso, I F da Rocha, L S Boas, F L Dulley, and C S Pannuti. The benefit of influenza vaccination after bone marrow transplantation. Bone Marrow Transplant, 36(10):897–900, 2005.

18. B Mohty, M Bel, M Vukicevic, M Nagy, E Levrat, S Meier, S Grillet, C Combescure, L Kaiser, Y Chalandon, J Passweg, C A Siegrist, and E Roosnek. Graft-versus-host disease is the major determinant of humoral responses to the AS03-adjuvanted influenza A/09/H1N1 vaccine in allogeneic hematopoietic stem cell transplant recipients. Haematologica, 96(6):896–904, 2011.

19. J Linnik, M Syedbasha, H M Kaltenbach, D Vogt, Y Hollenstein, L Kaufmann, N Cantoni, S Ruosch-Girsberger, A M S Müller, U Schanz, T Pabst, G Stüssi, M Weisser, J Halter, J Stelling, and A Egli. Association of host factors with antibody response to seasonal influenza vaccination in allogeneic hematopoietic stem cell transplant patients. J Infect Dis, 225(8):1482–1493, 2022.

20. M J M Janssen, A H W Bruns, F M Verduyn Lunel, R A P Raijmakers, R J de Weijer, N M Nanlohy, G P Smits, D van Baarle, and J Kuball. Predictive factors for vaccine failure to guide vaccination in allogeneic hematopoietic stem cell transplant recipients. Bone Marrow Transplant, 56(12):2922–2928, 2021.

21. H I Nakaya, J Wrammert, E K Lee, L Racioppi, S Marie-Kunze, W N Haining, A R Means, S P Kasturi, N Khan, G M Li, M McCausland, V Kanchan, K E Kokko, S Li, R Elbein, A K Mehta, A Aderem, K Subbarao, R Ahmed, and B Pulendran. Systems biology of vaccination for seasonal influenza in humans. Nat Immunol, 12(8):786–795, 2011.

22. J S Tsang, P L Schwartzberg, Y Kotliarov, A Biancotto, Z Xie, R N Germain, E Wang, M J Olnes, M Narayanan, H Golding, S Moir, H B Dickler, S Perl, F Cheung, G Obermoser, D Chaussabel, K Palucka, J Chen, J C Fuchs, J Ho, S Khurana, L R King, M Langweiler, H Liu, J Manischewitz, Z Pos, J G Posada, P Schum, R Shi, J Valdez, W Wang, H Zhou, D L Kastner, F M Marincola, J P McCoy, G Trinchieri, and N S Young. Global analyses of human immune variation reveal baseline predictors of postvaccination responses. Cell, 157(2):499–513, 2014.

23. M Cortese, A C Sherman, N G Rouphael, and B Pulendran. Systems biological analysis of immune response to influenza vaccination. Cold Spring Harb Perspect Med, 11(6):a038596, 2021.

24. M Cortese, T Hagan, N Rouphael, S Y Wu, X Xie, D Kazmin, F Wimmers, S Gupta, R van der Most, M Coccia, P S Aranuchalam, H I Nakaya, Y Wang, E Coyle, S Horiuchi, H Wu, M Bower, A Mehta, C Gunthel, S E Bosinger, Y Kotliarov, F Cheung, P L Schwartzberg, R N Germain, J Tsang, S Li, R Albrecht, H Ueno, S Subramaniam, M J Mulligan, S Khurana, H Golding, and B Pulendran. System vaccinology analysis of predictors and mechanisms of antibody response durability to multiple vaccines in humans. Nat Immunol, 26(1):116–130, 2025.

25. P S Arunachalam, M K D Scott, T Hagan, C Li, Y Feng, F Wimmers, L Grigoryan, M Trisal, V V Edara, L Lai, S E Chang, A Feng, S Dhingra, M Shah, A S Lee, S Chinthrajah, S B Sindher, V Mallajosyula, F Gao, N Sigal, S Kowli, S Gupta, K Pellegrini, G Tharp, S Maysel-Auslender, S Hamilton, H Aoued, K Hrusovsky, M Roskey, S E Bosinger, H T Maecker, S D Boyd, M M Davis, P J Utz, M S Suthar, P Khatri, K C Nadeau, and B Pulendran. Systems vaccinology of the BNT162b2 mRNA vaccine in humans. Nature, 596(7872):410–416, 2021.

26. S Hao, I Tomic, B B Lindsey, Y J Jagne, K Hoschler, A Meijer, J M Carreño Quiroz, P Meade, K Sano, C Peno, A G Costa-Martins, D Bogaert, B Kampmann, H Nakaya, F Krammer, T I de Silva, and A Tomic. Integrative mapping of preexisting influenza immune landscapes predicts vaccine response. J Clin Invest, 135(18):e189300, 2025.

27. J Z Amarin, D E Dulek, J Simmons, H Hayek, J D Chappell, C H Nochowicz, C L Kitko, J E Schuster, F M Muñoz, C E Bocchini, E A Moulton, S E Coffin, J L Freedman, M I Ardura, R L Wattier, G Maron, M Grimley, G Paulsen, L Danziger-Isakov, P A Carpenter, J A Englund, N B Halasa, A J Spieker, and S A Kalams. Immunophenotypic predictors of influenza vaccine immunogenicity in pediatric hematopoietic cell transplant recipients. Blood Adv, 8(8):1880–1892, 2024.

28. N B Halasa, B N Savani, I Asokan, A Kassim, R Simons, C Summers, J Bourgeois, C Clifton, L A Vaughan, C Lucid, L Wang, C Fonnesbeck, and M Jagasia. Randomized double-blind study of the safety and immunogenicity of standard-dose trivalent inactivated influenza vaccine versus high-dose trivalent inactivated influenza vaccine in adult hematopoietic stem cell transplantation patients. Biol Blood Marrow Transplant, 22(3):528–535, 2016.

29. C A DiazGranados, A J Dunning, M Kimmel, D Kirby, J Treanor, A Collins, R Pollak, J Christoff, J Earl, V Landolfi, E Martin, S Gurunathan, R Nathan, D P Greenberg, N G Tornieporth, M D Decker, and H K Talbot. Efficacy of high-dose versus standard-dose influenza vaccine in older adults. N Engl J Med, 371(7):635–645, 2014.

30. H S Izurieta, N Thadani, D K Shay, Y Lu, A Maurer, I M Foppa, R Franks, D Pratt, R A Forshee, T MaCurdy, C Worrall, A E Howery, and J Kelman. Comparative effectiveness of high-dose versus standard-dose influenza vaccines in US residents aged 65 years and older from 2012 to 2013 using Medicare data: a retrospective cohort analysis. Lancet Infect Dis, 15(3):293–300, 2015.

31. M Rivera-Izquierdo, A Verdejo-Iáñez, A Morales-Portillo, M González-Alcaide, A J Láinez-Ramos-Bossini, V Martínez-Ruiz, N F Fernández-Martínez, L M Martín-delosReyes, E Jiménez-Mejías, I Guerrero-Fernández de Alba, M D C Valero-Ubierna, P Lardelli-Claret, J A Schoenenberger-Arnaiz, and J J Jiménez-Moleón. High-dose versus standard-dose influenza vaccine for immunocompromised patients: a systematic review and meta-analysis of randomised clinical trials. J Infect, 91(2):106538, 2025.

32. S C Bendall, E F Simonds, P Qiu, E D Amir, P O Krutzik, R Finck, R V Bruggner, R Melamed, A Trejo, O I Ornatsky, R S Balderas, S K Plevritis, K Sachs, D Pe’er, S D Tanner, and G P Nolan. Single-cell mass cytometry of differential immune and drug responses across a human hematopoietic continuum. Science, 332(6030):687–696, 2011.

33. M H Spitzer and G P Nolan. Mass cytometry: single cells, many features. Cell, 165(4):780–791, 2016.

34. I Korsunsky, N Millard, J Fan, K Slowikowski, F Zhang, K Wei, Y Baglaenko, M Brenner, P Loh, and S Raychaudhuri. Fast, sensitive and accurate integration of single-cell data with Harmony. Nature Methods, 16(12):1289–1296, 2019.

35. D Hobson, R L Curry, A S Beare, and A Ward-Gardner. The role of serum haemagglutination-inhibiting antibody in protection against challenge infection with influenza A2 and B viruses. J Hyg (Lond), 70(4):767–777, 1972.

36. S Black, U Nicolay, T Vesikari, M Knuf, G Del Giudice, G Della Cioppa, T Tsai, R Clemens, and R Rappuoli. Hemagglutination inhibition antibody titers as a correlate of protection for inactivated influenza vaccines in children. Pediatr Infect Dis J, 30(12):1081–1085, 2011.

37. J Friedman, T Hastie, and R Tibshirani. Regularization paths for generalized linear models via coordinate descent. J Stat Softw, 33(1):1–22, 2010.

38. V A Traag, L Waltman, and N J van Eck. From Louvain to Leiden: guaranteeing well-connected communities. Sci Rep, 9(1):5233, 2019.

39. HIPC-CHI Signatures Project Team, HIPC-I Consortium, S Avey, F Cheung, D Fermin, J Frelinger, R Gaujoux, R Gottardo, P Khatri, S H Kleinstein, Y Kotliarov, H Meng, R Sauteraud, S S Shen-Orr, J S Tsang, F Vallania, E Anguiano, J Baisch, N Baldwin, R B Belshe, T P Blevins, D Chaussabel, M M Davis, E Fikrig, D E Grill, D A Hafler, E Henrich, S R Joshi, S M Kaech, R B Kennedy, S Mohanty, R R Montgomery, A L Oberg, G Obermoser, I G Ovsyannikova, A K Palucka, V Pascual, G Poland, B Pulendran, E Reinherz, A C Shaw, B Siconolfi, K D Stuart, S Tsang, I Ueda, J Wilson, and H J Zapata. Multicohort analysis reveals baseline transcriptional predictors of influenza vaccination responses. Science Immunology, 2(14):eaal4656, 2017.

40. I Guyon and A Elisseeff. An introduction to variable and feature selection. Journal of Machine Learning Research, 3:1157–1182, 2003.

41. T M Uyeki, H H Bernstein, J S Bradley, J A Englund, T M File, Jr, A M Fry, S Gravenstein, F G Hayden, S A Harper, J M Hirshon, M G Ison, B L Johnston, S L Knight, A McGeer, L E Riley, C R Wolfe, P E Alexander, and A T Pavia. Clinical practice guidelines by the Infectious Diseases Society of America: 2018 update on diagnosis, treatment, chemoprophylaxis, and institutional outbreak management of seasonal influenza. Clinical Infectious Diseases, 68(6):e1–e47, 2019.

42. A M Zorger, C Hirsch, M Baumann, M Feldmann, P J Bröckelmann, S Mellinghoff, I Monsef, N Skoetz, and N Kreuzberger. Vaccines for preventing infections in adults with haematological malignancies. Cochrane Database Syst Rev, 2025(5), 2025.

43. H T Maecker, J P McCoy, and R Nussenblatt. Standardizing immunophenotyping for the Human Immunology Project. Nat Rev Immunol, 12(3):191–200, 2012.

44. I Isnardi, Y-S Ng, L Menard, G Meyers, D Saadoun, I Srdanovic, J Samuels, J Berman, J H Buckner, C Cunningham-Rundles, and E Meffre. Complement receptor 2/CD21^−^ human naive B cells contain mostly autoreactive unresponsive clones. Blood, 115(24):5026–5036, 2010.

45. M Rakhmanov, B Keller, S Gutenberger, C Foerster, M Hoenig, G Driessen, M van der Burg, J J M van Dongen, E Wiech, M Visentini, I Quinti, A Prasse, N Voelxen, U Salzer, S Goldacker, P Fisch, H Eibel, K Schwarz, H-H Peter, and K Warnatz. Circulating CD21*^low^* B cells in common variable immunodeficiency resemble tissue homing, innate-like B cells. Proc Natl Acad Sci U S A, 106(32):13451–13456, 2009.

46. M A Pilkinton, K J Nicholas, C M Warren, R M Smith, S M Yoder, H K Talbot, and S A Kalams. Greater activation of peripheral T follicular helper cells following high dose influenza vaccine in older adults forecasts seroconversion. Vaccine, 35(2):329–336, 2017.

47. J Currenti, J Simmons, J Oakes, S Gaudieri, C M Warren, R Gangula, E Alves, R Ram, S Leary, J D Armitage, R M Smith, A Chopra, N B Halasa, M A Pilkinton, and S A Kalams. Tracking of activated cTfh cells following sequential influenza vaccinations reveals transcriptional profile of clonotypes driving a vaccine-induced immune response. Front Immunol, 14:1133781, 2023.

48. L M Park, J Lannigan, and M C Jaimes. OMIP-069: forty-color full spectrum flow cytometry panel for deep immunophenotyping of major cell subsets in human peripheral blood. Cytometry A, 97(10):1044–1051, 2020.

49. L Rodríguez-Guilarte, C Méndez, A Reyes, M Rios, F Román, D Moreno-Tapia, A Cabrera, D B Rivera, and A M Kalergis. Influenza vaccines promote humoral and cellular immune responses: a randomized, double-blind, phase 3 trial. Nat Commun, 17(1):407, 2025.

50. J Aitchison. The statistical analysis of compositional data. J R Stat Soc Series B Stat Methodol, 44(2):139–177, 1982.

51. A Colin Cameron, Jonah B Gelbach, and Douglas L Miller. Bootstrap-based improvements for inference with clustered errors. Rev Econ Stat, 90(3):414–427, 2008.

52. A C Davison and D V Hinkley. Bootstrap Methods and Their Application. Cambridge University Press, 1997.

53. A Chatterjee and S N Lahiri. Bootstrapping lasso estimators. J Am Stat Assoc, 106(494):608–625, 2011.

54. Yoshua Bengio and Yves Grandvalet. No unbiased estimator of the variance of *k*-fold cross-validation. J Mach Learn Res, 5:1089–1105, 2004.

55. Lara Lusa, Lisa M. McShane, Michael D. Radmacher, Joanna H. Shih, George W. Wright, and Richard Simon. Appropriateness of some resampling-based inference procedures for assessing performance of prognostic classifiers derived from microarray data. Stat Med, 26:1102–1113, 2007.

56. Sture Holm. A simple sequentially rejective multiple test procedure. Scand J Stat, 6(2):65–70, 1979.

57. H Bahakel, A J Spieker, H Hayek, J E Schuster, L Hamdan, D E Dulek, C L Kitko, T Stopczynski, E Batarseh, Z Haddadin, L S Stewart, A Stahl, M Potter, H Rahman, J Amarin, S A Kalams, C E Bocchini, E A Moulton, S E Coffin, M I Ardura, R L Wattier, G Maron, M Grimley, G Paulsen, C J Harrison, J Freedman, P A Carpenter, J A Englund, F M Munoz, L Danziger-Isakov, N Halasa, R Goyal, R McHenry, M Bender, S Barto, M Russo, L Shoemaker, K Truong, C Dvorak, K J Allison, S Naik, C Williams, S Blum, K Lacombe, and H Smith. Immunogenicity and reactogenicity of high-or standard-dose influenza vaccine in a second consecutive influenza season. J Infect Dis, 231(1):e123–e131, 2025.

