## Supplemental Materials for "Deep immune cell phenotyping predicts influenza vaccine immunogenicity in HCT recipients"

#### S1 Atlas construction and prediction methods

##### S1.1 Atlas construction and cellular features

The atlas converts single-cell marker measurements into one immune-composition profile per vaccination episode. Its construction separates fine groups of similar cells (micro-clusters) from the larger groups used as predictive features (meta-clusters, or cell populations).

**Micro-clustering.** One million cells sampled from outer-training-fold recipients were grouped into 5,000 micro-clusters by $k$-means. Their centers summarize the 30-marker profiles used for subsequent cell assignment.

**Meta-clustering.** Leiden community detection grouped a 30-nearest-neighbor graph of the micro-cluster centers. The 19 candidate resolutions were $r \in \{0.5, 0.75, \ldots, 5.0\}$. Each resolution produces a candidate grouping into meta-clusters, and therefore a candidate number of cell populations; an inner cross-validation on the composite outcome, run separately inside each outer training fold, selected the grouping used for prediction (supplemental Section S1.2).

**Cell assignment.** Each training and test cell was assigned to its nearest micro-cluster center and inherited that micro-cluster's meta-cluster label. Counts were summarized separately for each vaccination episode $(i, t)$; second-season episodes were assigned to the atlas of the fold holding out their recipient but never used to build it.

**Cellular features.** Let $n_{itj}$ be the number of cells assigned to meta-cluster $j$ in episode $(i, t)$. Adding 0.5 to each count handles populations with no sampled cells. The resulting fractions and centered log-ratio (CLR) features are

$$p_{itj} = \frac{n_{itj} + 0.5}{\sum_{\ell=1}^{k} (n_{it\ell} + 0.5)}, \qquad z_{itj} = \log(p_{itj}) - \frac{1}{k} \sum_{\ell=1}^{k} \log(p_{it\ell}). \tag{1}$$

Each feature expresses a population's abundance relative to the geometric mean abundance of all populations in that episode. This accounts for the relative nature of cell fractions.[50]

**Prediction.** The primary outcome is the composite log fold-rise, the mean of the three antigen log fold-rises. Single-outcome elastic nets predicted it with the `gaussian` family in `glmnet` and $\alpha = 0.9$. M3 combined baseline titers with cellular features; M4 also included clinical factors. Penalty strength was selected within training data using `lambda.min`. Multitask elastic nets that predict the three antigen log fold-rises jointly (the `mgaussian` family, same $\alpha$ and penalty rule) are reported as a supplementary analysis in Section S2.5.

#### S1.2 Selection of clustering resolution

The number of cell populations is not fixed in advance. It follows from the Leiden resolution, and the resolution was chosen inside each outer training fold by how well the resulting populations predicted the composite outcome in inner validation data. No held-out recipient took part in that choice. For resolution $r$, the criterion averaged three inner validation folds:

$$\mathrm{CV}(r) = \frac{1}{3}\sum_{\ell=1}^{3} \frac{\mathrm{MSE}_{\mathrm{M4},\ell}(r)}{\mathrm{MSE}_{\text{clinical OLS},\ell}}, \tag{2}$$

where both terms are composite mean squared errors in inner fold $\ell$: the numerator from the composite M4 elastic net fitted at resolution $r$, the denominator from an unpenalized least-squares model on baseline titers and clinical factors fitted in the same training subset. The ratio puts folds of unequal prediction difficulty on one scale. The resolution minimizing $\mathrm{CV}(r)$ was selected. Across the five outer folds this gave 42, 35, 42, 37 and 25 populations (resolutions 5.0, 4.5, 4.75, 4.5 and 3.25; supplemental Figure S1).

The gain does not hinge on this choice. Fixing the resolution, and with it the number of populations, at each value of the 19-point grid instead of selecting it gave held-out composite gains beyond baseline titers between 10.4% (5 populations) and 16.4% (35 populations), for example 11.4% at 10 populations, 11.3% at 18, 10.9% at 25, 16.4% at 35 and 14.8% at 42, with overlapping bootstrap intervals throughout (supplemental Figure S24). Predictive information therefore survives at every level of granularity we examined, and the result does not depend on how finely the cells are grouped.

#### S1.3 Baseline titers in a fold-rise model

Baseline antibody levels help distinguish the information supplied by immune-cell composition from differences already evident in serology. Although baseline titer also appears in the outcome, including it as a predictor gives a well-defined conditional regression:

$$\mathbb{E}[y_{itv} \mid \log(\mathrm{titer}_{itv,\mathrm{pre}}),\ \text{other predictors}]\,.$$

Because log fold-rise equals post-vaccination log titer minus baseline log titer, its baseline coefficient is one less than the corresponding coefficient in a model for post-vaccination log titer. Including baseline therefore changes the interpretation of that coefficient without creating algebraic circularity. The cellular coefficients describe associations with fold-rise after accounting for baseline antibody levels and, in M4, clinical factors.

#### S1.4 Uncertainty in model comparisons

Uncertainty in model comparisons was assessed by resampling recipients. The primary analysis has one episode per recipient; where an analysis uses more than one episode or the three antigens separately, a recipient's episodes and antigen measurements were resampled together. This preserves within-recipient dependence while comparing models on the same observations.[51,52] The bootstrap summarizes paired prediction errors. Coefficient selection frequencies describe the stability of fitted features; they are not coefficient significance tests. Standard bootstrap inference for selected lasso coefficients requires additional assumptions or modification.[53,54]

Sensitivity analyses examined 2,000 versus 5,000 micro-clusters, the three antigens modeled separately instead of the composite, all episodes of every recipient, a number of populations fixed rather than selected, and clinical baselines with pairwise interactions.

### S2 Cellular scores and predictor interpretation

#### S2.1 Cellular scores and observed antibody responses

##### S2.1.1 Score definition and grouping

The cellular composition score summarizes how pre-vaccination immune-cell composition contributes to predicted antibody fold-rise. The primary score is the composite one, built from the composite model. Per-antigen scores follow the same definition with that antigen's coefficients and are reported with the per-antigen models (Section S2.5). For outcome $v$, the composite or one of the three antigens, and held-out episode $(i, t)$, the score was

$$s_{itv} = \sum_{j=1}^{k_f} z_{itj} \widehat{\beta}_{jv}^{(-f)}, \tag{3}$$

where $f$ is the outer fold containing recipient $i$, $k_f$ is the number of populations in that fold's training atlas, and $z_{itj}$ is the episode's CLR feature for population $j$ in that atlas. The superscript $(-f)$ denotes an M3 coefficient fitted without any recipient in fold $f$; where a recipient contributed a second-season episode, both episodes were held out together. The score excludes the intercept and the baseline-titer terms, although those predictors were included when the coefficients were fitted. M3 contains no age, sex, dose or transplant-timing terms. The observed outcome was not residualized.

For each episode, we summarized baseline antibody levels across all three antigens by their geometric mean,

$$b_{it} = \exp\left\{ \frac{1}{3} \sum_{v=1}^{3} \log(\text{titer}_{itv,\text{pre}}) \right\}, \tag{4}$$

where $v$ indexes H1N1, H3N2 and B/Victoria. The sets $Q_1, \ldots, Q_4$ partition the 260 first-season recipients into quartiles of $\log b_{it}$ (67, 64, 66 and 63 recipients), matching Table 3. Thus, these are common baseline-titer groups across antigens, rather than quartiles of one antigen's baseline titer alone. For each quartile $q$ and outcome $v$, the score threshold was

$$c_{qv} = \text{median}\{s_{itv} : (i,t) \in Q_q\}. \tag{5}$$

The higher- and lower-score groups were, respectively,

$$\begin{aligned} H_{qv} &= \{(i,t) \in Q_q : s_{itv} > c_{qv}\}, \\ L_{qv} &= \{(i,t) \in Q_q : s_{itv} \leq c_{qv}\}. \end{aligned} \tag{6}$$

Each outcome has its own cellular coefficients and four median thresholds, so group membership can differ between the composite and the single antigens. The Q1 split, for example, assigns 33 recipients above the median to the higher-score group and 34 at or below it to the lower-score group. The median split provides a descriptive comparison of a continuous predictor; the cutoff was not chosen to maximize observed response separation and is not a validated clinical threshold. The pooled Q1–Q3 groups are $\bigcup_{q=1}^{3} H_{qv}$ and $\bigcup_{q=1}^{3} L_{qv}$: they retain the within-quartile assignments and reuse the same recipients displayed in the individual quartiles.

##### S2.1.2 Observed response summaries and intervals

Geometric mean fold-rise was $\exp(\overline{y})$ and a fourfold response was $y \geq \log 4$. Q1–Q3 pooling retained each quartile's score split. Confidence intervals resampled the 260 recipients ($B = 2{,}000$) while holding the fitted scores, quartiles and score cutoffs fixed, so they describe sampling variation conditional on the fitted scores rather than full model-training uncertainty. The per-antigen versions of the same comparison are in supplemental Table S4.

For a fixed outcome $v$ and quartile/score group $G$, define the observed fourfold-response indicator $R_{itv} = \mathbf{1}\{y_{itv} \geq \log 4\}$. The two panels in supplemental Figure S12 summarize the same recipients:

$$\text{GMFR}_{Gv} = \exp\left(\frac{\sum_{(i,t)\in G} y_{itv}}{|G|}\right), \qquad P_{Gv} = 100\frac{\sum_{(i,t)\in G} R_{itv}}{|G|}. \tag{7}$$

For bootstrap sample $b$, let $W_i^{(b)}$ be the number of times recipient $i$ is drawn in 260 draws with replacement from the 260 recipients. The resampled response percentage is

$$P_{Gv}^{(b)} = 100\frac{\sum_{(i,t)\in G} W_i^{(b)} R_{itv}}{\sum_{(i,t)\in G} W_i^{(b)}}. \tag{8}$$

Panel A instead exponentiates the weighted mean of $y_{itv}$ using these same weights. The intervals are the 2.5th and 97.5th percentiles across 2,000 replicates. Sampling is across all recipients, not separately within each quartile or score group, so the original group sizes are not fixed in the bootstrap. The observed antibody ratios, fitted scores, baseline quartiles and higher/lower labels are unchanged; only recipient multiplicities vary.

###### S2.1.3 Sensitivity to the score-group definition

The within-quartile split keeps approximately half of each baseline quartile in each score group. Combining these groups therefore preserves similar baseline-quartile representation, although it does not eliminate baseline differences within quartiles. As a sensitivity analysis, we combined all Q1–Q3 recipients first and split the scores at a single median. That pooled split produced a larger observed contrast, but it also produced a larger difference in geometric mean baseline titer between the two groups, because a single cutoff no longer balances the baseline quartiles. The within-quartile split is the more conservative of the two, and it is the one used throughout. Both comparisons use the same fitted scores and the same recipients. The single-median intervals hold scores and assignments fixed; the tests that repeat model selection reported in Table S4 apply only to the within-quartile procedure.

Within the individual quartiles the comparison is less precise than in the pooled Q1–Q3 groups, and for several antigen-by-quartile cells the conditional intervals for the fold-rise ratio include 1 and those for the response-rate difference include 0, the values that represent no difference between higher- and lower-score groups (supplemental Table S4).

###### S2.1.4 Tests accounting for model selection

Ordinary tests of association with cross-validated predictions can be anti-conservative.[55] We therefore used a restricted recipient wild bootstrap to calibrate these comparisons under a linear M1 conditional-mean null.[51] An unpenalized linear regression of the three log fold-rises on the three baseline log titers provided fitted means and leverage-adjusted (HC2) residuals. Each recipient's entire block of residuals across episodes and antigens was multiplied by $-1$ or $+1$ with equal probability (a Rademacher wild bootstrap) and added back to the fitted means, generating outcomes with no cellular effect beyond baseline titers. In each of 999 replicates we reselected the Leiden resolution, retuned the M3 penalty $\lambda$ and coefficients, and recomputed the scores and within-quartile median splits, holding the training-fold micro-centroids, candidate cluster maps, baseline quartiles and cross-validation folds fixed.

Test statistics were the high-minus-low mean log fold-rise and the fourfold-response rate difference, for the composite and for each antigen in Q1–Q4 and pooled Q1–Q3. The two-sided Monte Carlo $P$ value was $(1 + \sum_b I\{|T_b^*| \geq |T_{\text{obs}}|\})/1000$. For the per-antigen tests, Holm adjustment covered all 30 contrasts (three antigens, five strata, two endpoints); the ten composite contrasts were adjusted among themselves.[56] These exploratory tests assess whether score-group differences exceed those expected under the baseline-titer model, assuming symmetric recipient-level residuals. Repeating supervised selection incorporates uncertainty from learning the score; the plotted intervals instead condition on the fitted scores and groups. Supplemental Table S4 reports raw and adjusted values.

###### S2.1.5 Score centering and continuous associations

Because each fold has a different atlas, scores can have fold-specific locations: fold means of the raw composite score ranged from $-0.70$ to $-0.05$, against a within-fold SD of about 0.6. Subtracting each fold's mean score before pooling leaves the within-quartile splits and the reported contrasts unchanged in form. The Spearman correlation between the composite cellular composition score and the observed composite log fold-rise was 0.392, and 0.384 after removing the ranks of baseline titer from both variables. Supplemental Figure S11 shows the range of observed responses at similar cellular scores and the score-group comparisons for all three antigens; its lines are least-squares fits within baseline-titer quartiles.

#### S2.2 Signed predictor importance

For feature $j$, Figure 3 shows

$$I_j = d_j \left( \frac{1}{B} \sum_{b=1}^{B} \widehat{\theta}_j^{(b)} \right), \qquad B = 200, \tag{9}$$

where $\widehat{\theta}_j^{(b)}$ is the composite M4 coefficient from recipient-bootstrap refit $b$, expressed in the feature's original units, and $d_j$ is the sample standard deviation of that predictor across the 260 recipients in the shared atlas design. The same $d_j$ is used for every refit. It is not the standard deviation of the coefficient estimates. The bars are standardized conditional associations with log fold-rise, not percentages of explained variance.

Let $x_{itv} = \log(\text{titer}_{itv,\text{pre}})$ and $q_{itv} = \log(\text{titer}_{itv,\text{post}})$, so $y_{itv} = q_{itv} - x_{itv}$. If $\widehat{\gamma}_{vv}$ denotes the coefficient of the same antigen's baseline log titer in the fold-rise model, the corresponding coefficient in the implied post-vaccination log-titer model is $1 + \widehat{\gamma}_{vv}$:

$$\widehat{q}_{itv} = x_{itv} + \widehat{y}_{itv} = \widehat{a}_v + (1 + \widehat{\gamma}_{vv})x_{itv} + \sum_{u \neq v} \widehat{\gamma}_{uv} x_{itu} + \text{clinical and cellular terms.} \tag{10}$$

Thus, a negative baseline coefficient can indicate a smaller fold-rise while remaining consistent with a higher post-vaccination titer. This conversion uses the unstandardized coefficient, not the bar length in Figure 3. The same algebra applies to the composite outcome, whose baseline predictor is the mean baseline log titer across antigens: the negative baseline-titer coefficients in the composite model mean that a higher starting titer predicts a smaller proportional rise, not a lower titer after vaccination.

#### S2.3 Feature-selection stability

Feature-selection stability describes how consistently each population contributed across resampled datasets. Two hundred bootstrap samples of recipients were refitted with the composite M4 model, using the shared 35-population atlas to define the cellular features. The atlas and the reference penalty were fixed; the latter was selected by recipient-grouped cross-validation for M4 using the shared-atlas features. Ten of the 35 populations had nonzero coefficients in at least 50% of refits: cluster 4 (98.5%), cluster 8 (97.0%), cluster 32 (96.0%), cluster 27 (84.0%), cluster 17 (80.0%), cluster 29 (73.5%), cluster 1 (70.0%), cluster 33 (56.0%), cluster 26 (54.5%) and cluster 28 (52.5%; supplemental Figures S4, S5, and S6). Their standardized coefficients were $+0.224$ (cluster 4), $+0.152$ (32), $+0.125$ (27), $+0.046$ (26), $+0.041$ (33), $-0.028$ (28), $-0.086$ (1), $-0.103$ (29), $-0.125$ (17) and $-0.189$ (8). Eight of the ten pointed in the same direction in children and in adults (supplemental Table S6). Baseline titers were selected in 98.0–100% of refits, high-dose vaccine in 99.5%, sex and months since transplantation in 90.0% each and age in 67.5%. These descriptive stability frequencies are not $P$ values, and the fixed shared 35-population atlas is for interpretation rather than independent validation.

#### S2.4 Score groups and effects at a fixed baseline titer

Two readings of the composite cellular composition score are reported here: a comparison of score groups within baseline-titer quartiles, and the association estimated at a fixed baseline titer.

In the pooled comparison across the lower three baseline-titer quartiles, higher-score recipients had a geometric mean fold-rise across antigens of 9.9 versus 3.7 (ratio 2.71; 95% CI, 1.66–4.43), and 22.9 percentage points more of them had a composite fold-rise of at least four (supplemental Figure S13). Recipient-bootstrap confidence intervals hold fitted scores and score groups fixed, so they do not reflect model fitting. The null-bootstrap tests, by contrast, repeat model fitting: in each of 999 replicates, null outcomes were generated for all three antigens, the number of populations was reselected on those outcomes with the inner-validation rule of the primary analysis, and the composite model was refitted on their mean. The tests gave raw $P = .003$ for the pooled Q1–Q3 fold-rise ratio and $P = .018$ for the response difference, and Holm-adjusted values of $P = .030$ and $P = .162$ across the 10 composite comparisons. The Q1 fold-rise ratio gave raw $P = .019$ and adjusted $P = .162$ (supplemental Table S5).

Quartiles group recipients with somewhat different baseline titers, so we also estimated the association at a fixed baseline titer. Log fold-rise was regressed on the cellular score, the baseline log titer and indicators for the cross-validation folds. The score was centered within each cross-validation fold by subtracting that fold's mean score, because each fold has its own atlas. The baseline predictor was the log titer of the same antigen or, for the composite, the mean baseline log titer across antigens. At a fixed baseline titer, a difference in log fold-rise is a difference in log post-vaccination titer. The score percentiles were calculated across all recipients after this centering. Recipients at the 75th percentile of the score were estimated by regression to have post-vaccination titers 2.07 (H1N1), 2.56 (H3N2), 2.21 (B/Victoria) and 2.25 (composite) times those at the 25th percentile with the same baseline titer (supplemental Table S9). As for the score groups, these intervals hold fitted scores fixed and do not account for model fitting.

#### S2.5 Models for the three antigens separately

The composite outcome is the primary one. As a sensitivity analysis we also fitted the three antigen log fold-rises jointly, as a multitask elastic net with the `mgaussian` family, the same $\alpha = 0.9$ and the same `lambda.min` rule. The three measurements of an episode were kept in the same cross-validation fold and resampled together in the bootstrap, so the intervals account for the correlation among antigens within a recipient (Pearson correlations of the log fold-rises, 0.65 to 0.66). One number of populations was selected per outer fold and shared by the three antigens: the multitask penalty selects a common set of features across antigens by construction, so antigen-specific selection is not independently reproduced.

Out-of-fold mean squared error across the three antigens was 2.942 (M1), 2.808 (M2), 2.614 (M3) and 2.566 (M4); supplemental Table S3 gives the per-antigen errors and every pairwise comparison. Adding immune-cell composition to baseline titers reduced it by 11.2% (95% CI,

4.6–17.6%; $P = .003$), and adding composition to titers plus clinical factors by 8.6% (2.6–14.2%; $P = .006$). Clinical factors alone gave 4.5% ($P = .082$), both blocks together 12.8% ($P = .002$), clinical factors added to titers plus composition 1.8% ($P = .25$), and composition in place of clinical factors 6.9% ($P = .056$). Per antigen, composition beyond titers gave 8.7% for H1N1 (MSE 2.852 to 2.603; $P = .027$), 12.4% for H3N2 (3.445 to 3.019; $P < .001$) and 12.2% for B/Victoria (2.529 to 2.220; $P = .004$).

The per-antigen models therefore give the same conclusion as the composite, with smaller estimates and intervals that are wider relative to the estimates: splitting one outcome into three correlated ones costs precision without changing the direction of any comparison. Score groups defined from the per-antigen scores are reported with raw and Holm-adjusted $P$ values side by side in supplemental Table S4, where the adjustment covers all 30 contrasts (3 antigens, 5 strata, 2 endpoints). The pooled Q1–Q3 contrasts are consistent across antigens before adjustment (fold-rise ratios 2.10, 3.07 and 3.58 for H1N1, H3N2 and B/Victoria; raw $P = .020$, $P = .002$ and $P = .002$); after adjustment across 30 contrasts, B/Victoria retains adjusted $P = .030$ for its Q1 fold-rise comparison and its pooled response comparison, and the other antigens do not. We report both and read them together: an adjustment over 30 correlated contrasts of one underlying signal is conservative, and the primary evidence for added predictive value is the model comparison, not these score-group contrasts.

#### S2.6 Cellular scores from fewer markers and from fewer populations

Cellular scores were recomputed from the reduced-marker cell assignments of supplemental Section S4. For each fold, the same sample of up to 2,000 cells per episode was assigned to the fold's atlas using all 30 markers or the fold's 15 top-ranked markers, and M3 and its composite analogue were refitted. Before comparing scores, we subtracted each cross-validation fold's mean score, because every fold has its own atlas. Scores from 15 and 30 markers computed from the same cells were highly correlated (Pearson $r = 0.91$ to 0.93 for the three antigens and 0.91 for the composite), and 85–86% of recipients fell in the same score group. Pooled Q1–Q3 fold-rise ratios were somewhat smaller with 15 markers (composite 2.03, versus 2.29 with 30 markers from the same cells and 2.71 with all cells; supplemental Figure S21).

We next asked whether a few populations could provide similar predictive information to the full composition. For this analysis, the geometry of the shared atlas was built once from all cells without using outcomes. Its number of populations, 35, lies within the range that the inner cross-validation of the primary analysis selected inside training folds (25 to 42 across the 5 outer folds; supplemental Section S1.2). The primary analysis instead rebuilt the atlas within each training fold. Using the shared 35-population atlas and the primary outer folds, we refitted M3 on 100 recipient bootstrap resamples inside each training fold and ranked populations by their bootstrap-averaged standardized coefficient. CLR values were standardized using means and standard deviations from the training fold only. The three most positive and the three most negative populations formed an unweighted signature. We calculated this score as the mean standardized CLR value of the positive populations minus that of the negative populations, then applied the same calculation to the held-out fold. This single predictor was added to the three baseline titers. The selected populations could differ across training folds.

For the composite outcome, titers plus the six-population signature reduced MSE relative to titers alone by 18.6% (95% CI, 9.1–27.0%), against 18.1% (9.3–26.4%) for titers plus all 35 shared-atlas populations (supplemental Table S7). Six unweighted populations therefore carry most of what the full weighted composition carries, which is what makes a reduced panel plausible. Removing the four lineage-negative populations (clusters 3, 19, 24 and 29) and recomputing composition from the other 31 gave the same gain (18.6%; 9.7–26.9%), so the signal does not rest on cells that the panel could not assign to a lineage. CD21-low B cells (cluster 4) and activated double-negative T cells (cluster 32) were positive members in all five folds, naive $CD8^+$ T cells (cluster 27) in four and $CD25^+$ effector-memory $CD4^+$ T cells (cluster 16) in one. The negative members were $CCR4^+$ central-memory $CD8^+$ T cells (cluster 8) and cycling monocytes (cluster 17) in all five folds, activated lineage-negative cells (cluster 29) in four and exhausted effector-memory $CD8^+$ T cells (cluster 1) in one.

#### S2.7 Direction of shared predictors in each cohort

Supplemental Table S6 gives, for the 10 populations selected in at least half of the pooled composite model's bootstrap refits, how often each was selected and the direction of its coefficient when the composite model with titers and composition was refitted in 200 bootstrap samples of each cohort, the refits behind supplemental Figure S16. Each coefficient was averaged over the refits and multiplied by the population's SD in that cohort. Eight of the 10 pointed in the same direction in children and in adults. The two exceptions were the populations with the smallest pooled coefficients among the ten: plasmablasts (cluster 28; $-0.042$ in children, $+0.039$ in adults) and senescent CD4$^+$ T cells (cluster 33; $-0.082$ and $+0.152$).

#### S2.8 Clinical summaries of the cellular composition score

Because each primary fold has its own atlas, we standardized each fold's out-of-fold scores to mean 0 and SD 1 before pooling them. Because the score is on the scale of predicted log fold-rise, one SD of the raw composite score corresponds to an $\exp(0.64) \approx 1.9$-fold difference in predicted fold-rise. Fifths of the standardized composite score were defined over the 260 first-season recipients (52 in each fifth) and used for every cohort.

The association between the score and a fourfold composite rise was estimated by logistic regression on the log geometric-mean baseline titer and the standardized score. We added a score-by-cohort interaction to test whether the association differed between children and adults, and in a separate model adjusted for log months since transplantation, vaccine dose, age and cohort. A linear regression of the composite log fold-rise on the same terms gave the fold-rise ratio per SD. Spearman correlations used the observed composite log fold-rise; the partial correlation removed the ranks of baseline titer from both variables. AUCs used the out-of-fold composite predictions of M1 and M3 (and of M2 and M4). Among recipients with a pre-vaccination HAI titer below 1:40 for an antigen, we report the percentage reaching at least 1:40 four weeks after the second dose. Comparing the lowest fifth of the score with the other four fifths, this was 50% (11 of 22) versus 71% (70 of 98) for H1N1, 24% (6 of 25) versus 73% (66 of 91) for H3N2, and 29% (4 of 14) versus

87% (77 of 89) for B/Victoria (supplemental Figure S9B).

Training-aware $P$ values used the same 999 null replicates described above. Each statistic in supplemental Table S1 was recomputed on every replicate, and $P = (1 + \sum_{b=1}^{B} I\{|T_b^*| \geq |T_{\text{obs}}|\})/(B+1)$. These tests ask whether the association exceeds what the model-fitting pipeline produces when cellular composition carries no information beyond baseline titers.

#### S2.9 Score groups within baseline-titer quartiles

To express the cellular composition score (Section 2.5) in terms of observed antibody responses, we compared score groups at similar baseline antibody levels. In the pooled comparison of recipients from the lower three baseline-titer quartiles, higher-score groups had 2.1- to 3.6-fold larger geometric mean fold-rises for all three antigens, although individual responses overlapped substantially.

Within each baseline-titer quartile, we compared recipients above versus at or below the median cellular composition score for that outcome. For the combined Q1–Q3 comparison, we combined the three higher-score groups and, separately, the three lower-score groups. This preserves approximately equal representation of each baseline quartile in the two groups. Geometric mean fold-rise ratios (higher versus lower score) were 2.10 for H1N1, 3.07 for H3N2 and 3.58 for B/Victoria (supplemental Figure S11), with fourfold-response differences of 17.7, 24.9 and 28.9 percentage points. For the composite outcome, combined Q1–Q3 geometric mean fold-rises were 9.9 versus 3.7 (ratio 2.71), with a 22.9 percentage-point difference in fourfold response. A regression adjusting for baseline titer estimated post-vaccination titers at the 75th percentile of the composite score to be 2.2 times those at the 25th percentile (95% CI, 1.8–2.8; supplemental Table S9). A simpler, unweighted signature built from three positively and three negatively associated populations, chosen inside each training fold, gave a gain close to that of all 35 populations on the shared atlas (supplemental Section S2.6).

For the composite outcome, the combined Q1–Q3 fold-rise comparison had raw $P = .003$ under the null bootstrap that repeats model fitting, and Holm-adjusted $P = .030$ across the ten composite comparisons; the response-rate comparison had raw $P = .018$ and adjusted $P = .162$

(supplemental Table S5). Among the per-antigen contrasts, which carry a 30-comparison adjustment, B/Victoria kept adjusted $P = .030$ for its Q1 fold-rise comparison and for its combined response-rate comparison; the other antigens did not (supplemental Table S4). These exploratory comparisons illustrate the cellular signal; the overall model comparisons provide the primary evidence for added predictive value.

##### S2.10 Training in one cohort and testing in the other

To compare the same cell populations across cohorts, the shared 35-population atlas was held fixed. Test recipients' antibody outcomes were excluded when fitting these prediction models, but their cells had helped define the shared clusters. These analyses therefore evaluate prediction with a shared atlas, rather than independently validating atlas construction. For transfer between cohorts, we compared the cellular elastic net with an unpenalized model containing baseline titers and clinical factors. Both models were fitted in one cohort and evaluated in the other, with the composite outcome. A cellular model trained in children predicted the adults better than the unpenalized model, with 40.9% lower test error (95% CI, 32.4–48.4%; $P < .001$). The reverse direction did not transfer: trained in adults and tested in children, the cellular model gave 12.7% higher error ($-32.3$ to 6.0%; $P = .21$; supplemental Figure S17). The transfer is therefore asymmetric, and the pediatric cohort, the larger of the two with 147 of the 260 recipients, is the one that supplies a model the other cohort can use. Prediction gains across baseline-titer quartiles also differed between cohorts (supplemental Figure S15).

#### S3 Additional results and sensitivity analyses

##### S3.1 Selected numbers of cell populations

Inner cross-validation on the composite outcome selected 42, 35, 42, 37 and 25 cell populations across the five outer folds (median 37), at resolutions 5.0, 4.5, 4.75, 4.5 and 3.25. Selected resolutions varied, and several candidate resolutions had similar validation errors (supplemental Figure S1). Fixing the number of populations instead of selecting it left the gain in the same range (Section S1.2).

#### S3.2 Micro-cluster count and joint antigen prediction

**Micro-cluster count.** In the all-episode analysis (Section S3.7), repeating the nested cross-validation procedure with 2,000 micro-clusters gave 12.9% M1→M3 improvement versus 13.9% with 5,000. Selected meta-cluster counts were 5, 5, 8, 34 and 32 versus 8, 33, 31, 5 and 28. Both settings showed resolution variability (supplemental Figure S23).

**Joint versus separate fitting of the three antigens.** In the same all-episode analysis, with recipient-grouped penalty tuning, elastic nets fitted to one antigen at a time improved 13.2% over M1 for M3 and 16.7% for M4, compared with 13.9% and 17.6% for the joint multitask fits. The small differences support similar predictive performance for joint and separate fitting (supplemental Figure S22).

#### S3.3 Interactions among baseline titers and clinical factors

To allow the association of one baseline predictor with response to depend on another, $\mathrm{M2_{int}}$ added all 21 pairwise product terms to the seven M2 predictors: three baseline log titers, dose, age, sex and transplant timing. For example, a dose-by-timing term allows the dose association to vary with time since transplantation. $\mathrm{M4_{int}}$ added the immune-cell composition features to that same interaction model, without interactions involving cellular features. M2 and M4 included the corresponding predictors without product terms. All six models were single-outcome elastic nets on the composite outcome.

All six models used the same recipient-grouped outer training/test splits. Within each outer training set, M1–M4 shared five-fold penalty tuning, whereas $\mathrm{M2_{int}}$ and $\mathrm{M4_{int}}$ shared three-fold penalty tuning. Thus, outer test recipients were identical across comparisons, but the interaction sensitivity analysis used a different inner tuning scheme from the primary analysis. Cellular models reused the original outer-fold atlases and selected resolutions.

Table S8 expresses every model's prediction gain relative to the same titers-only baseline, M1. Adding interactions did not improve prediction over M2 or M4. Immune-cell composition retained predictive value in the interaction model: $\mathrm{M4}_{\mathrm{int}}$ improved 15.7% over M1, compared with 3.1% for $\mathrm{M2}_{\mathrm{int}}$. Using $\mathrm{M2}_{\mathrm{int}}$ as the denominator, the cellular gain was 13.0%; the separate comparison of M3 with $\mathrm{M2}_{\mathrm{int}}$ gave 12.6%. These percentages use different denominators and are not additive.

#### S3.4 Antigen responses and time since transplantation

An association with time since transplantation would support immune recovery as an interpretation of the cellular signal. Regressing episode-specific months since transplant on CLR cluster features explained 49.9% of its variance in this cohort ($R^2 = 0.499$). The largest absolute Spearman correlations were for cluster 3 ($\mathrm{CD38}^+$ lineage-negative; $\rho = -0.52$) and cluster 23 (DC-like APC; $\rho = -0.46$).

Pearson correlations between antigen log fold-rises were 0.65 for H1N1–H3N2, 0.65 for H1N1–B/Victoria, and 0.66 for H3N2–B/Victoria. Geometric mean baseline titers were 42.3, 48.1, and 48.1, respectively; H3N2 therefore did not have a markedly lower overall baseline titer. These summaries are descriptive; row-level correlation and paired-test $P$ values were not used because the three antigen measurements come from the same recipient.

Per-episode prediction benefit was the M1 squared error minus the M3 squared error on the composite outcome. Its linear slope against episode-specific months since transplant was $0.011$ (95% recipient-bootstrap CI, $-0.022$ to $0.043$) error units per month. This descriptive check did not identify a linear trend.

Clinical models and resolution selection used months since transplantation at the vaccination visit, recorded for all 260 recipients (median, 6.3; range, 3.0–35.9), and age at consent.

#### S3.5 Cohort differences across baseline antibody levels

Using the shared 35-population atlas and separate prediction-model fits, adult M1→M3 gains were greatest in the lowest baseline-titer quartile (39.1%, against 26.8%, 19.3% and $-32.2\%$ in the other three), whereas pediatric gains were greatest in the highest quartile (29.0%, against $-0.2\%$, 7.2% and 5.7%; supplemental Figure S15). In the pooled analysis the gain was largest in the lower two quartiles (18.3% in Q1 and 14.9% in Q2, against 12.5% in Q3 and 7.9% in Q4). These exploratory patterns do not establish a common baseline-titer threshold across age groups.

#### S3.6 Second-season episodes

Eighty-one second-season episodes had cytometry. Seven episodes (six adult and one pediatric) had no recorded pre-vaccination (4) or post-vaccination (3) titer and were excluded, because their fold-rises are undefined. Of the remaining 74, 72 came from recipients with a first-season episode in the analysis set and 2 from recipients whose first-season episode was not in it; those 2 enter the season-level validation but not the within-recipient comparisons. Each second-season episode was mapped to the atlas of the outer fold holding out its recipient and scored with that fold's models, and its score was standardized with that fold's first-season mean and SD, so both seasons are on the first season's scale. The recorded interval between a recipient's two episodes was 6.9–14.8 months (median 11.5). The adult second-season immunogenicity and safety results were reported separately.[57]

Prediction error for the composite outcome fell by 39.2% when immune-cell composition was added to baseline titers (95% recipient-bootstrap CI, 28.3–48.9%), and the area under the curve for a fourfold composite rise rose from 0.648 to 0.706 (difference 0.058; $-0.029$ to 0.145; DeLong $P = .20$). The odds ratio per SD of the score was 1.81 (95% CI, 0.92–3.54; $P = .086$), smaller and less precise than in the 260 first-season recipients.

Response after a single dose differed between seasons: 57% of returning recipients (39 of 69) reached a fourfold composite rise four weeks after the first dose in the second season, against 23% (16 of 69) of the same recipients and 23% (60 of 259) of all recipients in the first season. In the first season the score predicted this one-dose response (odds ratio per SD, 2.59; 95% CI, 1.73–3.87; $P < .001$), whereas in the second season it did not (1.34; 0.69–2.58; $P = .38$), consistent with a ceiling once most recipients respond.

Scores rose from the first to the second season in all four response-transition groups, with median increases of 0.69 SD in the 16 recipients who responded in neither season, 0.93 SD in the 22 who became responders, 0.12 SD in the 6 who stopped responding and 0.25 SD in the 28 who responded in both. Median baseline titers moved in opposite directions: 0.34 times the first-season value in recipients who became responders and 16.8 times in those who stopped responding (0.56 and 2.67 times in the two groups whose status did not change). Among the 38 recipients who did not respond in the first season, the second-season score did not differ between those who responded in the second season and those who did not (median 0.88 versus 0.63 SD; Wilcoxon $P = .31$), and neither did the change in score (0.93 versus 0.69 SD; $P = .36$). Under the first-season logistic model of Figure 4C, the median modeled probability of a fourfold rise rose from 0.31 to 0.68 in recipients who became responders, rose from 0.22 to 0.39 in those who responded in neither season, fell from 0.75 to 0.18 in those who stopped responding, and fell from 0.57 to 0.41 in those who responded in both, whose higher second-season starting titers outweighed their higher scores.

#### S3.7 All episodes of every recipient

The primary analysis uses one episode per recipient, so the atlases, the resolution selection and the fitted models are free of within-recipient repetition. As a sensitivity analysis, the same pipeline was run on all 334 episodes with complete titers, from 262 recipients, with both episodes of a returning recipient kept in the same outer fold and resampled together. The numbers in this subsection come from that all-episode pipeline with the multitask outcome, the three antigens modeled jointly, and are not comparable term by term with the composite first-season results reported elsewhere. Adding immune-cell composition to baseline titers reduced prediction error by 13.9% (95% CI, 7.9–19.7%; $P < .001$), and adding it to titers plus clinical factors by 8.6% (95% CI, 3.3–14.2%; $P = .002$). The cellular score behaved as in the primary analysis: 21% of episodes in its lowest fifth had a fourfold composite rise, compared with 56% of the remaining episodes, and each SD was associated with 2.3-fold higher odds of a fourfold rise at a given baseline titer. Supplemental Figures S22 and S23 come from this all-episode analysis.

#### S3.8 Responders and non-responders with the same score

Recipients with a similar cellular composition can still differ in response. Within each fifth of the composite score, we compared responders with non-responders on baseline titer, clinical factors, lineage shares and population shares, as standardized differences pooled across fifths (supplemental Figure S14). At the same score, responders had markedly lower baseline titers (standardized difference $-1.01$) and a longer interval since transplantation ($+0.67$). Among cellular features, responders had fewer $CD38^+$ lineage-negative cells (cluster 3, $-0.55$), fewer activated lineage-negative cells (cluster 29, $-0.50$), a smaller lineage-negative share overall ($-0.47$), fewer $TIGIT^+$ lineage-negative cells (cluster 24, $-0.35$) and more naive $CD8^+$ T cells (cluster 27, $+0.33$). After residualizing each feature on the baseline titer within the fifth, most cellular differences shrank but kept their direction, so part of what distinguishes responders at the same score is their lower starting titer.

#### S3.9 Fitted predictions under the two vaccine-dose models

To describe fitted antibody responses under the two vaccine-dose models, separate random forests were fitted in the high- and standard-dose groups and applied to all episodes. These partly in-sample predictions are shown in supplemental Figure S25. They are not independently validated counterfactual outcomes and do not establish who benefits from a dose change. Lower and higher prediction groups were defined by the bottom or top quartile under both fitted dose models.

### S4 Reduced-marker panels and rare cell populations

The reduced-marker analysis assessed whether a smaller cytometry panel could retain predictive information from the full-panel atlas. Within each training-fold atlas, markers were ranked by variation between meta-cluster centers. Cells were assigned to the nearest full-panel micro-centroid using only the retained markers, and M4 was refitted with recipient-grouped penalty tuning. The same sample of up to 2,000 cells per episode was used at every marker budget; the full-panel reference used that same sampling depth. The consensus 15-marker set comprises CD19, CD20, CD3, CD4, CD8, CD21, CXCR5, CCR6, CCR2, HLA-DR, CD86, CD25, CD38, CD27 and CD57.

#### S4.1 Marker ranking and model refitting

Marker ranking used the variance across meta-cluster centers of each standardized marker, with each meta-cluster center calculated by averaging its constituent micro-centroids. Thus, markers were ranked by separation of the existing clusters, not by their association with antibody response. The numbers of meta-clusters were fixed at 42, 35, 42, 37 and 25 in the five outer folds for every marker budget. Micro-centroids and their mapping to meta-clusters were held fixed; only the marker coordinates used for cell assignment were reduced, followed by re-estimation of M4 coefficients and penalty strength. This evaluates prediction after adapting the model to the reduced panel; the atlas geometry and micro-cluster-to-meta-cluster mapping remain fixed.

At eight versus six markers, test RMSE for the composite response was 1.410 versus 1.380, while mean agreement with full-panel cell labels fell from 32.1% to 28.0%. No smaller panels were tested. Label agreement measures consistency with the full-panel assignments, not accuracy against independently established cell identities. Different assignments can preserve predictive information if they retain similar between-recipient composition differences, and the refitted prediction model can adapt to the changed features. The small RMSE differences are compatible with redundant marker information, changes in measurement noise, or sampling variation. They do not identify an optimal marker set.

##### S4.2 Cell-label agreement

To assess cell-label agreement separately from prediction, 200,000 cells in the shared 35-population atlas were reassigned to its fixed micro-centroids using each reduced marker set. We compared the new meta-cluster label with the full-panel label, separately among all cells and among the 39,059 cells in the populations selected in at least 50% of bootstrap refits. This is a cell-weighted label-agreement measure, not the percentage of populations recovered, and it does not re-cluster the reduced panel. At 15 markers, agreement was 49.3% among cells in selected populations and 53.8% overall. This shared-atlas analysis is separate from the per-fold prediction analysis in Figure 7.

##### S4.3 Rare-population abundance

Cluster sizes were summarized as the median and interquartile range of per-episode cell fractions. Clusters 32, 33, 28 and 4 had median frequencies of 0.204%, 0.225%, 0.463% and 0.481%, respectively, and all four were stably selected. Cluster 32 had a median of 80 measured cells per episode. No cluster was excluded on frequency alone. Five episodes had fewer than 10,000 profiled cells, including two with fewer than 1,000; rare-cluster estimates in these episodes warrant caution. These rare populations contributed reproducibly to model fitting; their measurement precision and biological annotations require separate assessment.

#### S5 Illustrative assay-noise calculation

##### S5.1 Assumed noise model

To put prediction errors on an interpretable scale, the following calculation considers an assumed level of assay noise. The noise variance was not measured in this study. For episode $(i, t)$ and outcome $v$, here the composite, write

$$y_{itv} = f_v(\mathbf{x}_{it}) + \epsilon_{itv}, \tag{11}$$

where $f_v(\mathbf{x}_{it})$ is the response component predictable from the available features. For this illustration, the residual $\epsilon_{itv}$ is assumed to represent independent, mean-zero measurement noise with variance $\sigma_\epsilon^2$. Suppressing the indices,

$$\text{Var}(y) = \text{Var}\big(f(\mathbf{x})\big) + \sigma_\epsilon^2. \tag{12}$$

#### 1215 S5.2 Illustrative HAI error

Hemagglutination inhibition (HAI) titers are reported in doubling dilutions (e.g., 1:10, 1:20, 1:40, ...). One dilution step equals $\log 2 \approx 0.693$ log-units. The following calculation is illustrative: it assumes that the standard deviation of the combined error in log fold-rise is one dilution step. This is not an empirically estimated noise variance for this dataset:

$$\sigma_\epsilon \approx \log 2 \approx 0.693, \qquad \sigma_\epsilon^2 \approx (\log 2)^2 \approx 0.48. \tag{13}$$

#### 1216 S5.3 Observed and assumed variance

From the observed data, the variance of the composite outcome is $\text{Var}(y) \approx 3.217$. Substituting into Equation (12):

$$\text{Var}\big(f(\mathbf{x})\big) \approx 3.217 - 0.480 = 2.737. \tag{14}$$

#### 1217 S5.4 Implied upper bound on prediction

Under this assumption, the minimum expected prediction error is $\sigma_\epsilon^2$. The corresponding upper bound on the coefficient of determination is

$$R^2_{\text{ceiling}} = 1 - \frac{\sigma_\epsilon^2}{\text{Var}(y)} = 1 - \frac{0.480}{3.217} \approx 0.851. \tag{15}$$

#### S5.5 Comparison with observed model errors

Table S10 compares model errors with the assumed noise variance. The 15.3% relative MSE improvement (M1→M3) corresponds to closing about 19% of the gap between the titers-only model and the assumed noise floor:

$$\text{Gap closed} = \frac{\text{MSE}_{\text{M1}} - \text{MSE}_{\text{M3}}}{\text{MSE}_{\text{M1}} - \sigma_\epsilon^2} = \frac{2.277 - 1.929}{2.277 - 0.480} \approx 0.194. \tag{16}$$

Under the illustrative $0.480$ variance assumption, the $R^2$ ceiling is approximately 85%. If instead pre- and post-vaccination titers each had independent one-dilution standard-deviation errors, their log-ratio error variance would be $2(\log 2)^2 \approx 0.96$, giving a ceiling near 70% and a gap-closed estimate near 26%. Neither assumption is a measured assay-noise estimate, so these calculations provide scale context rather than a validated performance ceiling.

#### S6 Supplemental tables

**Supplemental Table S1: Clinical summaries of the cellular composition score.** Composite cellular scores are out-of-fold and standardized within cross-validation fold; a fourfold rise is a geometric mean fold-rise across the three antigens of at least four. Brackets give 95% intervals: recipient-level bootstrap ($B = 2{,}000$) for rates, correlations, AUCs and MSE reductions, and recipient-clustered (sandwich) intervals for odds and fold-rise ratios. Conventional $P$ values are Wald tests with recipient-clustered standard errors, DeLong tests for AUCs, or recipient-bootstrap tests for MSE reductions. Training-aware $P$ values come from 999 null-bootstrap replicates that repeat resolution selection, penalty tuning and coefficient fitting under no cellular effect beyond baseline titers; the smallest attainable value is $1/(B + 1)$. The odds ratios are adjusted for baseline titer; clinical factors are months since transplantation, vaccine dose, age and cohort. MSE reductions are for the composite outcome.

| Statistic | Recipients | Estimate [95% interval] | $P$ | Training-aware $P$ |
|---|---|---|---|---|
| Fourfold rise, lowest fifth vs other fifths (%) | All recipients | 15 vs 51; difference 36 [23, 47] | – | ≤.001 |
| Fourfold rise, lowest fifth vs other fifths (%) | Children | 22 vs 47; difference 25 [5, 44] | – | – |
| Fourfold rise, lowest fifth vs other fifths (%) | Adults | 10 vs 57; difference 47 [31, 62] | – | – |
| Odds ratio for fourfold rise per SD | All recipients | 2.15 [1.55, 2.98] | <.001 | .007 |
| Odds ratio for fourfold rise per SD | Children | 1.80 [1.16, 2.82] | .009 | .041 |
| Odds ratio for fourfold rise per SD | Adults | 3.09 [1.85, 5.18] | <.001 | .006 |
| Odds ratio for fourfold rise per SD | All, adjusted for clinical factors | 2.15 [1.52, 3.03] | <.001 | – |
| Ratio of odds ratios, adults vs children | Score-by-cohort interaction | 1.80 [0.90, 3.63] | .097 | – |
| Fold-rise ratio per SD at fixed baseline titer | All recipients | 1.87 [1.60, 2.18] | <.001 | – |
| Spearman correlation with composite fold-rise | All recipients | 0.39 [0.28, 0.49] | – | ≤.001 |
| Spearman correlation with composite fold-rise | Children | 0.37 [0.21, 0.51] | – | .002 |
| Spearman correlation with composite fold-rise | Adults | 0.50 [0.35, 0.62] | – | ≤.001 |
| Partial Spearman correlation given baseline titer | All recipients | 0.38 [0.27, 0.49] | – | ≤.001 |
| AUC, titers (M1) vs titers + composition (M3) | All recipients | 0.75 vs 0.80; difference 0.050 [0.013, 0.089] | .009 | ≤.001 |
| AUC, titers (M1) vs titers + composition (M3) | Children | 0.81 vs 0.83; difference 0.021 [-0.023, 0.068] | .36 | .055 |
| AUC, titers (M1) vs titers + composition (M3) | Adults | 0.66 vs 0.76; difference 0.098 [0.035, 0.163] | .003 | ≤.001 |
| AUC, titers + clinical (M2) vs all predictors (M4) | All recipients | 0.78 vs 0.81; difference 0.034 [0.002, 0.067] | .034 | – |
| Pooled model, MSE reduction M1→M3 (%) | Children | 9.4 [-3.9, 21.2] | .15 | – |
| Pooled model, MSE reduction M2→M4 (%) | Children | 6.7 [-5.9, 17.7] | .26 | – |
| Pooled model, MSE reduction M1→M3 (%) | Adults | 22.9 [13.1, 32.0] | <.001 | – |
| Pooled model, MSE reduction M2→M4 (%) | Adults | 20.1 [10.8, 28.6] | <.001 | – |

**Supplemental Table S2: Cell-population annotations.** Population number, broad lineage, cell type label, and key differentiating markers (mean expression on the original scale) for the 35 populations of the shared atlas. Annotations were assigned using canonical marker profiles; the CyTOF panel lacks dedicated myeloid markers (CD14, CD16), so APC/monocyte annotations were inferred by exclusion ($CD3^{-}CD19^{-}HLA\text{-}DR^{+}$). Key markers distinguish a population from others of its lineage; low values mark markers it lacks. Low/high in labels refer to relative marker expression, not population frequency; double-negative T cells (DNT) have a $CD3^{+}CD4^{-}CD8^{-}$ phenotype. Plasmablasts are listed with B cells. CM, central memory; EM, effector memory; EMRA, effector memory re-expressing CD45RA, assigned here as $CD45RO^{-}CCR7^{-}$ because the panel has no CD45RA; Mono, monocyte; APC, antigen-presenting cell.

| # | Lineage | Cell type label | Key markers |
|---|---|---|---|
| *B cell / Plasmablast* | | | |
| 4 | B cell | CD21-low B | CD19=5.0, CD21=0.97, CD27=0.95 |
| 9 | B cell | Activated B | CD19=4.4, OX40=1.2, CD137=1.3, CD138=1.3 |
| 12 | B cell | Cycling B | CD19=4.2, Ki-67=1.7 |
| 13 | B cell | Naive B BTLA high | CD19=4.5, BTLA=2.1, CXCR5=3.6, CD21=3.1 |
| 26 | B cell | $CCR7^{-}$ B cell | CD19=3.5, CXCR5=2.3, CCR7=0.67 |
| 28 | B cell | Plasmablast | CD38=5.1, Ki-67=1.8, CD19=2.8, CD20=0.92 |
| 30 | B cell | Transitional B | CD19=4.0, CD38=3.4, CD27 low |
| 31 | B cell | $CD25^{+}$ B | CD19=4.3, CD25=1.6, CXCR5=3.1 |
| 34 | B cell | Naive B BTLA low | CD19=4.0, CCR7=1.5, BTLA=0.66 |
| *$CD4^{+}$ T cell* | | | |
| 6 | $CD4^{+}$ T | EM $CD4^{+}$ T $CD25^{-}$ | CD4=4.0, CD45RO=2.6, ICOS=1.6, CD25=0.47 |
| 7 | $CD4^{+}$ T | $EMRA^{+}$ $CD4^{+}$ T | CD4=3.1, CD45RO=1.0, CCR7=0.78, CD27=2.2 |
| 10 | $CD4^{+}$ T | Naive $CD4^{+}$ T $CD25^{+}$ | CD4=3.6, CCR7=1.6, CD25=1.9, CD127=2.6 |
| 14 | $CD4^{+}$ T | Naive $CD4^{+}$ T $CD25^{-}$ | CD4=3.6, CCR7=1.9, CD45RO=0.94, CD25=0.30 |
| 16 | $CD4^{+}$ T | EM $CD4^{+}$ T $CD25^{+}$ | CD4=3.9, CD45RO=2.6, CCR4=2.4, CD25=2.5 |
| 25 | $CD4^{+}$ T | CM $CD4^{+}$ T | CD4=4.0, CD45RO=3.0, PD-1=2.2, ICOS=1.9, CXCR5=0.95 |
| 33 | $CD4^{+}$ T | Senescent $CD4^{+}$ T | CD4=4.1, CD57=3.5, CD27=1.3 |
| *$CD8^{+}$ T cell* | | | |
| 1 | $CD8^{+}$ T | Exhausted EM $CD8^{+}$ T | CD8=3.6, CD45RO=2.4, PD-1=1.7, TIGIT=1.6 |
| 8 | $CD8^{+}$ T | CM $CD8^{+}$ T $CCR4^{+}$ | CD8=2.4, CCR7=1.3, CCR4=1.5, CD45RO=1.6 |
| 15 | $CD8^{+}$ T | $CD57^{+}$ $CD8^{+}$ T | CD8=3.2, CD57=3.4, CD27=1.4, CD28=1.2 |
| 18 | $CD8^{+}$ T | $EMRA^{+}$ $CD8^{+}$ T $TIGIT^{+}$ | CD8=3.1, TIGIT=2.0, CD45RO=0.82, CCR7=0.52 |
| 22 | $CD8^{+}$ T | $EMRA^{+}$ $CD8^{+}$ T $TIGIT^{-}$ | CD8=3.4, TIGIT=0.44, CD45RO=1.0, CCR7=0.59 |
| 27 | $CD8^{+}$ T | Naive $CD8^{+}$ T | CD8=4.0, CCR7=1.5, CD27=3.4, CD45RO=0.75 |
| *APC / Monocyte* | | | |
| 2 | APC/Mono | Resting Mono | CCR2=3.0, HLA-DR=2.6, CD69=0.26 |
| 5 | APC/Mono | Activated Mono | CCR2=3.3, HLA-DR=3.3, CD69=1.8 |
| 11 | APC/Mono | Activated APC | HLA-DR=1.7, CD86=0.44, CD69=1.7 |
| 17 | APC/Mono | Cycling Mono | CCR2=3.3, CD86=1.5, Ki-67=1.1, CD69=0.39 |
| 23 | APC/Mono | DC-like | HLA-DR=3.1, CD86=1.7, CCR2=0.69 |
| 35 | APC/Mono | Pan-positive APC | CCR2=2.7, HLA-DR=3.0, OX40=1.7, CD137=1.7 |
| *NK cell* | | | |
| 21 | NK | NK-like | CD57=3.7, CD38=3.0, CD137=1.3, CD3=0.75 |
| *Double-negative T / Lineage-negative* | | | |
| 20 | DN T | Resting DNT | CD3=4.0, CD4=0.52, CD8=0.60, CD38=1.1 |
| 32 | DN T | Activated DNT | CD3=3.9, CD38=4.0, TIGIT=2.2, Ki-67=1.4 |
| 3 | $Lineage^{-}$ | $CD38^{+}$ $lineage^{-}$ | CD38=2.8, all lineage low |
| 19 | $Lineage^{-}$ | Low-expression $lineage^{-}$ | All markers $<1.8$ |
| 24 | $Lineage^{-}$ | $TIGIT^{+}$ $lineage^{-}$ | TIGIT=1.8, CD38=3.1 |
| 29 | $Lineage^{-}$ | Activated $lineage^{-}$ | CD138=1.4, CD38=3.0, CXCR3=1.4 |

**Supplemental Table S3: Models for the three antigens separately.** The primary analysis predicts the composite response (Table 2); here the three antigen log fold-rises are predicted jointly by a multitask elastic net, which selects one feature set for all three. The three measurements of a recipient stay in the same cross-validation fold and are resampled together. **A.** Mean squared error of the out-of-fold predictions. **B.** Percent reduction in mean squared error between models, overall and by antigen, with two-sided $p$ from a recipient-level paired bootstrap ($B = 2{,}000$). The overall intervals are 11.2% (4.6 to 17.6) for M1→M3 and 8.6% (2.6 to 14.2) for M2→M4.

**A. Prediction error by model**

| Model | Overall | H1N1 | H3N2 | B/Victoria |
|---|---|---|---|---|
| M1 | 2.942 | 2.852 | 3.445 | 2.529 |
| M2 | 2.808 | 2.725 | 3.242 | 2.459 |
| M3 | 2.614 | 2.603 | 3.019 | 2.220 |
| M4 | 2.566 | 2.558 | 2.931 | 2.210 |

**B. Percent reduction in error, $p$ in parentheses**

| Comparison | Overall | H1N1 | H3N2 | B/Victoria |
|---|---|---|---|---|
| M1→M3 (composition over titers) | 11.2 (0.003) | 8.7 (0.027) | 12.4 (<0.001) | 12.2 (0.004) |
| M1→M4 (full over titers) | 12.8 (0.002) | 10.3 (0.019) | 14.9 (<0.001) | 12.6 (0.008) |
| M2→M4 (composition over clinical) | 8.6 (0.006) | 6.1 (0.077) | 9.6 (0.001) | 10.1 (0.008) |
| M2→M3 (composition vs clinical) | 6.9 (0.056) | 4.5 (0.265) | 6.9 (0.095) | 9.7 (0.040) |
| M1→M2 (clinical over titers) | 4.5 (0.082) | 4.5 (0.147) | 5.9 (0.038) | 2.8 (0.329) |
| M3→M4 (clinical on top of composition) | 1.8 (0.252) | 1.7 (0.337) | 2.9 (0.141) | 0.5 (0.776) |

**Supplemental Table S4: Exploratory cellular-score contrasts and training-aware null-bootstrap tests.** Fold ratio is the ratio of geometric mean observed fold-rises (higher versus lower score). Rate difference is the corresponding fourfold-response difference in percentage points. Each antigen uses its own within-quartile score split; Q1–Q3 retains those assignments. Raw $p$ values compare observed separation with 999 recipient wild-bootstrap replicates under a linear titers-only conditional-mean null, refitting supervised resolution selection, M3 coefficients, penalty and score groups. $p_{\mathrm{H}}$ is Holm-adjusted across all 30 comparisons. These working-null tests incorporate training uncertainty; the conditional intervals in Figures 4 and S11 do not. Effects are descriptive and do not establish causal or individually validated treatment-response differences.

| Antigen | Stratum | Fold ratio | $p$ | $p_{\mathrm{H}}$ | Rate diff. | $p$ | $p_{\mathrm{H}}$ |
|---|---|---|---|---|---|---|---|
| H1N1 | Q1 | 4.04 | 0.022 | 0.440 | 25.9 | 0.046 | 0.644 |
| H1N1 | Q2 | 2.07 | 0.120 | 1.000 | 21.9 | 0.109 | 1.000 |
| H1N1 | Q3 | 1.15 | 0.744 | 1.000 | 6.1 | 0.680 | 1.000 |
| H1N1 | Q4 | 1.34 | 0.436 | 1.000 | 7.1 | 0.476 | 1.000 |
| H1N1 | Q1–Q3 | 2.10 | 0.020 | 0.420 | 17.7 | 0.027 | 0.486 |
| H3N2 | Q1 | 4.30 | 0.025 | 0.475 | 26.0 | 0.039 | 0.608 |
| H3N2 | Q2 | 2.65 | 0.038 | 0.608 | 34.4 | 0.008 | 0.184 |
| H3N2 | Q3 | 2.60 | 0.034 | 0.578 | 15.2 | 0.273 | 1.000 |
| H3N2 | Q4 | 1.27 | 0.522 | 1.000 | 6.8 | 0.429 | 1.000 |
| H3N2 | Q1–Q3 | 3.07 | 0.002 | 0.056 | 24.9 | 0.002 | 0.056 |
| B/Victoria | Q1 | 7.32 | 0.001 | 0.030 | 32.1 | 0.011 | 0.242 |
| B/Victoria | Q2 | 3.51 | 0.004 | 0.100 | 34.4 | 0.007 | 0.168 |
| B/Victoria | Q3 | 1.82 | 0.189 | 1.000 | 21.2 | 0.094 | 1.000 |
| B/Victoria | Q4 | 1.46 | 0.392 | 1.000 | 16.6 | 0.180 | 1.000 |
| B/Victoria | Q1–Q3 | 3.58 | 0.002 | 0.056 | 28.9 | 0.001 | 0.030 |

**Supplemental Table S5: Composite antibody endpoint.** The composite outcome is the mean of the three antigen log fold-rises, the log of the geometric mean fold-rise across antigens. **A.** Percent reduction in out-of-fold mean squared error of the composite outcome, from single-outcome elastic nets refitted on the composite at each fold's resolution. Intervals are from a recipient-level paired bootstrap ($B = 2{,}000$). Composite MSE was 2.277 (M1), 2.140 (M2), 1.929 (M3) and 1.872 (M4). **B.** Composite cellular-score groups within baseline-titer quartiles. Geometric mean composite fold-rise (higher versus lower score), their ratio with recipient-bootstrap 95% CI, and the difference in the percentage of episodes with composite fold-rise of at least four. These intervals hold fitted scores and score groups fixed and do not reflect model fitting. Raw $p$ values compare the observed separation with 999 recipient wild-bootstrap replicates under a titers-only null model, repeating resolution selection, penalty tuning, coefficient fitting and score grouping to account for model fitting. $p_{\mathrm{H}}$ is Holm-adjusted across the ten comparisons in panel B (Supplementary Section S2.4).

**A. Model comparisons**

| Comparison | ΔMSE (%) | 95% CI | $p$ |
|---|---|---|---|
| Add cells to titers (M1→M3) | 15.3 | [7.0, 23.2] | <0.001 |
| Add cells to titers + clinical (M2→M4) | 12.5 | [4.6, 20.3] | 0.001 |
| Add clinical to titers (M1→M2) | 6.0 | [−0.5, 11.9] | 0.072 |
| Add clinical to titers + cells (M3→M4) | 3.0 | [−1.7, 7.2] | 0.196 |
| Add both to titers (M1→M4) | 17.8 | [8.7, 26.5] | 0.001 |
| Cells instead of clinical (M2 vs M3) | 9.9 | [0.6, 18.4] | 0.039 |

**B. Composite cellular-score groups**

| Stratum | Fold-rise | Ratio [95% CI] | $p$ | $p_{\mathrm{H}}$ | Rate diff. | $p$ | $p_{\mathrm{H}}$ |
|---|---|---|---|---|---|---|---|
| Q1 (lowest) | 45.3 vs 9.9 | 4.57 [1.84, 10.65] | 0.019 | 0.162 | 23.1 | 0.083 | 0.492 |
| Q2 | 6.5 vs 2.8 | 2.34 [1.13, 4.81] | 0.036 | 0.252 | 21.9 | 0.116 | 0.492 |
| Q3 | 3.3 vs 1.7 | 1.92 [1.00, 3.58] | 0.082 | 0.492 | 24.2 | 0.086 | 0.492 |
| Q4 (highest) | 1.7 vs 1.2 | 1.42 [0.93, 2.25] | 0.273 | 0.546 | 0.5 | 0.937 | 0.937 |
| Q1–Q3 combined | 9.9 vs 3.7 | 2.71 [1.66, 4.43] | 0.003 | 0.030 | 22.9 | 0.018 | 0.162 |

**Supplemental Table S6: Direction of the stably selected populations in each cohort.** Populations selected in at least 50% of 200 recipient-bootstrap refits of the pooled composite model with titers, clinical factors and immune-cell composition (M4). Pooled coefficient: bootstrap-averaged coefficient times the population's standard deviation. Per cohort: the composite model with titers and composition (M3) was refitted in 200 recipient-bootstrap samples of each cohort with one penalty strength for both cohorts; selection is the percentage of refits with a nonzero coefficient, and the coefficient is averaged over refits and multiplied by the population's standard deviation in that cohort. Positive values indicate a higher predicted composite log fold-rise. 8 of 10 populations had the same direction in both cohorts.

| | | Pooled | Selection (%) | | Coefficient | |
|---|---|---|---|---|---|---|
| Population | Label | coefficient | Children | Adults | Children | Adults |
| 4 | CD21-low B | +0.224 | 53 | 98 | +0.084 | +0.278 |
| 32 | Activated DNT | +0.152 | 88 | 76 | +0.162 | +0.117 |
| 27 | Naive $CD8^+$ T | +0.125 | 57 | 82 | +0.081 | +0.164 |
| 26 | $CCR7^-$ B cell | +0.046 | 34 | 21 | +0.035 | +0.009 |
| 33 | Senescent $CD4^+$ T | +0.041 | 62 | 86 | −0.082 | +0.152 |
| 28 | Plasmablast | −0.028 | 48 | 48 | −0.042 | +0.039 |
| 1 | Exhausted EM $CD8^+$ T | −0.086 | 20 | 92 | −0.001 | −0.272 |
| 29 | Activated $lineage^-$ | −0.103 | 25 | 60 | −0.029 | −0.119 |
| 17 | Cycling Mono | −0.125 | 57 | 29 | −0.109 | −0.038 |
| 8 | CM $CD8^+$ T $CCR4^+$ | −0.189 | 25 | 92 | −0.018 | −0.257 |

**Supplemental Table S7: Six-population signature and lineage-negative populations (shared atlas).** All rows use the shared 35-population atlas, whose geometry was built once from all cells without outcomes, with the number of populations taken from the primary analysis; its reference gain therefore differs from the primary per-fold estimate in Table 2. Percent reduction in out-of-fold mean squared error relative to baseline titers alone, overall across antigens and for the composite outcome, with recipient-level paired bootstrap 95% CIs ($B = 2{,}000$). The signature was chosen inside each training fold (three most positive and three most negative populations by bootstrap-averaged standardized coefficient) and is the mean standardized CLR value of the positive populations minus that of the negative populations, standardized with training-fold means and standard deviations. For the row excluding lineage-negative populations (clusters 3, 19, 24 and 29), we recomputed the centered log-ratio on the remaining 31 populations.

| Model | Overall ΔMSE (%) [95% CI] | $p$ | Composite ΔMSE (%) [95% CI] | $p$ |
|---|---|---|---|---|
| Titers + 35 populations | 12.7 [5.6, 19.5] | <0.001 | 18.1 [9.3, 26.4] | <0.001 |
| Titers + 31 populations, no lineage-negative | 13.3 [6.5, 20.0] | <0.001 | 18.6 [9.7, 26.9] | <0.001 |
| Titers + six-population signature | 14.3 [7.1, 21.0] | 0.001 | 18.6 [9.1, 27.0] | <0.001 |

**Supplemental Table S8: Prediction with interactions among baseline titers and clinical factors.** Overall out-of-fold MSE and relative reduction compared with M1, calculated as $100(1 - \mathrm{MSE}_M/\mathrm{MSE}_{M1})$. All rows use the same outer test folds and the composite outcome; inner tuning is described in supplemental Section S3.3. Percentages use unrounded MSE values.

| Model | Predictors | MSE | Reduction vs M1 (%) |
|---|---|---|---|
| M1 | Baseline titers | 2.277 | 0.0 |
| M2 | Titers + clinical factors | 2.140 | 6.0 |
| $\mathrm{M2}_{\mathrm{int}}$ | M2 + pairwise interactions | 2.207 | 3.1 |
| M3 | Titers + cellular features | 1.929 | 15.3 |
| M4 | Titers + clinical + cellular features | 1.872 | 17.8 |
| $\mathrm{M4}_{\mathrm{int}}$ | $\mathrm{M2}_{\mathrm{int}}$ + cellular features | 1.920 | 15.7 |

**Supplemental Table S9: Post-vaccination titer ratio associated with the cellular score at the same baseline titer.** Log fold-rise was regressed on the cellular score, the baseline log titer of the same antigen (for the composite, the mean baseline log titer across antigens) and indicators for the cross-validation folds. At a fixed baseline titer a fold-rise ratio equals a post-vaccination titer ratio, so the table gives the regression-estimated ratio of post-vaccination titers between episodes at the 75th and 25th percentiles of the score. Each cross-validation fold's mean score was subtracted before calculating percentiles across all episodes. Recipient-bootstrap intervals hold fitted scores fixed and do not account for model fitting.

| Outcome | Titer ratio | 95% CI |
|---|---|---|
| H1N1 | 2.07 | [1.63, 2.63] |
| H3N2 | 2.56 | [2.00, 3.30] |
| B/Victoria | 2.21 | [1.77, 2.76] |
| Composite | 2.25 | [1.82, 2.76] |

**Supplemental Table S10: Model performance under an illustrative assay-noise assumption.** Values are for the composite outcome. The assumed noise variance is 0.480; it was not estimated from these data.

| Model | MSE | RMSE | $R^2$ | % of $R^2$ ceiling |
|---|---|---|---|---|
| M1 (titers only) | 2.277 | 1.509 | 0.292 | 34% |
| M2 (titers + clinical) | 2.140 | 1.463 | 0.335 | 39% |
| M3 (titers + immune composition) | 1.929 | 1.389 | 0.400 | 47% |
| M4 (full) | 1.872 | 1.368 | 0.418 | 49% |
| Assumed noise floor | 0.480 | 0.693 | 0.851 | 100% |

#### S7 Supplemental figures

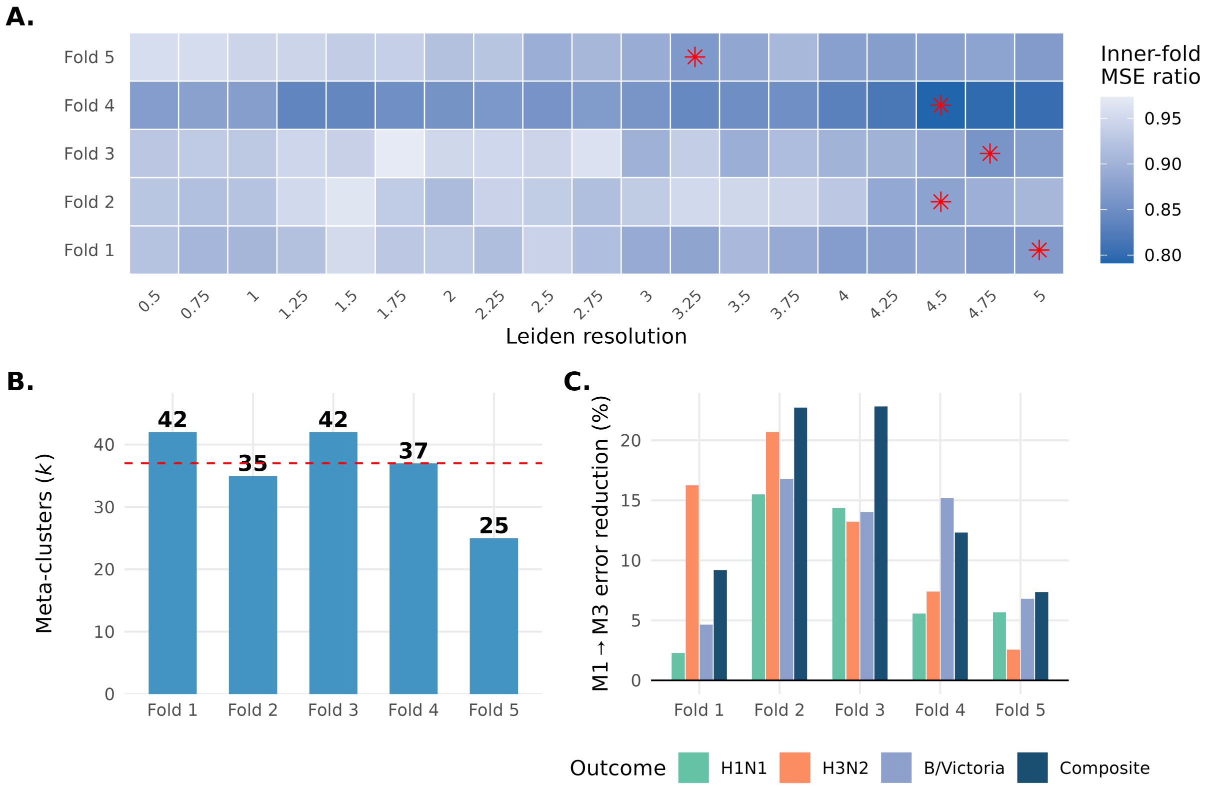


**Supplemental Figure S1: Selection of the number of cell populations. A.** Selection criterion for each candidate clustering resolution and outer fold: the composite-response error of M4 divided by that of the unpenalized baseline-titer-plus-clinical model, averaged over the 3 inner folds. Stars mark the selected resolution; values below 1 favor M4. **B.** Numbers of meta-clusters selected in the five outer folds (42, 35, 42, 37 and 25); the dashed line marks the median of 37. **C.** Held-out reduction in mean squared prediction error from adding immune-cell composition to baseline titers (M1→M3) in each outer fold, for the composite outcome and for each antigen in the models of the three antigens.

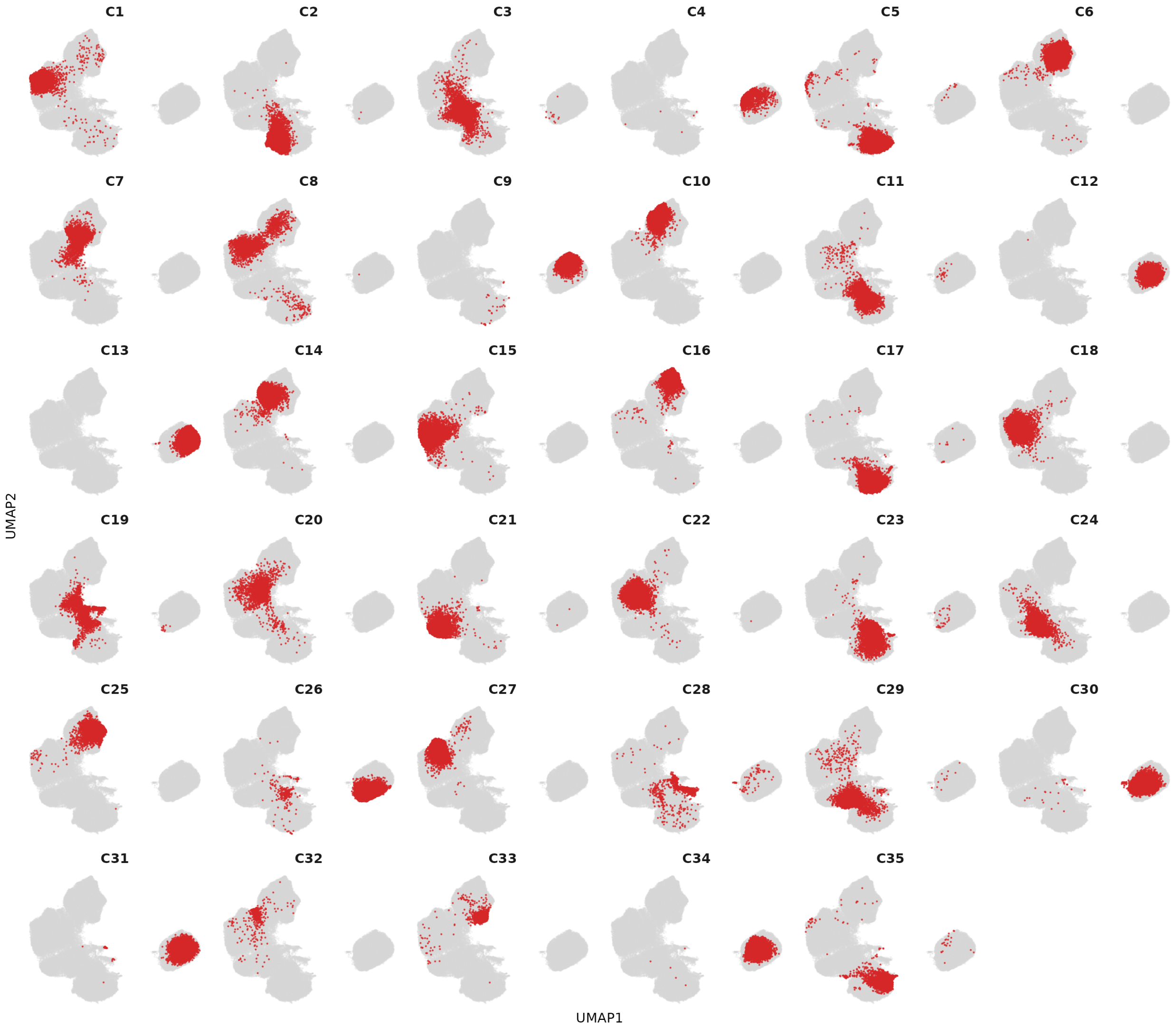


**Supplemental Figure S2: Location of each cell population in the shared atlas.** Each panel highlights one meta-cluster against the other cells in gray. UMAP summarizes marker-profile similarity; marker profiles and annotations are provided in the accompanying heatmap and annotation table.

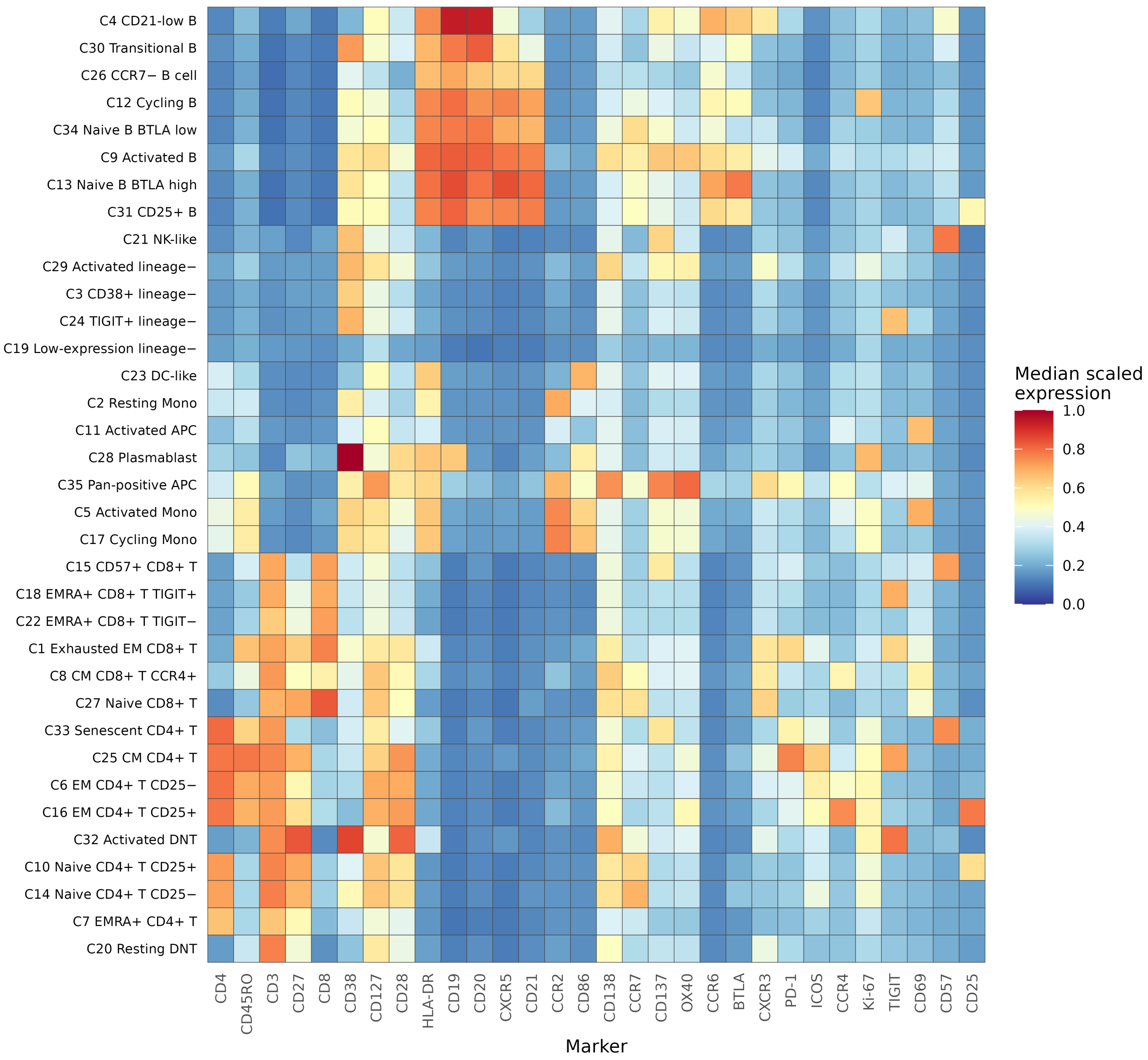


**Supplemental Figure S3: Marker expression across annotated cell populations.** Colors show median relative expression within each of the 35 meta-clusters: blue indicates low expression, pale yellow intermediate expression, and red high expression. Each marker was clipped at its first and 99th percentiles and scaled to $[0, 1]$ across sampled cells before calculating cluster medians. Rows are ordered by hierarchical clustering with Ward linkage. Colors compare relative expression rather than positivity gates. Cluster numbers identify the shared atlas used in this study; label abbreviations are defined in supplemental Table S2.

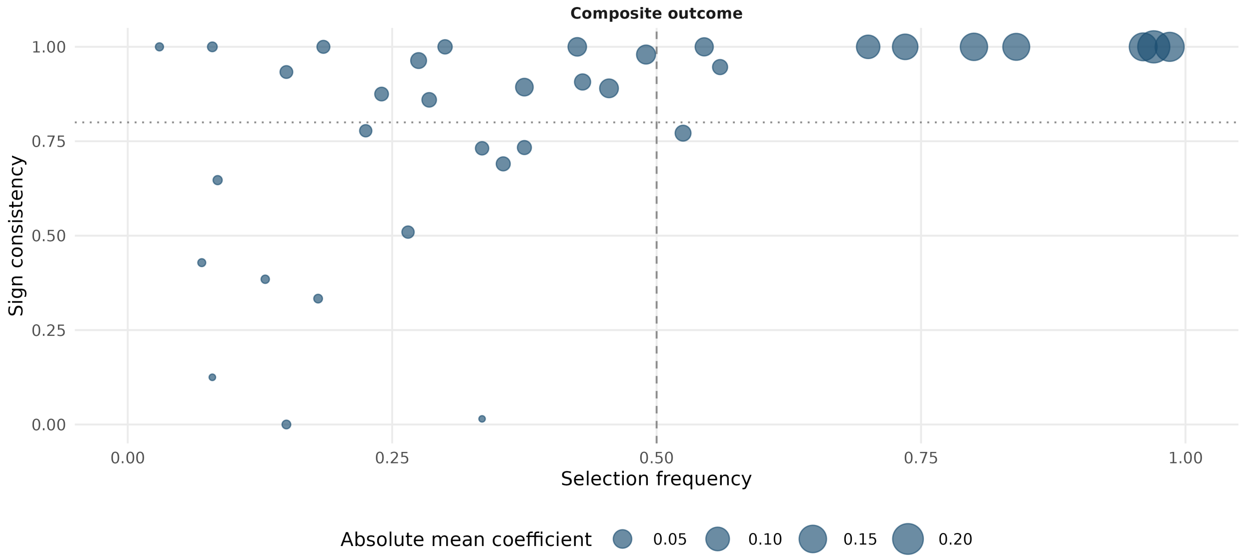


**Supplemental Figure S4: Feature-selection stability.** Selection frequency and sign consistency for the 35 shared-atlas populations across 200 bootstrap composite M4 refits. Point size is the absolute mean coefficient. Dashed vertical and dotted horizontal lines mark 50% selection and 0.8 sign consistency. Ten populations meet the 50% selection criterion; selection frequency is descriptive, not a significance test.

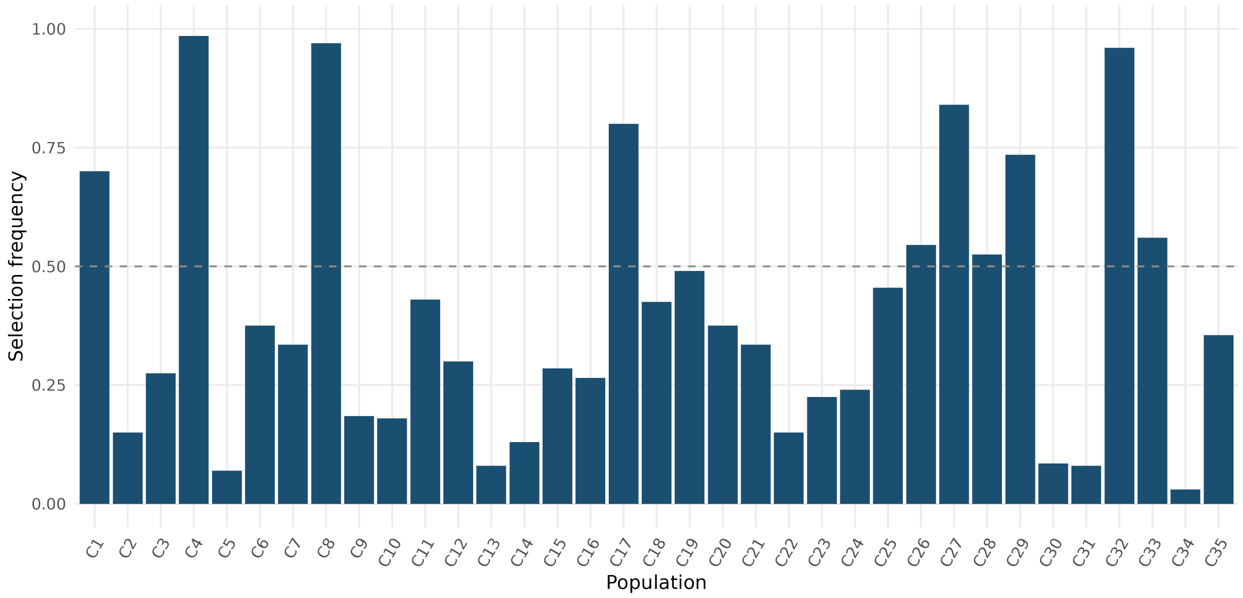


**Supplemental Figure S5: Cluster-selection frequency.** Fraction of 200 bootstrap composite M4 fits retaining each shared-atlas population. The dashed line marks 50%; ten populations meet this criterion. Clusters 4, 8 and 32 are selected in 98.5%, 97.0% and 96.0% of refits.

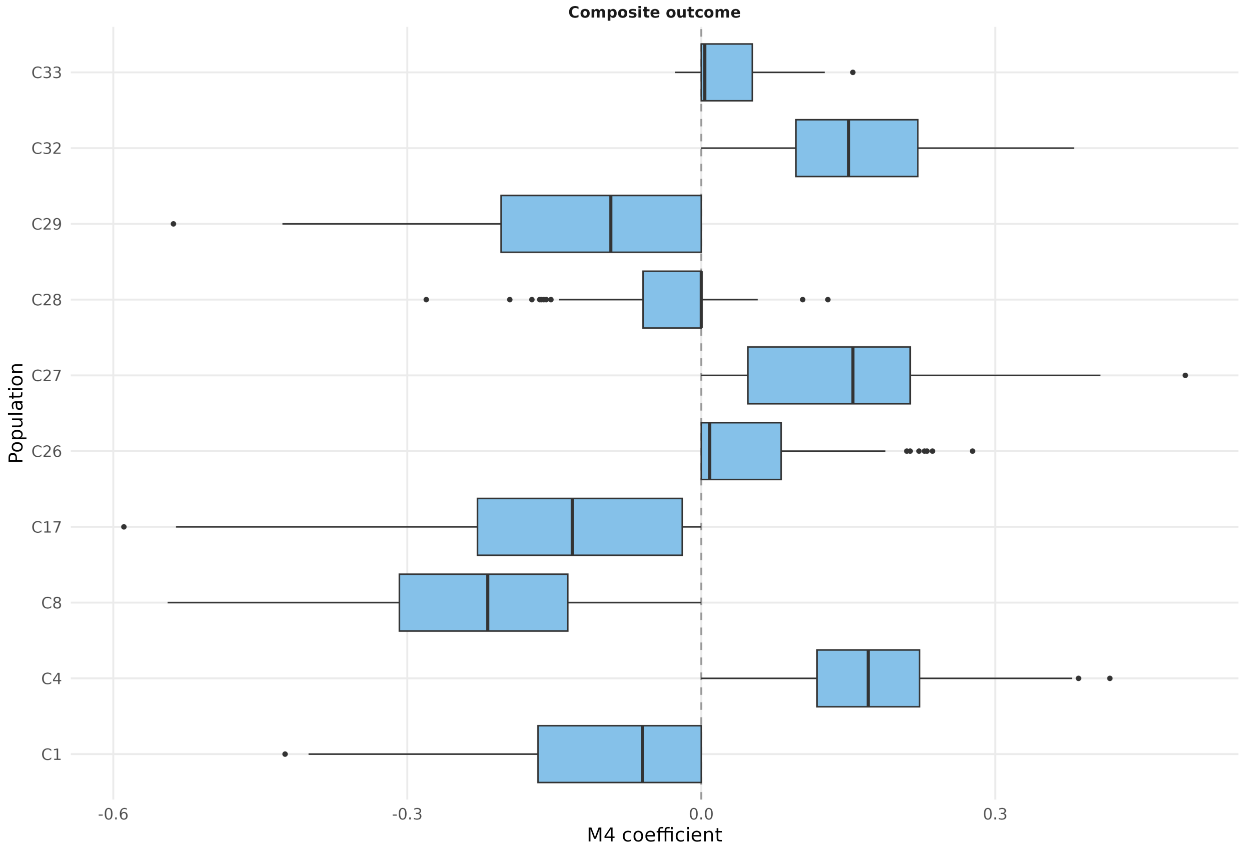


**Supplemental Figure S6: Coefficient distributions for selected clusters.** Composite M4 coefficients from 200 bootstrap refits for the ten populations selected in at least half of refits. Boxes show medians and interquartile ranges; whiskers extend to 1.5 interquartile ranges and points denote remaining values. Atlas geometry and penalty strength were fixed. Positive and negative signs describe conditional model associations.

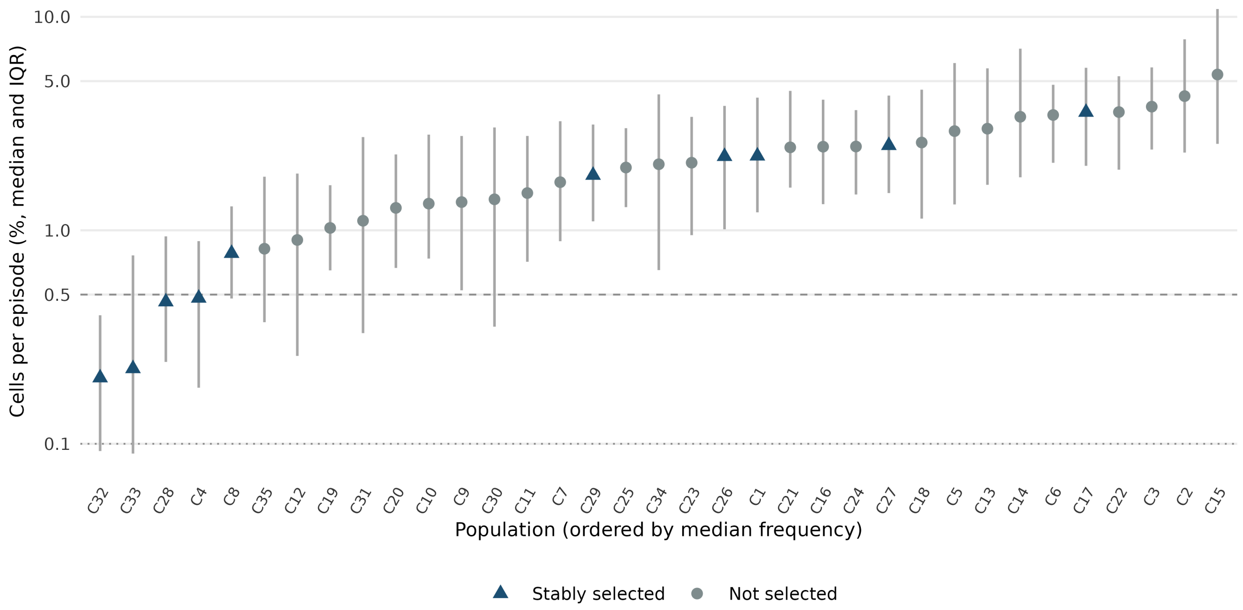


**Supplemental Figure S7: Abundance and selection stability of rare cell populations.** Points show median cell fractions across 260 first-season recipients; whiskers show interquartile ranges. Populations are ordered by median frequency. Blue triangles identify populations selected in at least 50% of 200 composite M4 bootstrap refits; gray circles identify the others. Dashed and dotted lines mark 0.5% and 0.1%, respectively. All four populations with median frequency below 0.5% were stably selected. These frequency thresholds are descriptive, rather than estimates of measurement precision.

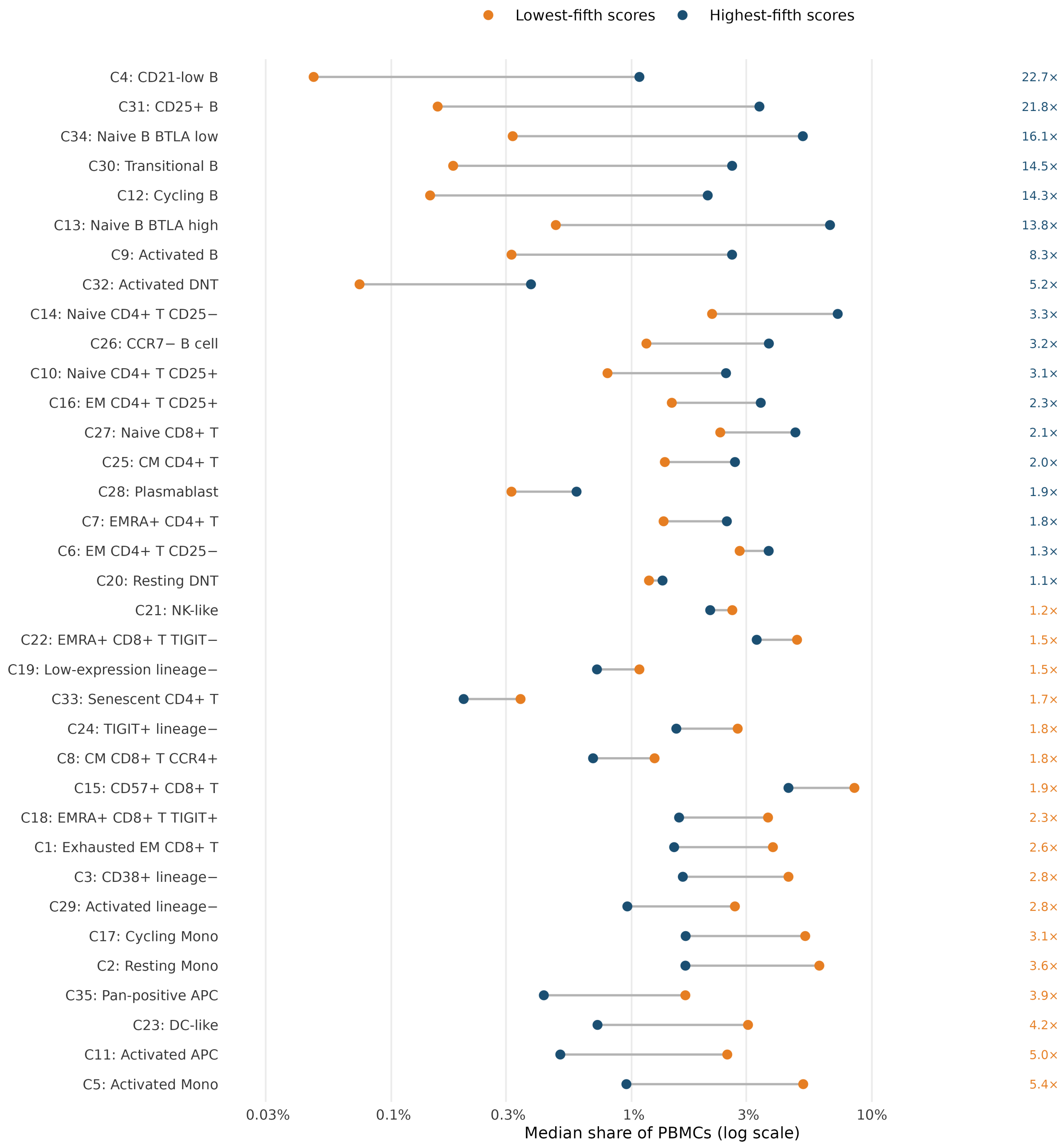


**Supplemental Figure S8: Population composition at low and high cellular scores.** Median share of PBMCs for each of the 35 shared-atlas populations among recipients in the lowest and highest fifth of the composite score; numbers give the fold difference, colored by the group with the larger share.

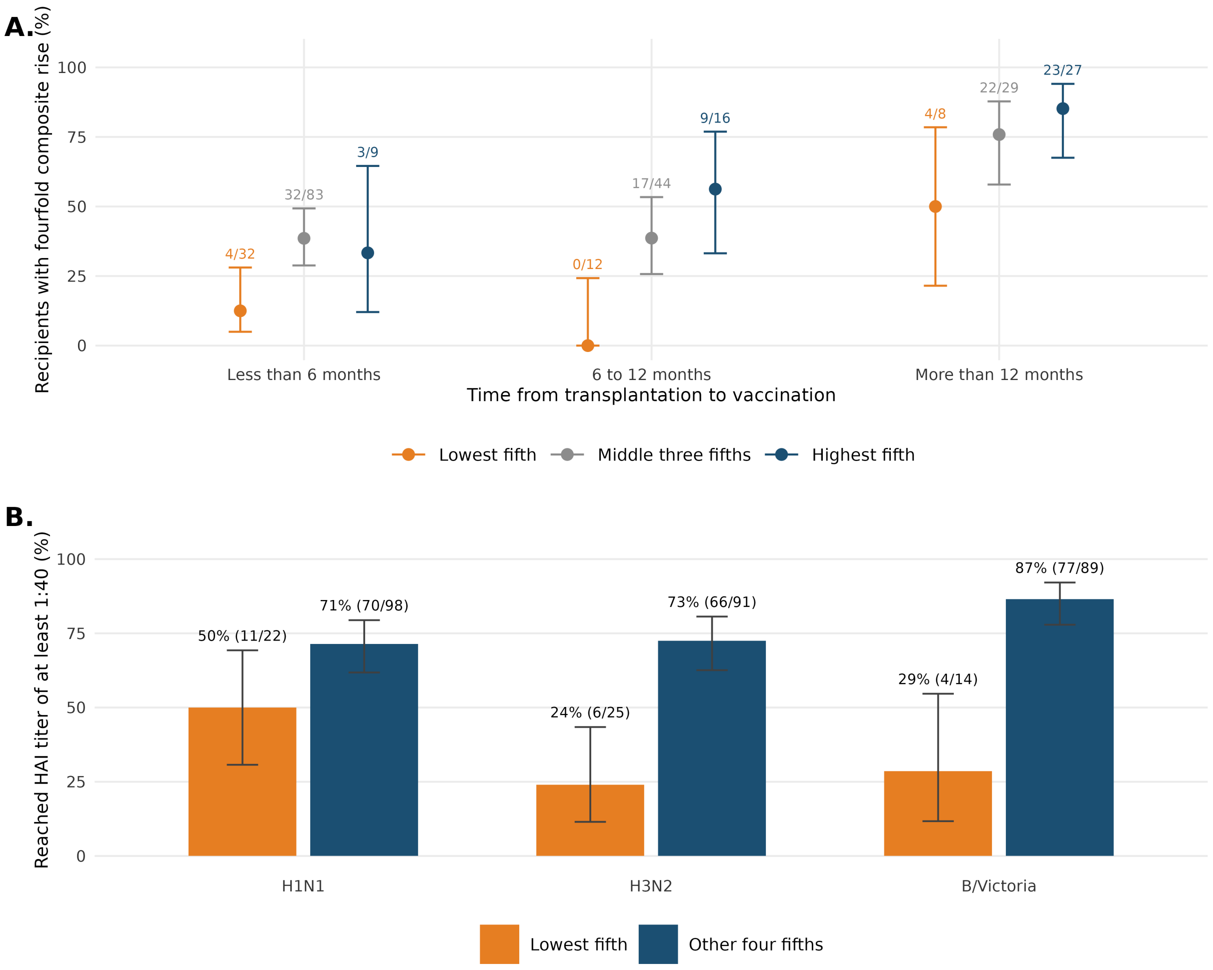


**Supplemental Figure S9: Response by score, time since transplantation, and pre-vaccination protection.** **A.** Percentage of recipients with a fourfold composite rise by time from transplantation to vaccination and by composite-score group (lowest fifth, middle three fifths, highest fifth); labels give responders and recipients, and whiskers 95% Wilson intervals. **B.** Among recipients with a pre-vaccination HAI titer below 1:40 for an antigen, percentage reaching at least 1:40 four weeks after the second dose, for recipients in the lowest fifth of the composite score and the other four fifths.

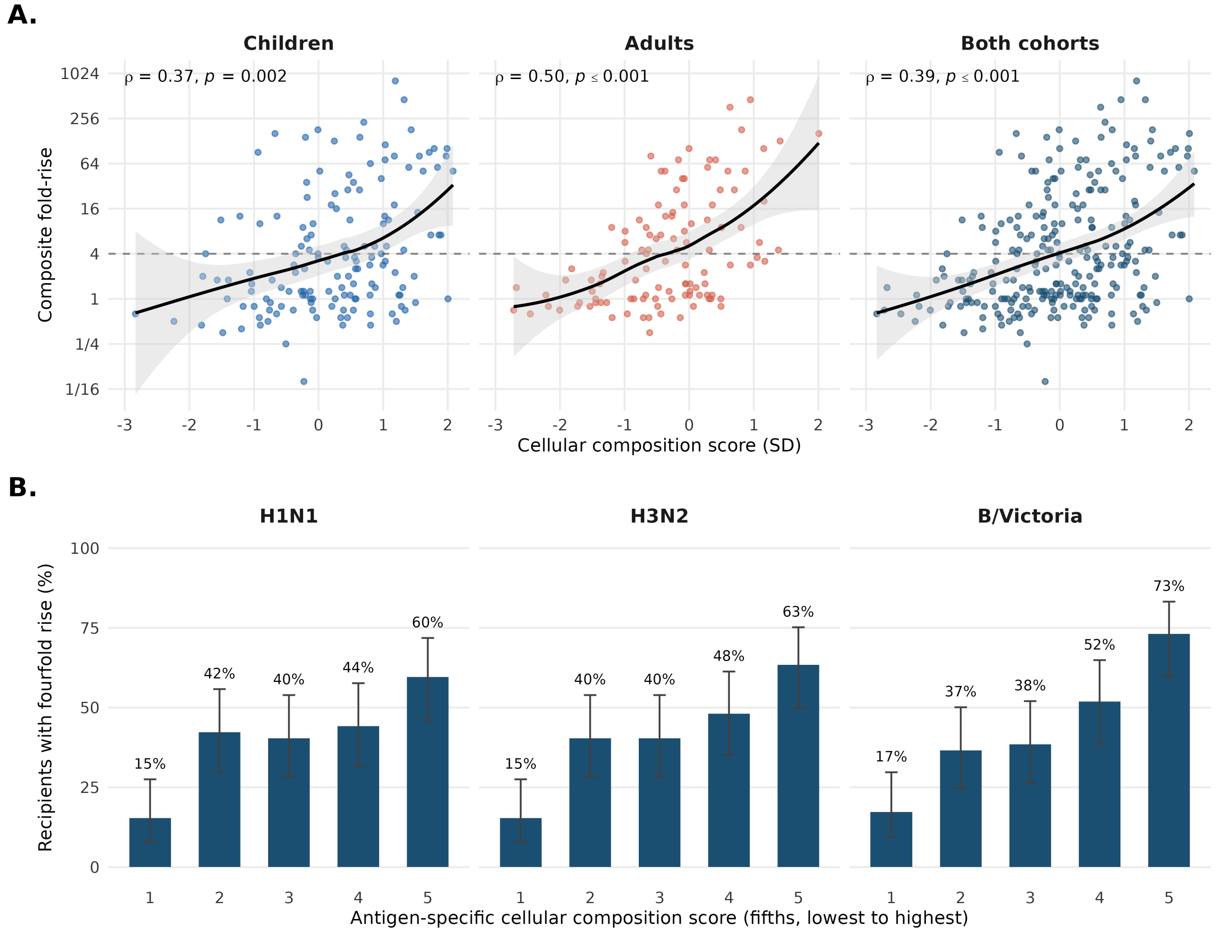


**Supplemental Figure S10: Cellular composition score and observed fold-rise. A.** Composite score against observed composite fold-rise in children, adults, and both cohorts together. Curves are LOESS fits with 95% bands; $\rho$ is Spearman's rank correlation, with the null-bootstrap $P$ value. The dashed line marks a fourfold rise. **B.** Percentage of recipients with a fourfold rise for each antigen, by fifth of that antigen's own cellular score from the models of the three antigens, with 95% Wilson intervals.

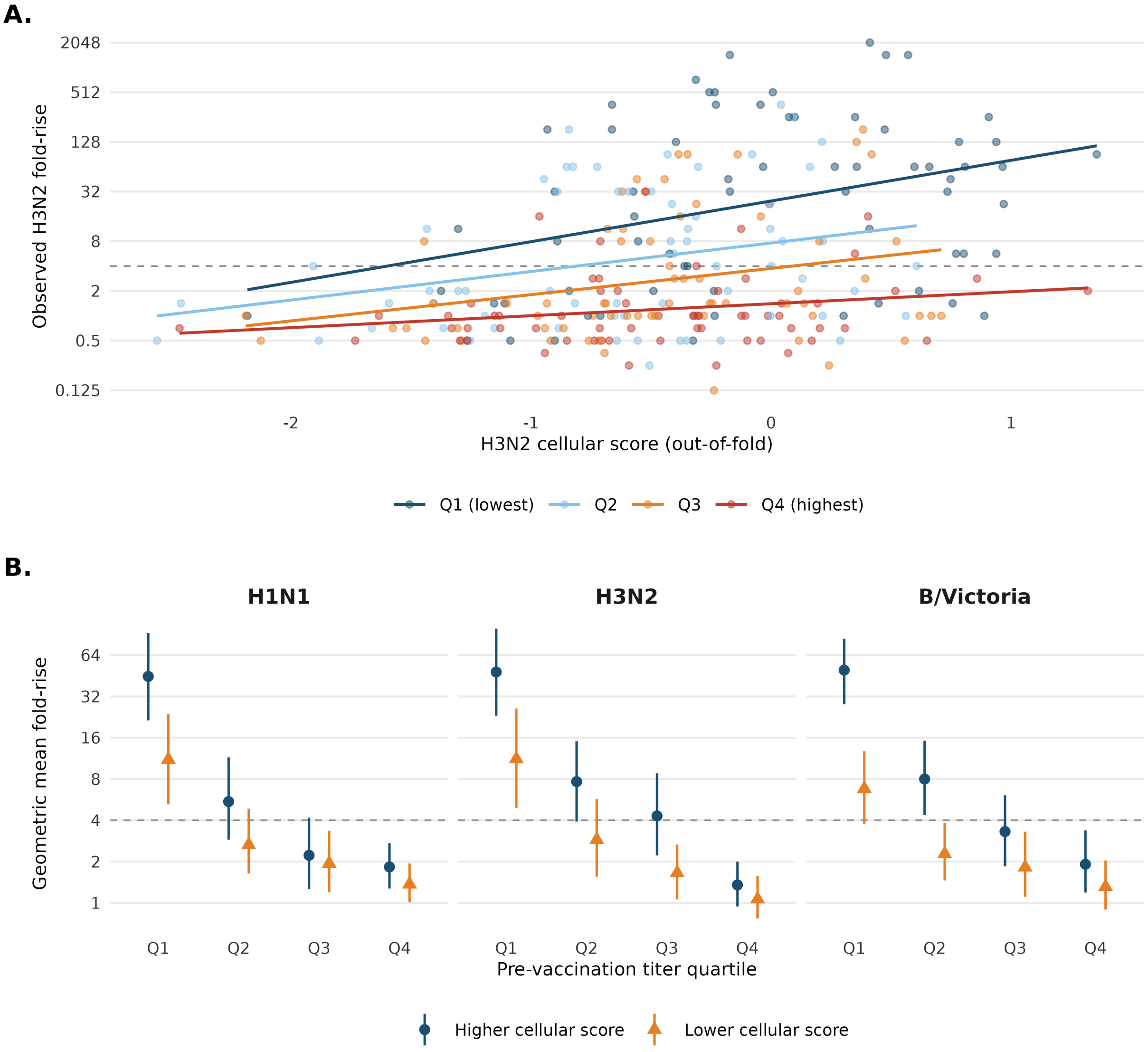


**Supplemental Figure S11: Cellular scores and observed responses across three antigens. A.** Observed H3N2 fold-rise versus cellular score for 260 recipients, colored by baseline-titer quartile, with a least-squares line on the log scale within each quartile. **B.** Geometric mean fold-rise for higher- and lower-score groups within each baseline quartile. Each antigen has its own cellular score and median split. Whiskers show 95% bootstrap intervals with scores and groups held fixed. Dashed lines mark a fourfold rise. Scores include only the cellular terms of models fitted without the recipient. Adjusted tests that repeat model selection are reported in supplemental Table S4.

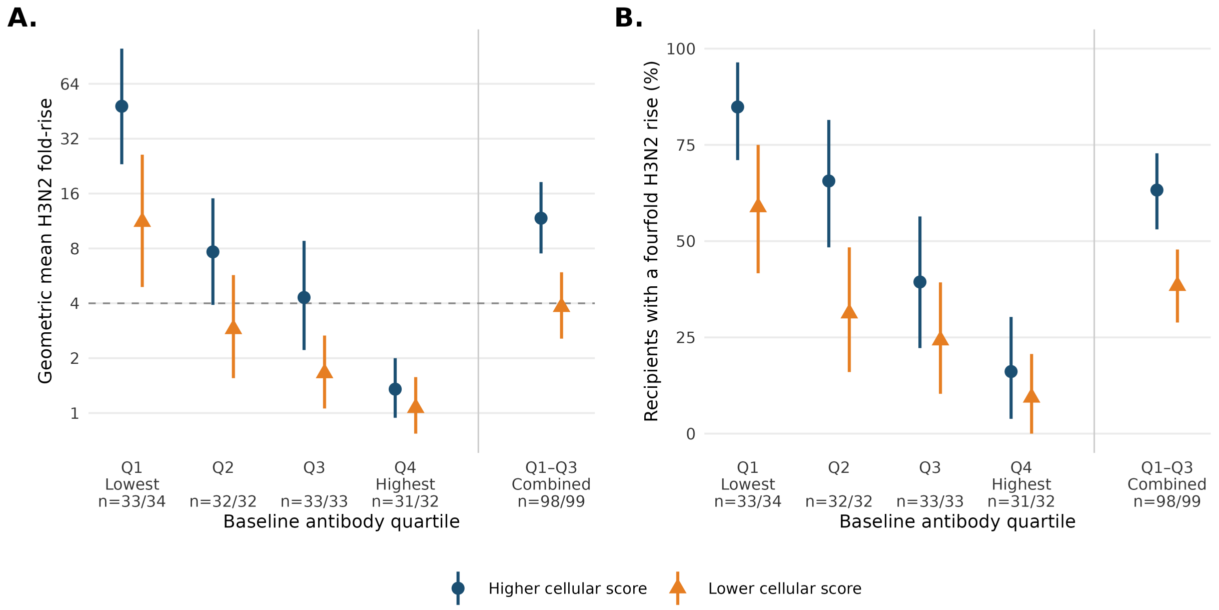


**Supplemental Figure S12: Observed H3N2 antibody responses by cellular score and baseline antibody level.** Cellular scores summarize the cellular contribution to predictions from models fitted without that recipient. Within each quartile of the geometric mean baseline titer across three antigens, recipients were classified above versus at or below the median cellular score. **A.** Geometric mean antibody fold-rise from before vaccination to four weeks after the second dose; the dashed line marks a fourfold rise. **B.** Percentage of recipients achieving at least a fourfold rise. The combined Q1–Q3 comparison pools the groups already defined within each quartile. Counts below the axes give higher/lower-score group sizes. Whiskers show 95% bootstrap intervals with scores and groups held fixed. These exploratory H3N2 comparisons did not remain significant after adjustment for 30 tests; full results are in supplemental Table S4.

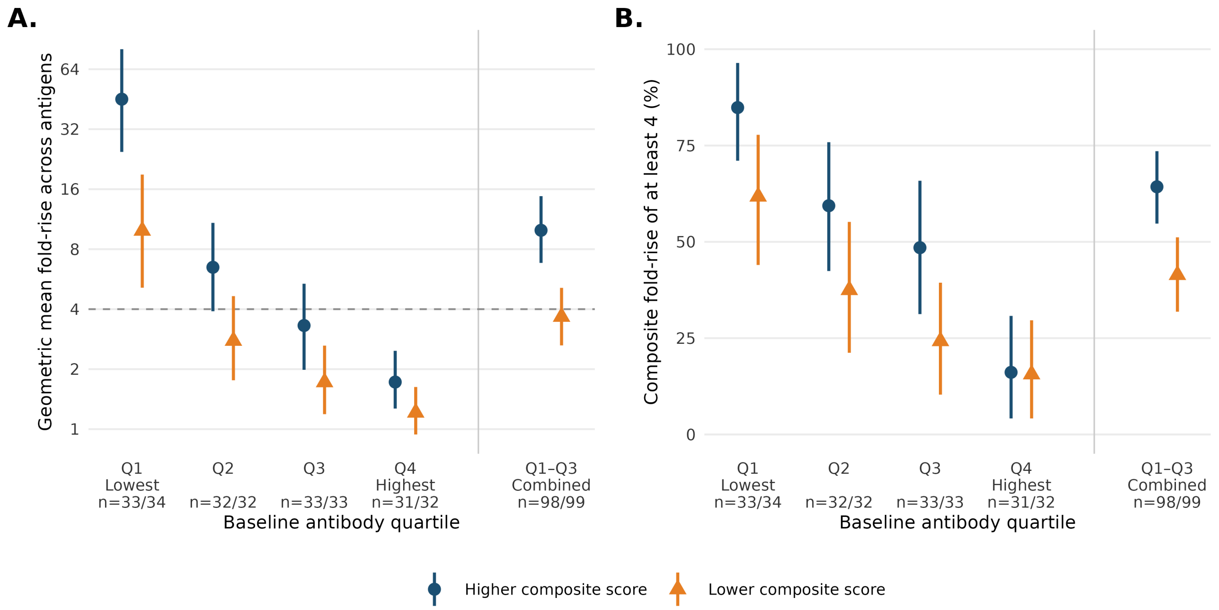


**Supplemental Figure S13: Observed composite antibody responses by composite cellular score and baseline antibody level.** Responses are shown as the geometric mean fold-rise across H1N1, H3N2 and B/Victoria. **A.** Geometric mean composite fold-rise. **B.** Percentage of recipients with a composite fold-rise of at least four. Within each baseline-titer quartile, recipients were split at the median composite score. Q1–Q3 combines the three higher-score and the three lower-score groups. Whiskers are recipient-bootstrap 95% CIs that hold fitted scores and score groups fixed. Tests that repeat model fitting and score grouping are in supplemental Table S5. Numbers give recipients in the higher and lower groups.

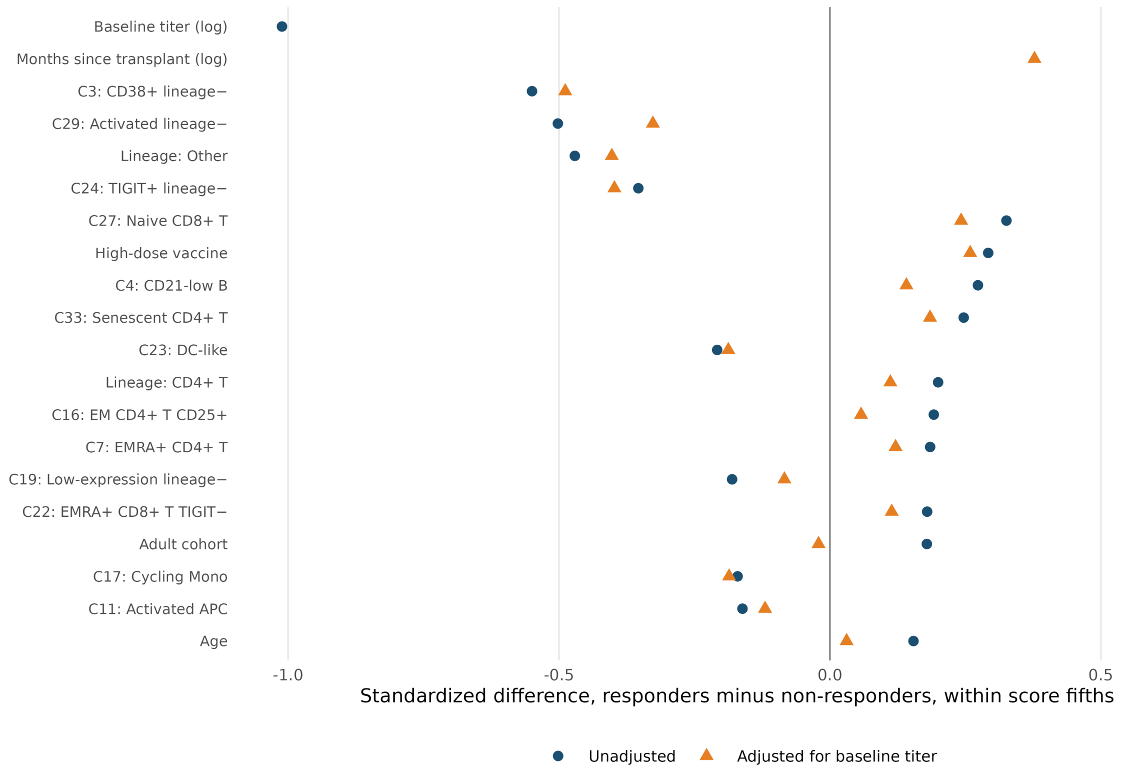


**Supplemental Figure S14: What differs between responders and non-responders with the same cellular score.** Standardized differences (responders minus non-responders) within fifths of the composite cellular score, pooled across fifths and weighted by fifth size, for the 20 features with the largest absolute difference. Navy circles are unadjusted; orange triangles residualize each feature on the log baseline titer within the fifth. Population shares enter as log shares. Positive values mean the feature is higher in responders.

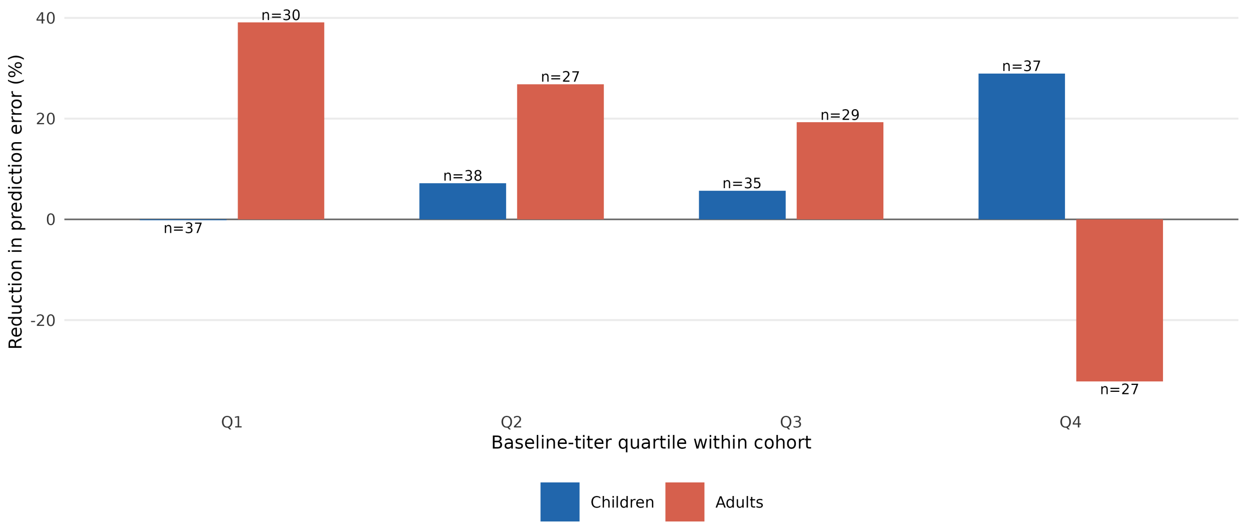


**Supplemental Figure S15: Prediction gains across baseline antibody levels in children and adults.** Bars show the reduction in mean squared error of the predicted composite response from adding immune-cell composition to baseline titers, within each cohort's baseline-titer quartiles. Positive values favor the model with immune-cell composition. Numbers give recipient counts. The models were refitted within each cohort on the shared 35-population atlas built from both cohorts; with about 30 recipients per cell these comparisons are exploratory.

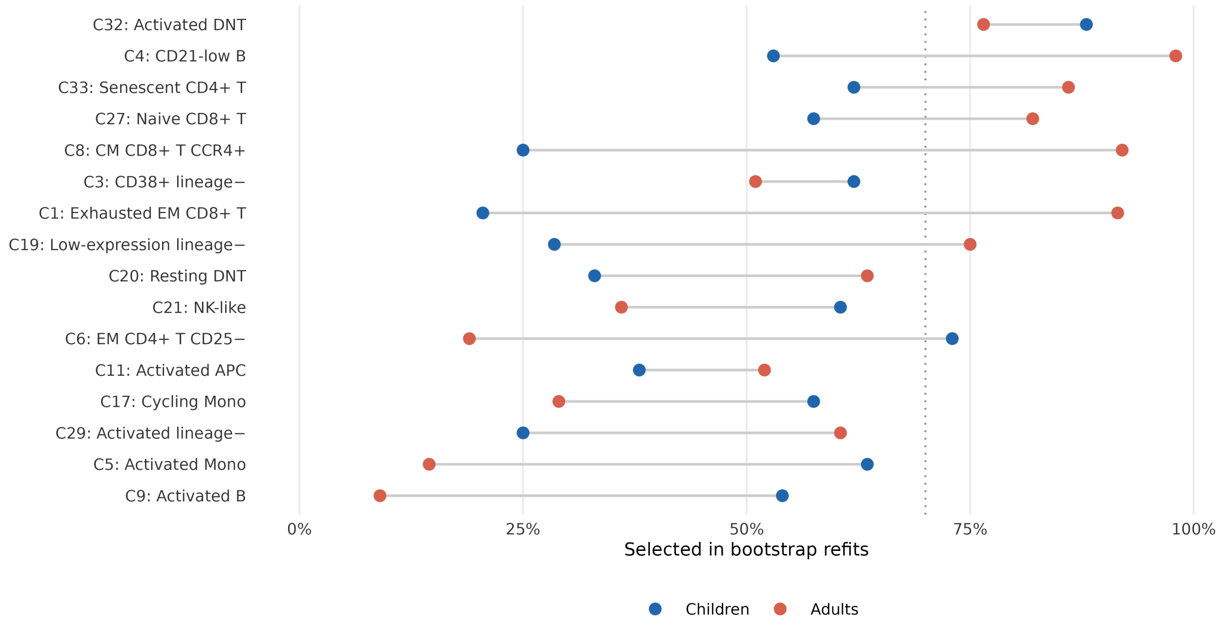


**Supplemental Figure S16: How often each population was selected in each cohort.** The model with baseline titers and immune-cell composition was refitted on the shared 35-population atlas in 200 bootstrap resamples of recipients within each cohort, with one penalty strength for both. Points give the percentage of refits in which a population had a nonzero coefficient; the populations shown are those reaching 50% in at least one cohort, and the dotted line marks 70%. Directions of association are in supplemental Table S6. Because the atlas is shared across cohorts, these comparisons are exploratory.

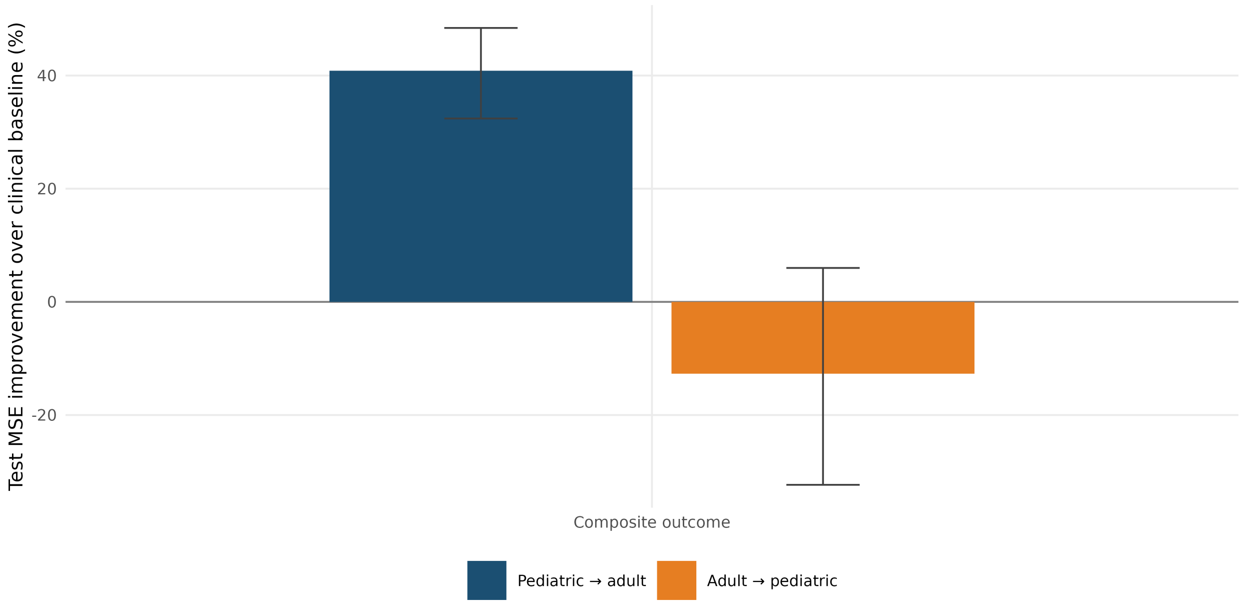


**Supplemental Figure S17: Prediction in one cohort using a model trained in the other.** Bars show the reduction in test mean squared error of the composite response for the cellular elastic net relative to an unpenalized model containing baseline titers and clinical factors, with 95% recipient-bootstrap intervals. Positive values favor the cellular model. Models were trained in pediatric recipients and tested in adults, or trained in adults and tested in pediatric recipients. The shared atlas was constructed from both cohorts, so these are exploratory transfer comparisons.

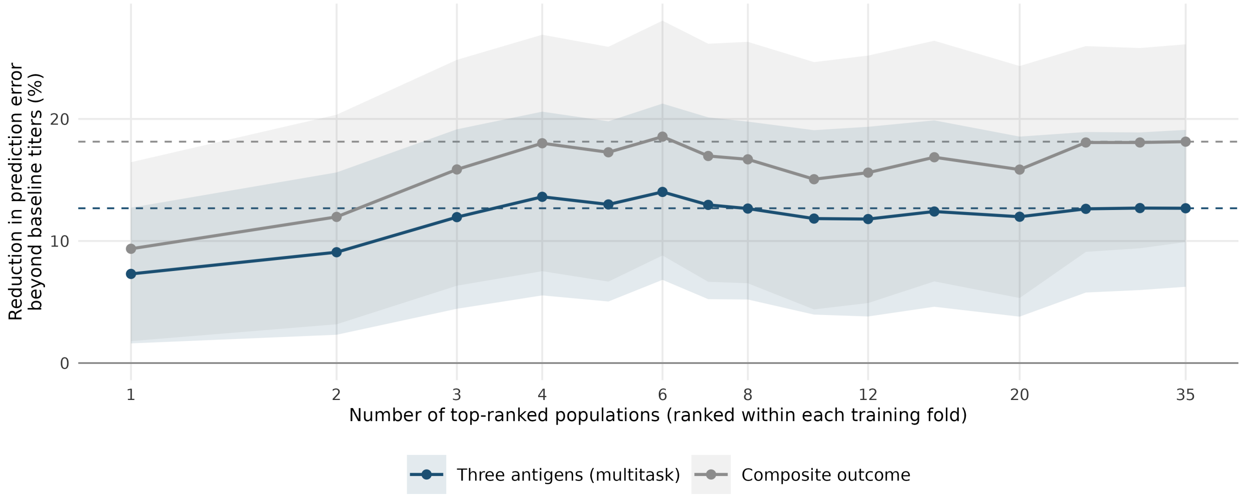


**Supplemental Figure S18: Prediction from the top-ranked populations.** Reduction in out-of-fold mean squared error beyond baseline titers on the shared atlas when M3 includes only the $k$ top-ranked populations. Populations were ranked inside each training fold by the magnitude of their bootstrap-averaged standardized coefficient. Dashed lines mark the gain with all 35 populations; ribbons are 95% recipient-bootstrap intervals.

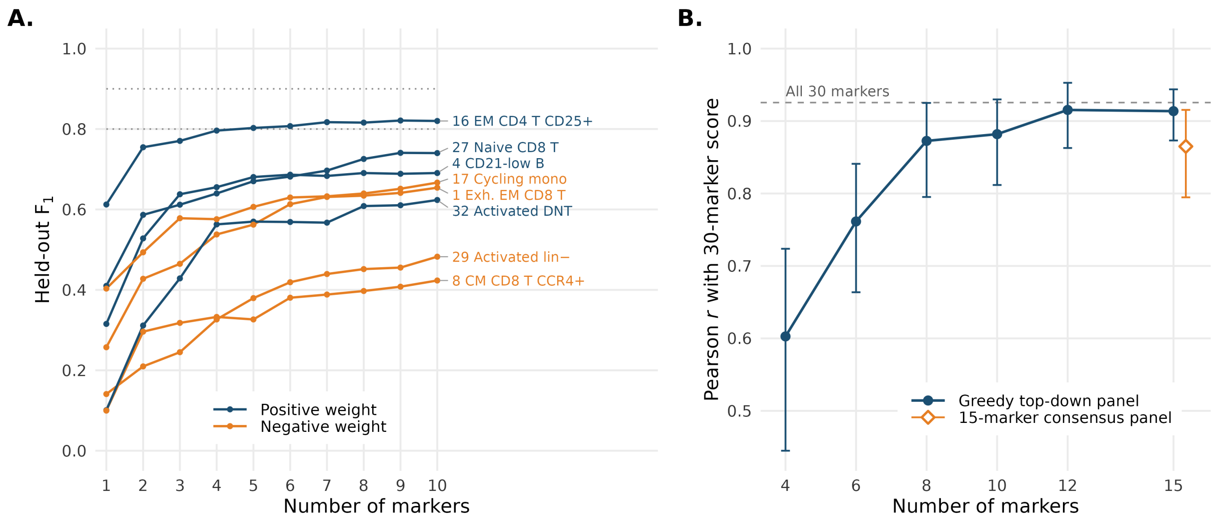


**Supplemental Figure S19: Top-down marker selection for the populations that drive the score.** Cells sampled from 78 held-out recipients were classified using markers chosen by greedy forward selection in training recipients. **A.** Held-out $F_1$ for identifying each of the eight populations that entered the six-population signature in at least one fold, by number of markers (one-versus-rest quadratic logistic regression); dotted lines mark 0.8 and 0.9. **B.** Pearson correlation between the composite score computed from reduced-marker cell labels (random forest over the 35 populations) and the 30-marker score from the same sampled cells, with 95% recipient-bootstrap intervals. The diamond marks the 15-marker consensus panel and the dashed line the 30-marker random-forest ceiling. Score weights come from a composite model fitted to all 260 first-season recipients and are descriptive.

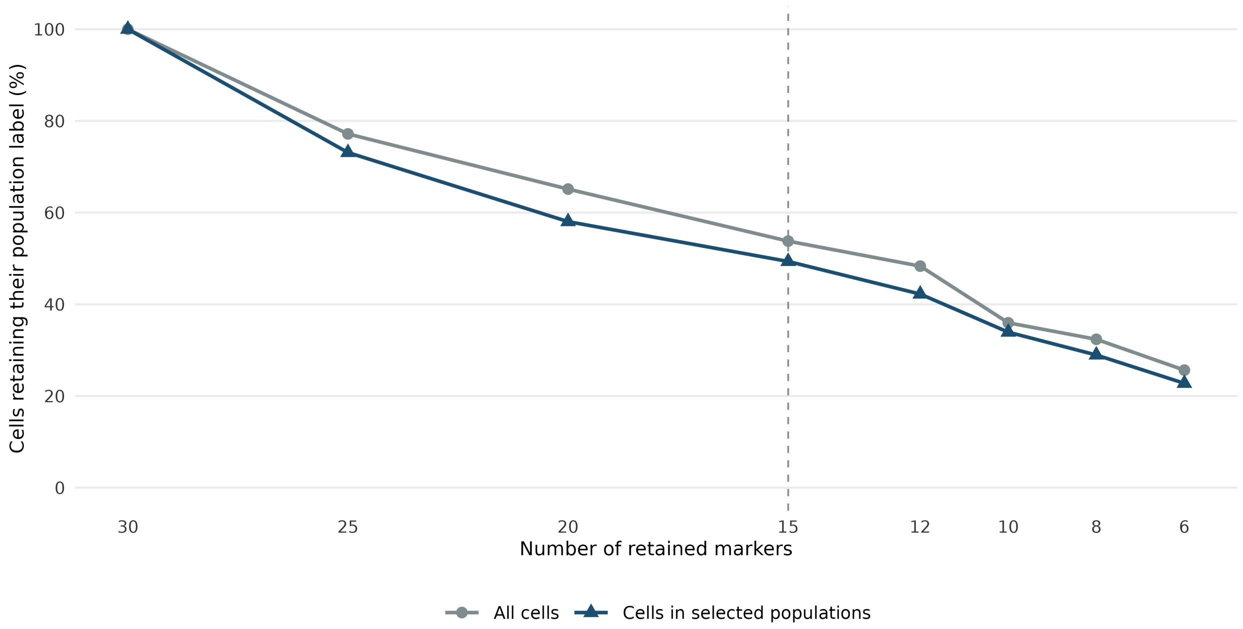


**Supplemental Figure S20: Cell-label agreement with fewer markers.** Cells were reassigned to the shared atlas using reduced marker sets. Curves show the percentage retaining their full-panel meta-cluster label, among all 200,000 sampled cells (gray) or the 39,059 cells belonging to the stably selected populations (blue). Stable selection means inclusion in at least 50% of 200 composite M4 bootstrap refits. The dashed line marks 15 markers, where agreement was 53.8% overall and 49.3% among cells in selected populations. This figure describes cell assignment; prediction error is shown separately in Figure 7.

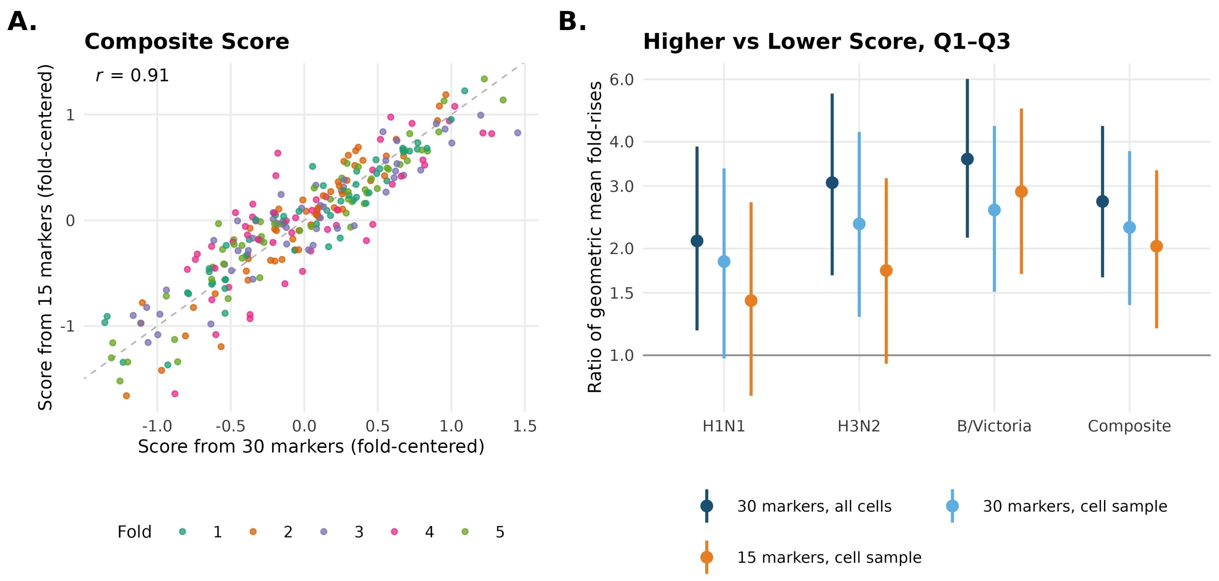


**Supplemental Figure S21: Cellular scores computed from 15 versus 30 markers. A.** Composite scores of held-out recipients computed from the same cell sample (up to 2,000 cells per episode) assigned with all 30 markers or with each fold's 15 top-ranked markers. Each cross-validation fold's mean score was subtracted. The dashed line marks equality. **B.** Ratio of geometric mean fold-rises between higher- and lower-score groups pooled across the lower three baseline-titer quartiles, for scores from all cells with 30 markers, from the cell sample with 30 markers and from the cell sample with 15 markers. Whiskers are recipient-bootstrap 95% CIs that hold fitted scores and score groups fixed.

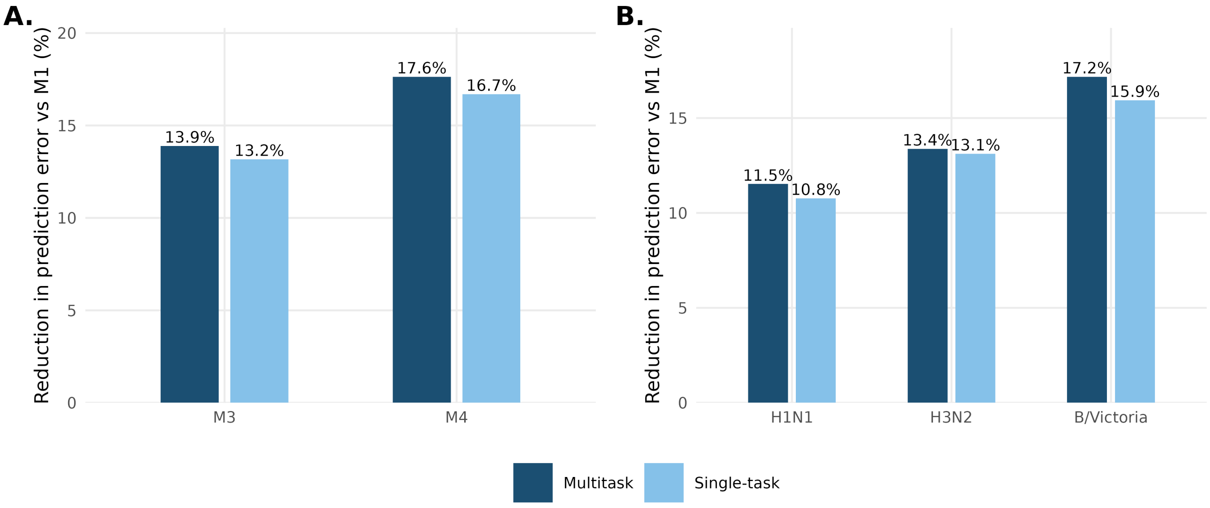


**Supplemental Figure S22: Joint and separate prediction of the three antigen responses.** This comparison uses all 334 episodes with complete titers (supplemental Section S3.7). Multitask elastic nets fitted the outcomes jointly; single-task elastic nets fitted each antigen separately. Both used the same outer folds and recipient-grouped penalty-tuning folds. **A.** Overall reduction in mean squared prediction error relative to baseline titers alone (M1). **B.** Antigen-specific reductions for the model adding immune-cell composition to titers (M3).

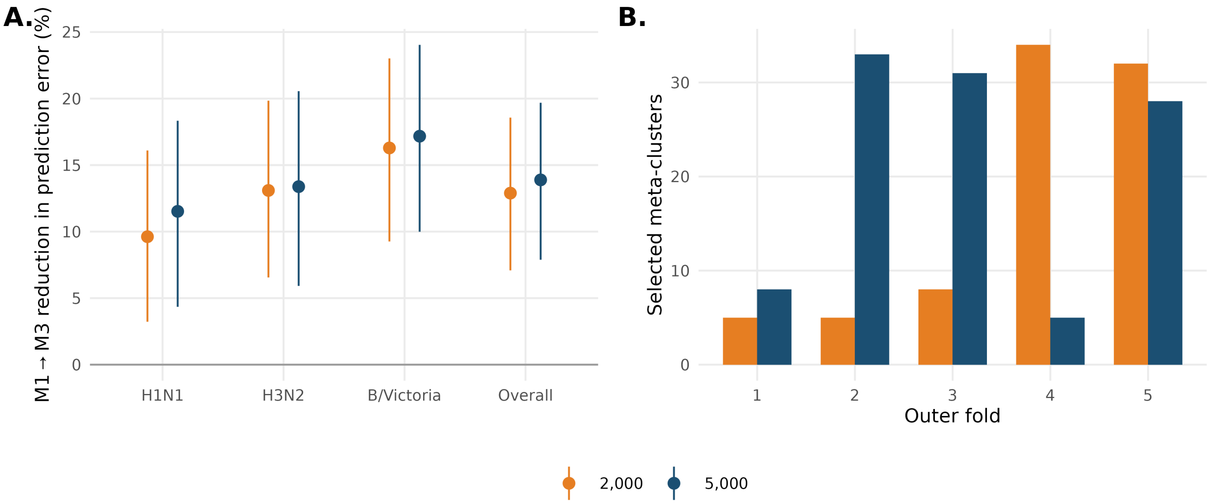


**Supplemental Figure S23: Sensitivity to micro-cluster count.** This comparison uses all 334 episodes with complete titers (supplemental Section S3.7). **A.** M1→M3 MSE reductions with bootstrap 95% intervals for 2,000 and 5,000 micro-clusters. Overall gains are 12.9% and 13.9%. **B.** Selected meta-cluster counts in the five outer folds. Both settings use the same validation procedure and episode-specific time since transplantation.

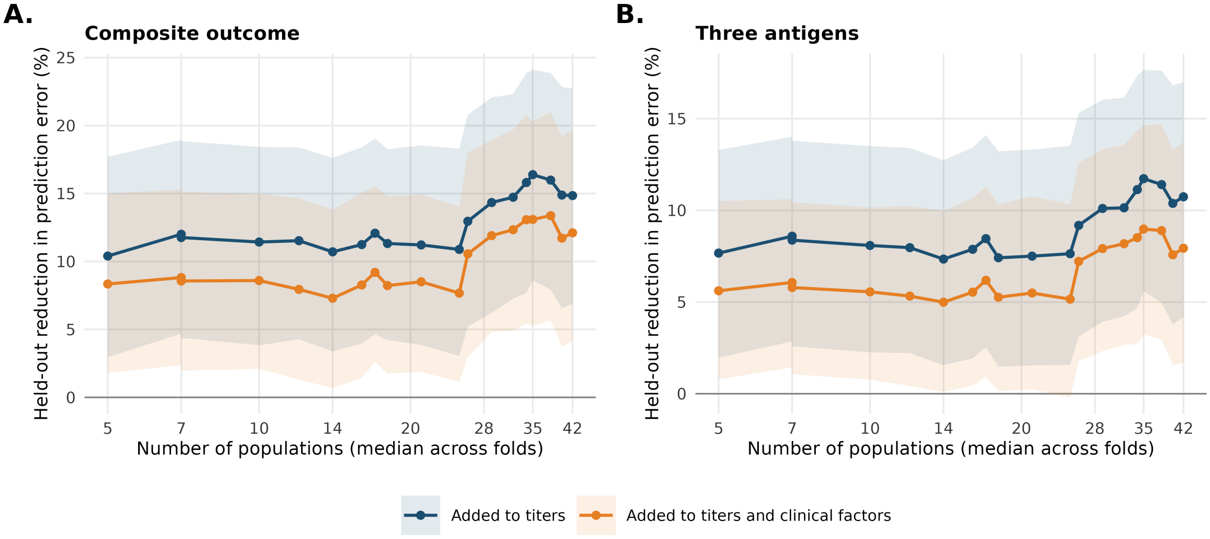


**Supplemental Figure S24: Prediction does not depend on how finely the cells are grouped.** Held-out reduction in mean squared prediction error from adding immune-cell composition, when the Leiden resolution, and with it the number of populations, is fixed at each value of the grid instead of being selected inside each training fold. **A.** Composite outcome. **B.** The three antigens modeled jointly. Points are the median number of populations across the five folds; ribbons are 95% recipient-bootstrap intervals. This figure is a diagnostic: the number of populations in the primary analysis was selected inside each training fold, never on held-out data.

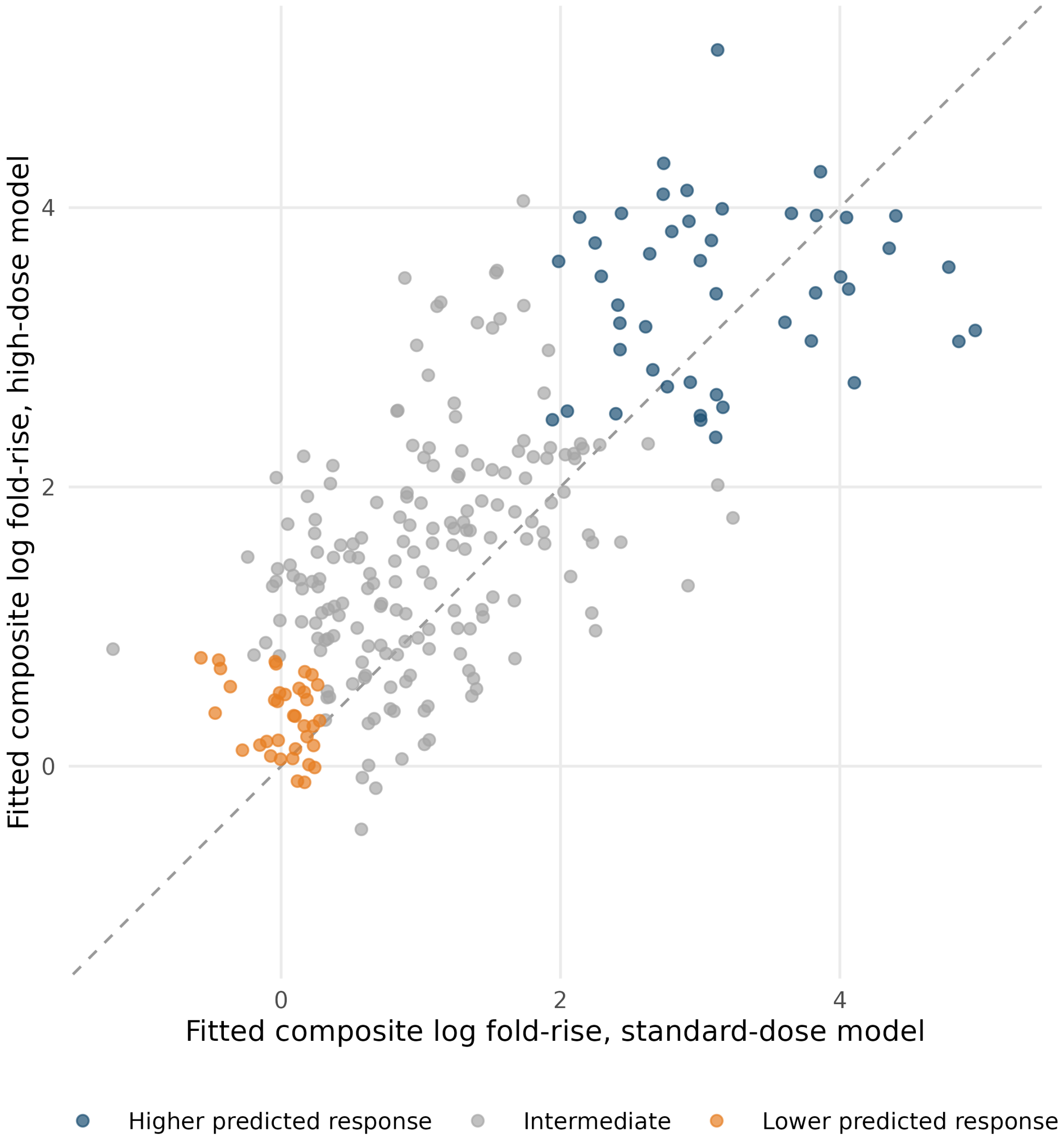


**Supplemental Figure S25: Fitted antibody responses under the two vaccine-dose models.** Axes show each recipient's fitted composite log fold-rise from separate random forests trained in standard-dose and high-dose recipients, each with 500 trees (Spearman $\rho = 0.74$ between the two). Orange identifies recipients in the lowest quartile under both models; blue identifies those in the highest quartile under both. The dashed line marks equal predictions. These partly in-sample predictions describe fitted patterns and do not estimate validated individual treatment benefits.

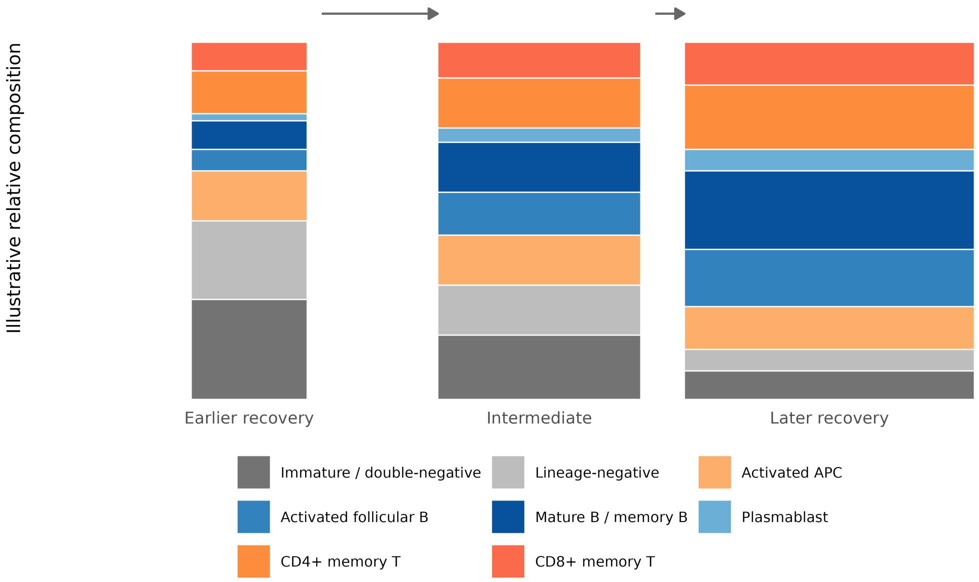


**Supplemental Figure S26: Illustration of possible changes during immune reconstitution.** The schematic depicts a proposed shift toward mature lymphoid populations as immune recovery progresses. Colored segments represent hypothetical relative abundances, and increasing bar widths illustrate possible expansion of the circulating cell compartment. Neither the proportions nor the widths are measured estimates from this study. The schematic provides biological context for the observed longitudinal associations.
